# What works for whom: a systematic review of inequalities in inclusion and effectiveness of social interventions for mental ill-health

**DOI:** 10.1101/2025.04.16.25325952

**Authors:** Anna Greenburgh, Helen Baldwin, Hannah Weir, Zara Asif, Dionne Laporte, Mark Bertram, Achille Crawford, Gabrielle Duberry, Shoshana Lauter, Brynmor Lloyd-Evans, Cassandra Lovelock, Jayati Das-Munshi, Craig Morgan

## Abstract

**Purpose:** People living with mental ill-health experience social and economic disadvantages, which contribute to poor outcomes and limit effectiveness of treatments. Interventions to improve social and economic circumstances have been developed, however, little is known about whether these interventions are effective for the most marginalised and disadvantaged groups, and those most in need of support.

**Method:** We conducted a systematic review in line with a pre-defined protocol to identify interventions to improve the social and economic circumstances of people experiencing mental ill-health. We included relevant records from two previous systematic reviews and updated their searches across four databases. We synthesised the intervention domains and locations of research, participant characteristics, and if effectiveness varied by participant gender, socioeconomic position, and race or ethnicity, and related indicators.

**Results:** We identified 266 relevant studies across 34 countries. Certain intervention domains were better researched than others (e.g. housing and employment vs. debt and social security advice). Participant characteristics were poorly reported and so understanding related to inclusiveness and generalisability of research is limited. Only 8% of papers reported any stratified results and statistical reporting standards were poor, limiting our ability to determine what works for whom. There was some indication that interventions are less effective for those in lower socioeconomic groups.

**Conclusion:** Improved reporting and representation of marginalised groups, stratified analyses of intervention data, and replication of results is needed to confidently draw conclusions about what works for whom in this field.

## INTRODUCTION

People who experience mental ill-health are, compared with the general population, typically more disadvantaged across multiple domains, including education, employment (1,2), housing stability and quality (3–5), income and finances (6,7), and social isolation (8,9). Further, those with severe mental ill-health (e.g. psychosis) are more likely to have been exposed, over the life course, to violence, trauma, and discrimination (10–12). These adversities increase risk of onset of mental ill-health and subsequent poor outcomes (1,8,13), contributing to an entrenched cycle of poor mental health and social exclusion. Furthermore, access to, and the effectiveness of, psychological and pharmacological treatment varies by indicators of socioeconomic position, thereby maintaining and widening inequalities in outcomes (14).

This relationship between social adversity and treatment outcomes highlights the need for mental health services to directly address the social and economic conditions of people with mental ill-health. Individuals with severe mental ill-health living in deprivation report significant unmet social needs (15), and failure of services to respond to such needs may contribute to lack of trust in services. This is particularly relevant to racial and ethnic inequalities in mental health systems. For example, in the UK, Black people with severe mental ill-health are more likely than people with severe mental ill-health from other ethnic groups to experience social and economic adversity (16–18) and this is further exacerbated by pervasive inequalities in access to mental health care (19–21). Therefore, the relationship between social adversity and poorer treatment outcomes means the failure of services to address social needs disproportionally disadvantages people from Black minoritised groups. Further dimensions of marginalisation within mental health care include social class, gender, and comorbid physical health and substance use problems, whereby the most socially excluded typically face multiple, intersecting disadvantages and occupy multiple marginalised statuses (22).

Interventions have been developed to interrupt this cycle of disadvantage by improving the social and economic circumstances of people with mental ill-health. Two recent systematic reviews synthesised this work (23,24), finding consistent evidence that Housing First (HF) and Individual Placement and Support (IPS) were effective in addressing housing and employment needs, especially for people with severe mental ill-health. Additionally, some studies indicated that family psychoeducation interventions and supported socialisation interventions were effective in improving social circumstances. Both reviews highlighted the lack of research in other domains, especially related to finances (e.g., debt and social security).

An important but neglected aspect of this research concerns which interventions work for whom and in what contexts. At present, it is unclear to what extent studies are inclusive of marginalised groups and those facing multiple forms of adversity, or indeed whether studies report the characteristics of their samples in sufficient detail to assess this. As the key aim of social interventions is to improve outcomes for those experiencing social and economic adversity, it is essential that they are effective in supporting those who experience the highest level of need, otherwise they risk maintaining or exacerbating systemic inequalities.

### Aims

We conducted a systematic review to:

i. Map the domains and contexts of research testing social interventions for people living with mental ill-health (severe mental illness and/or common mental disorders);
ii. Summarise the gender, ethnicity and socioeconomic status of the participants recruited within these studies.
iii. Assess how effectiveness of interventions varies according to these characteristics.

## METHODS

We conducted a two-staged systematic review in line with a pre-established review protocol. This study was delivered in partnership with an advisory board comprising people with lived experience of mental ill-health, service providers, third-sector workers, and academics who met regularly and made joint decisions about the research methodology, data analysis, and write-up.

We first identified studies included in two recent reviews (23,24). Both reviews included research detailing interventions which were designed to improve social and economic outcomes in adults with mental ill-health (see SI for full inclusion criteria).

We then updated these reviews to identify literature published between January 2020 and February 2024, searching MEDLINE, Web of Science, PsycINFO, and CENTRAL (see SI for full inclusion criteria and search strategies). In line with the previous reviews, our inclusion criteria focused on non-pharmacological interventions designed to improve social or economic circumstances of adults with a severe mental illness or common mental disorder in any one of the following domains: housing/homelessness; money and basic needs; work and education; social isolation and connectedness; family, intimate and caring relationships; victimisation and exploitation; offending; rights, inclusion and citizenship (see SI for full details).

We performed study selection for the updated search in duplicate (HB, AG, HW, ZA) at the title/abstract and the full-text screening stages. Individual researchers completed data extraction (see SI) which was checked by a second researcher (HB, AG, HW, ZA, DL). Conflicts at all stages were resolved through team discussion.

We extracted quality appraisals for studies included in the two previous reviews (i.e., Killaspy et al (2022) used the Kmett (25); Barnett et al (2022) used the Cochrane Rist of Bias tool for RCTs (26)). For studies identified in the updated search, we used the Kmet quality assessment checklist as this could be applied to both quantitative and qualitative studies. A random proportion (10%) of quality appraisals were conducted by a second researcher (HB, AG, HW, ZA).

For data synthesis, we first summarised participant characteristics (diagnoses, sex/gender, ethnicity or related categories and socioeconomic position), country of research, and domain of interventions categorised into nine different domains broadly in line with classification frameworks (27).

Where data were not available on ethnicity or race, we extracted and synthesised any data on other related concepts including nationality, immigration status, heritage and indigeneity. This was necessary as countries have varying legal frameworks regarding such data (for example reporting data relating to ethnicity or race is illegal in France). Estimating inclusion of marginalised groups with respect to race and ethnicity is complex not least due to variations in such legal frameworks, conceptualisations of such social constructs, and structural racism within health research (28,28–30). Language and understanding related to these concepts are continually changing and it is crucial for researchers to keep these concepts under review while researching health inequalities (31). In this paper, we report race and/or ethnicity data from individual studies using the language used in the respective studies.

We narratively synthesised studies reporting stratified analyses to assess whether effectiveness of interventions on social or economic inclusion outcomes varied by gender, ethnicity/race, or socioeconomic status and related indicators. For RCTs, we included any study where authors had assessed interaction effects between treatment condition and either gender, ethnicity/race, or socioeconomic status; or conducted subgroup analyses or responder analyses based on these sociodemographic categories. For non-RCTs, we considered any study where authors assessed whether the intervention-related changes in social or economic inclusion outcomes varied based on these sociodemographic categories, including any subgroup analyses. We also synthesised studies that tested interventions developed for broader populations but reported results only concerning any one of these sociodemographic subgroups (e.g. women). We supplemented our synthesis with a separate summary of results of studies exclusively including participants with a psychotic disorder (see SI).

## RESULTS

### Study Characteristics

We included 165 studies from Barnett et al (2022) and Killaspy et al (2022) after de-duplication (n=8) and removal of one meta-analysis. The updated database search identified a further 101 relevant records; therefore, 266 papers were included (See SI, Figure 1 for PRISMA and SI Table 3 for summary of study characteristics). For the studies identified in the updated search, Kmet quality appraisal scores ranged from 69%-100% for quantitative studies, and 65%-95% for qualitative studies.

**Figure 1.**
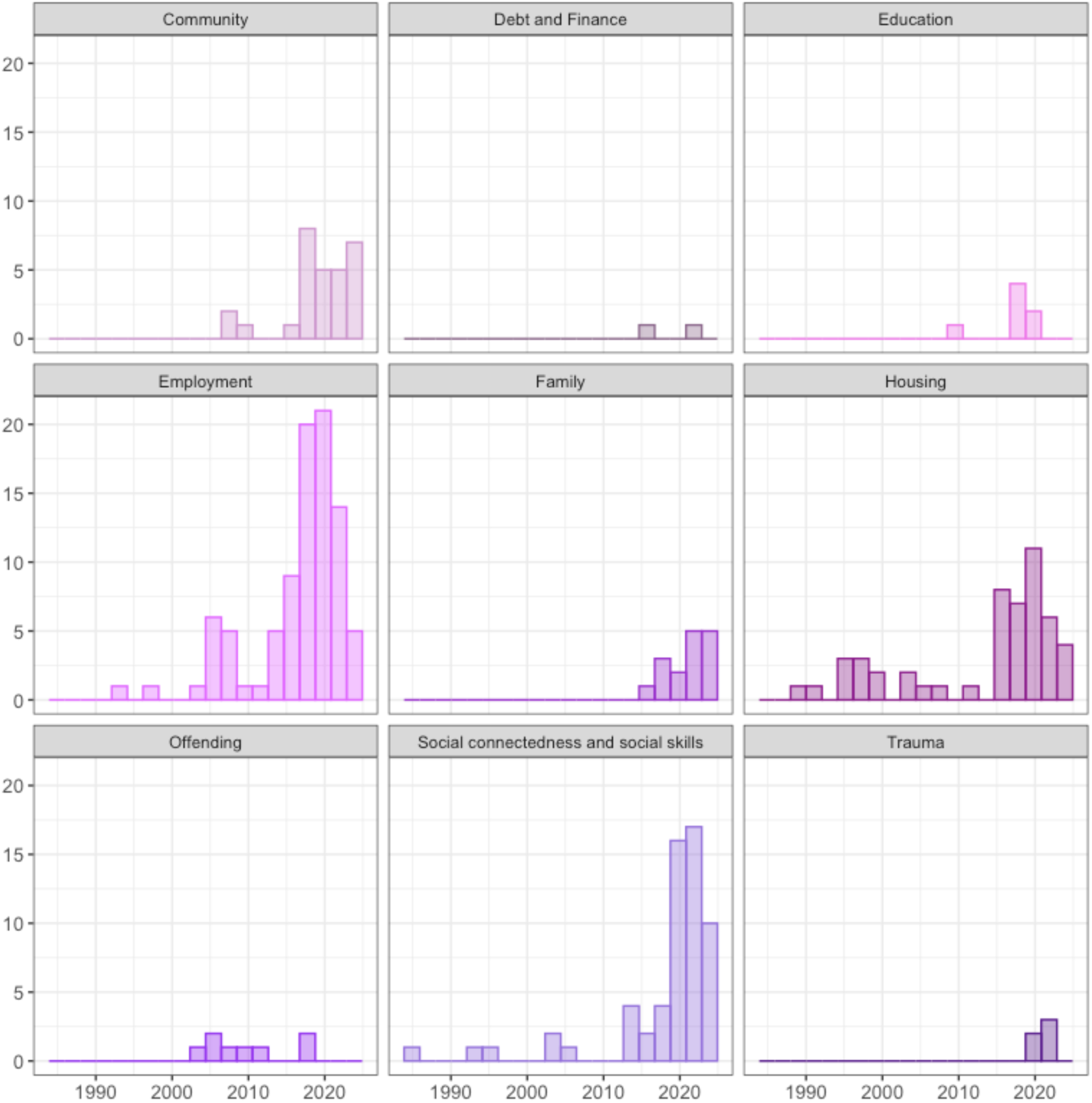
Histograms of primary life domains addressed by social intervention research over time. NB. Barnett et al (2022) included systematic reviews (published from database inception- February 2020) and randomised controlled trials (RCTs) (published from 2000 - August 2020); whereas Killaspy et al (2022) included any peer-reviewed paper reporting primary empirical data published between January 2016 and July 2020. The updated search included records from July 2020-February 2024.

#### Intervention domains

Interventions addressed: Employment (n=90; 34%), Social connectedness and social skills (n=65; 24%), Housing (n=50; 19%), Community support (n=31; 11%), Family (n=18; 7%), Education (n=11; 4%), Offending (n=9; 3%), Debt and Finance (n=2; 1%), and Trauma and Victimisation (n=5; 1%), where some addressed multiple domains (n=15) (see SI for examples of interventions in each domain).

#### Geographical location

The included studies were conducted across 34 countries - the majority in the USA (n=98) (Figure 2; see SI for full list). Most studies were conducted within an urban setting (65%; both urban-rural: 9%; rural: 2%; unknown: 24%).

**Figure 2.**
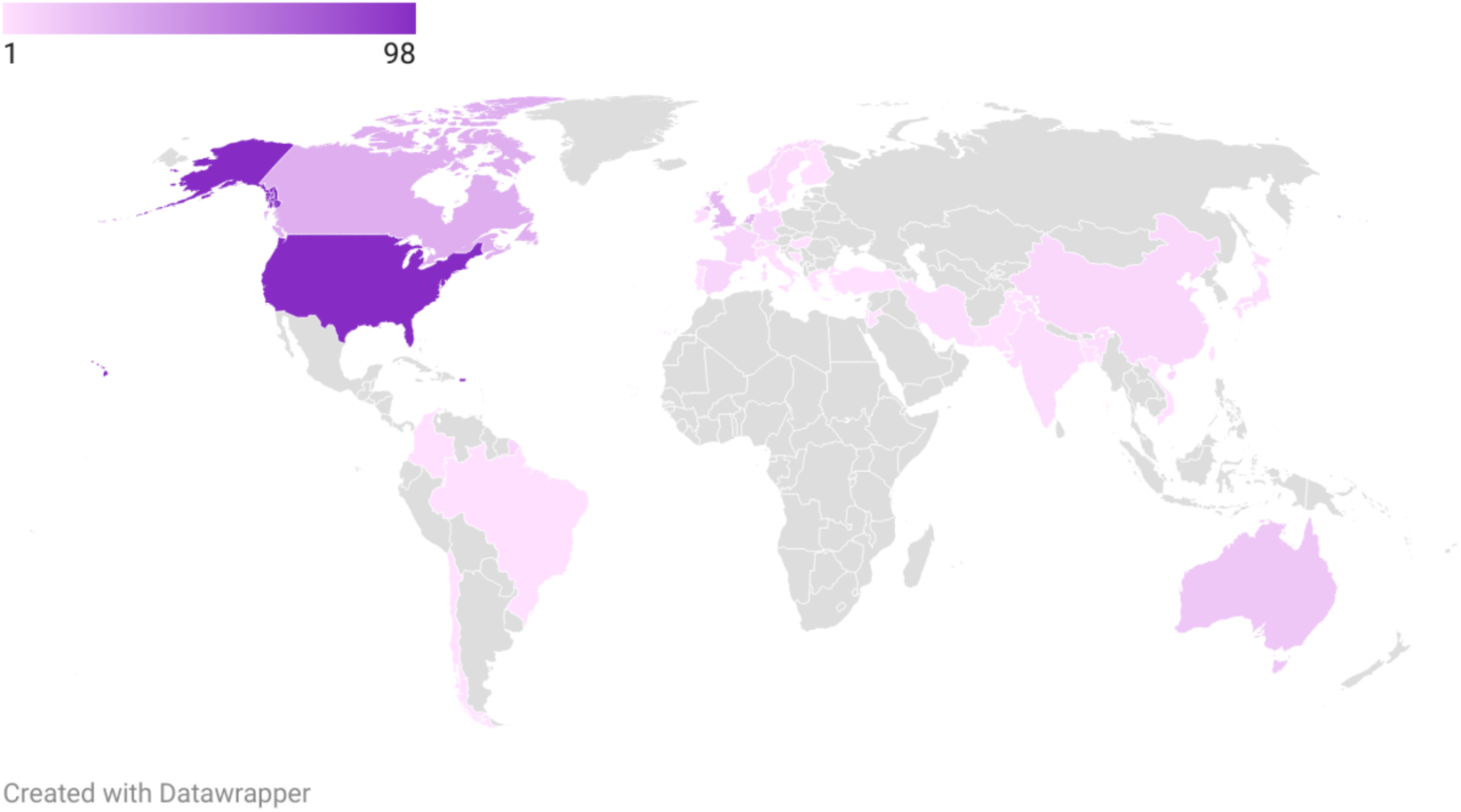
A heatmap of the geographical locations in which the research was conducted, where the count represents the number of academic manuscripts deriving from each country.

### Participant characteristics

No studies reported data on intersecting dimensions of marginalisation.

#### Gender

Most studies reported a male/female dichotomy only: just 10 of the 266 studies (4%) reported on inclusion of individuals who identified as non-binary, transgender, gender-free or ‘other’. Fewer women were involved in research than men; the average inclusion of women was 43% (SD=22%), compared with 58% for men (SD=22%). However, this may in part reflect variations in the prevalence of mental health problems by sex, e.g., higher proportion of men experience psychosis.

#### Ethnicity, race, migration status, nationality, indigeneity, heritage

We found a range of relevant indicators were used in included studies, including ethnicity, race, nationality, migration status, indigeneity, and heritage. We extracted information as it was reported in each study – and it was largely unclear whether data pertained to participants’ self-ascribed identities or researcher observations.

Overall, 126 studies (47%) did not include any data on ethnicity, race, nationality, migration status, indigeneity, heritage or related indicators. For the 140 studies (53%) that did report on such data, there was variation in methods used. For example, research in some countries (e.g. France) was restricted to only reporting nationality according to a binary (i.e. French national/not); in the UK and USA, ethnicity and race statistics were typically reported according to pre-defined census categories. Studies in the UK focused on the concept of ethnicity; studies in the USA typically reported statistics categorised by racial and/or ethnic groups.

In the UK and USA, ethnicity and/or race were reported by 104 of the 119 (87%) studies conducted in these countries. We were able to calculate pooled statistics for representation of ethnic and racial groups in these countries as most studies reported data according to consistent categories (see Tables 1 and 2). However, these pooled statistics are only based on studies which report this data, and are therefore likely to be inflated estimates, as studies which do not include any given ethnic group are more likely not to report any data. For example, only seven of the 21 studies conducted in the UK report including any Black participants and therefore it is possible that the remainder did not include any Black participants (see Table 1).

Although systematically assessing representativeness was beyond our aims, it is important to interpret these results in light of sociodemographic variations in prevalence of mental ill-health and service use – where the discrepancy between these two factors is commonplace given that many people face barriers to accessing support. For example, in the UK, prevalence of psychotic disorder is higher among Black men than men from any other ethnic group and common mental disorders are higher among Black women than women from other ethnic groups; however Black people are much less likely to access treatment (Adult Psychiatry Morbidity Survey, 2014). The question of representativeness of the studies included in this review is complicated by contextual differences – whereby ‘typical’ population sociodemographics vary by geographical regions, diagnoses, and service types. Indeed, the studies included in this review span a great number of contexts and participant populations. Therefore, it is not possible to robustly assess representativeness from the results presented here. Nevertheless, the majority-White composition of the included studies (see Table 1 & 2) indicates that this work may not be generalisable to many service contexts.

**Table 1.** A summary of race and ethnicity representation across studies conducted in the United Kingdom (n=21). These statistics are calculated only from studies reporting data in these respective census categories. Studies not reporting data were excluded from these calculations.

|  | ***Asian or***<br>***Asian***<br>***British*** | ***Black, Black British,***<br>***Caribbean or African*** | ***Mixed or multiple***<br>***ethnic groups*** | ***White***<br>***(incl. Roma)*** | ***Other ethnic group***<br>***(Arab/ any other)*** | ***Unknown/ rather***<br>***not say*** |
| --- | --- | --- | --- | --- | --- | --- |
| ***N studies***<br>***reporting*** | 6 | 7 | 5 | 13 | 9 | 2 |
| ***Mean (%)*** | 9.04 | 28.04 | 10.41 | 70.85 | 12.59 | 0.63 |
| ***SD (%)*** | 9.06 | 22.93 | 11.14 | 24.24 | 14.67 | 0.53 |
| ***Range*** | 1.75-26.5 | 6-70.97 | 1.5-29.03 | 17.4-100 | 0.75-50 | 0.25-1 |
| ***IQR*** | 5.5 | 20.95 | 8.7 | 26 | 6 | 0.36 |
| ***Median*** | 6 | 18 | 8 | 72.2 | 10 | 0-0.62 |

**Table 2.** A summary of race and ethnicity representation in studies conducted in the United States of America (n=98). These statistics are calculated only from studies reporting data in these respective census categories. Studies not reporting data were excluded from these calculations.

|  | **Race** |  |  |  |  |  |  | **Ethnicity** |  |  |  |  |
| --- | --- | --- | --- | --- | --- | --- | --- | --- | --- | --- | --- | --- |
|  | *White/*<br>*Caucasian* | *Black or*<br>*African*<br>*American* | *American*<br>*Indian and*<br>*Alaska*<br>*Native* | *Asian* | *Asian/*<br>*Pacific*<br>*Islander* | *Race:*<br>*Native*<br>*Hawaiian*<br>*and Other*<br>*Pacific*<br>*Islander* | *Two or*<br>*more*<br>*races* | *Hispanic or*<br>*Latino* | *White*<br>*alone, non*<br>*Hispanic or*<br>*Latino* | *Black non-*<br>*Hispanic* | *Non-*<br>*Hispanic* | *Race/*<br>*ethnicity*<br>*unknown/*<br>*other* |
| **N studies**<br>**reporting** | 78 | 69 | 8 | 19 | 3 | 2 | 14 | 53 | 5 | 2 | 6 | 42 |
| **Mean (%)** | 47.00 | 42.21 | 3.85 | 3.64 | 2.89 | 3.5 | 11.52 | 13.43 | 65.62 | 35.55 | 84.32 | 19.13 |
| **SD (%)** | 20.59 | 23.21 | 2.58 | 4.97 | 3.21 | 0.71 | 11.70 | 11.13 | 28.04 | 7.71 | 8.33 | 20.34 |
| **Range** | 10.34-94.6 | 2.5-86.2 | 1-9.46 | 0-21 | 1-6.6 | 3-4 | 0.5-38 | 0-53 | 27-97 | 30.1-41 | 76-95.9 | 0.01-81 |
| **IQR** | 34.90 | 40 | 1.84 | 1.96 | 2.80 | 0.5 | 11.26 | 15.46 | 35.6 | 5.45 | 11.99 | 25 |
| **Median** | 49 | 41 | 3.5 | 2.7 | 1.08 | 3.5 | 8.95 | 12.5 | 64.7 | 35.55 | 81.27 | 10 |

It is difficult to assess inclusion of specific groups in countries beyond the UK and USA, due to lack of consistent reporting. Of the 149 studies conducted elsewhere, 41 (28%) reported data on ethnicity, nationality, and indigeneity (or related indicators). Many of these were conducted in Canada (n=17), and many of these (n=9/17) reported this data according to binary categorisations (53%) (e.g. “White” v “non-White”). For studies conducted beyond Canada, the UK and the USA, 17 (81% of the 21 studies reporting relevant data) studies reported data only according to a binary. Due to these limitations in the data, we crudely estimated the inclusion of people from minority groups based on ethnicity, race, nationality, heritage, indigeneity and related factors all together in research in each country (see SI table 5).

#### Socioeconomic position

Indicators of socioeconomic position (including social class, occupation, tenure, financial situation, and education (32)) were reported in 208 studies (78%). There was considerable heterogeneity in which indicators were used and studies often reported more than one indicator; the most common proxy was education (n=148), followed by employment or vocation status (n=65), homelessness (current/lifetime) (n=33), income (n=22), receipt of welfare benefits (n=20), living status (i.e. owning home, private rental, social housing etc) (n=11), geography-specific socioeconomic stratum (n=4), debt (n=2), neighbourhood factors including the UK Index of Multiple Deprivation (n=2), availability of food each day (n=1), parental socioeconomic status (n=1), housing stability (n=1), and savings (n=1). Due to the heterogeneity of the socioeconomic indicators used, and the variation in upper/lower bounds of socioeconomic disadvantage across geographies, it was not possible to define and assess inclusion of socioeconomically marginalised groups in a meaningful way across studies, and so we were not able to provide pooled summary statistics regarding the inclusion of the most socioeconomically marginalised groups here. This calls for standardised reporting processes of socioeconomic adversity in future studies.

#### Dual-diagnosis and comorbid physical health problems

Fifty-four studies (20%) reported substance use or abuse (current and/or lifetime) as an exclusion criterion to participation. Where the proportions with current substance abuse or dependence were reported (n=69), this ranged from 1-100% (mean=56%, SD=32%). Comorbid physical ill-health data were reported less frequently (n=27) and five studies excluded participants on this basis.

### Stratified analyses

Very few papers (n=20; 8%) reported stratified results by gender, race and/or ethnicity, or socioeconomic position. Some studies reported multiple stratified analyses (29 analyses reported in total). No studies stratified results by multiple intersecting dimensions of marginalisation (e.g. women from low-income backgrounds). Most analyses were conducted on data from RCTs (n=16) and many exclusively included people with severe mental illness (n=13) (see Table 3; SI Table 6 for further information).

Many factors limit our ability to draw conclusions about variation in effectiveness of social interventions for the different groups examined. There was no replication of stratified analyses as each pertained to studies of different interventions and settings. Further, authors reported sufficient effect size data to interpret the results in less than half of stratified analyses (45%; 13 of 29); rather, if effects were found to be not ‘significant’, researchers often did not report any data. This is problematic with respect to the likely underpowering of such analyses.

Nevertheless, emerging evidence indicates that effectiveness of interventions may vary for different groups. People from lower socioeconomic groups seem to benefit less from some interventions. The extent to which outcomes vary by socioeconomic or sociodemographic group likely depends on the specific domains, designs and contexts of interventions. However, given the sparsity and limitations of existing evidence, it is not yet possible to investigate these patterns.

#### Gender

Fifteen studies (5%) stratified analyses by gender. This included one non-controlled prospective study of an employment intervention (33), one quasi-experimental controlled study of a social connectedness intervention (34), and 13 RCTs (see Table 3). Of the 13 RCTs, only 5 reported effect sizes data regarding the possible moderating effect of gender on the impact of the intervention. Two found a stronger effect for men (35,36), one reported a subgroup analysis finding evidence for effectiveness in women (37), and two studies reported data consistent with a range of interpretations (38,39). For example in a ‘strong’ quality-assessed, two-arm, multicentre RCT, which tested the efficacy of a psychoeducational intervention programme designed to reduce caregiver burden for carers of people with schizophrenia (39), a subgroup analysis suggested that the intervention may be more effective for women (i.e., there was a moderate association with burden reduction among women but not men). However, data were statistically consistent with parameter values ranging from a considerable level of reduced risk to a considerable level of increased risk of burden reduction (Coefficient (95%CI): Female: 0.14 (-0.12- 0.41)).

There was no replication within these 5 RCTs: they all tested different interventions across the domains of housing, employment, social connectedness and family relationships, and many suffered limitations such as small number of women included (36).

#### Minoritised ethnic groups and Indigenous peoples

We identified only 3 studies (1%) that stratified analyses by ethnic group or related indicators. These included one qualitative study of a Housing First intervention in Canada (40), one RCT of independent vs. staffed group living for homeless people in the USA (41), and one RCT of social skills training for people with schizophrenia (42) (see Table 3). This is too limited an evidence base to draw any conclusions about variation in effectiveness of social interventions by minoritised ethnic or racial group. Nevertheless, the RCT of independent vs. staffed group living for homeless people did find that, at 18- month follow up, African American and Hispanic participants experienced on average 37 more days of homelessness than White participants, across both conditions (an average of 59 more days of homelessness in the independent living condition; and 15 more days in the staffed condition). This suggests poorer outcomes in both intervention arms for minoritised populations, but that this inequality may be reduced in staffed living interventions. However, this is a single study, rated as having a high risk of bias. The other RCT of social skills training reported that there was no ‘significant’ interaction effect by ethnic group, making it impossible to assess whether there was any hint of a variation in effect.

#### Socioeconomic position

Eleven studies (4%) stratified results by indicators of socioeconomic position. Three of these were non-RCTs, examining interventions in the domains of housing (43), education (44), and social connectedness (34). None of these studies reported sufficient information on effect size and/or group means to interpret their results.

Evidence from 4 RCTs indicated that interventions were less effective for people in lower socioeconomic groups (see Table 3). These interventions spanned the domains of victimisation, debt and finances, employment, and family relationships. For example, in one study rated as strong quality, responder analysis of a vocational peer support intervention found that participants receiving social security payments were less likely to be classified as responders, i.e. become employed at any time during the 12-month follow-up (those receiving social security payments comprised 83% of non-responders but 52% of responders) (47).

One further RCT was inconclusive regarding whether an employment intervention was more effective for people in higher socioeconomic positions, as data were statistically consistent with parameter values ranging from lower to higher likelihood of intervention success (e.g. OR=0.67, CI=0.32-1.40). However, although interventions may be less effective for those facing more challenging socioeconomic conditions, this strong quality RCT highlighted that this inequality is reduced in the intervention condition compared with treatment as usual: the impact of previous work history on vocational recovery outcomes was lower for participants receiving IPS compared with those receiving service as usual (38).

Three further RCTs reported stratified analyses. However, they did not report sufficient accompanying data to interpret their analyses.

**Table 3.**
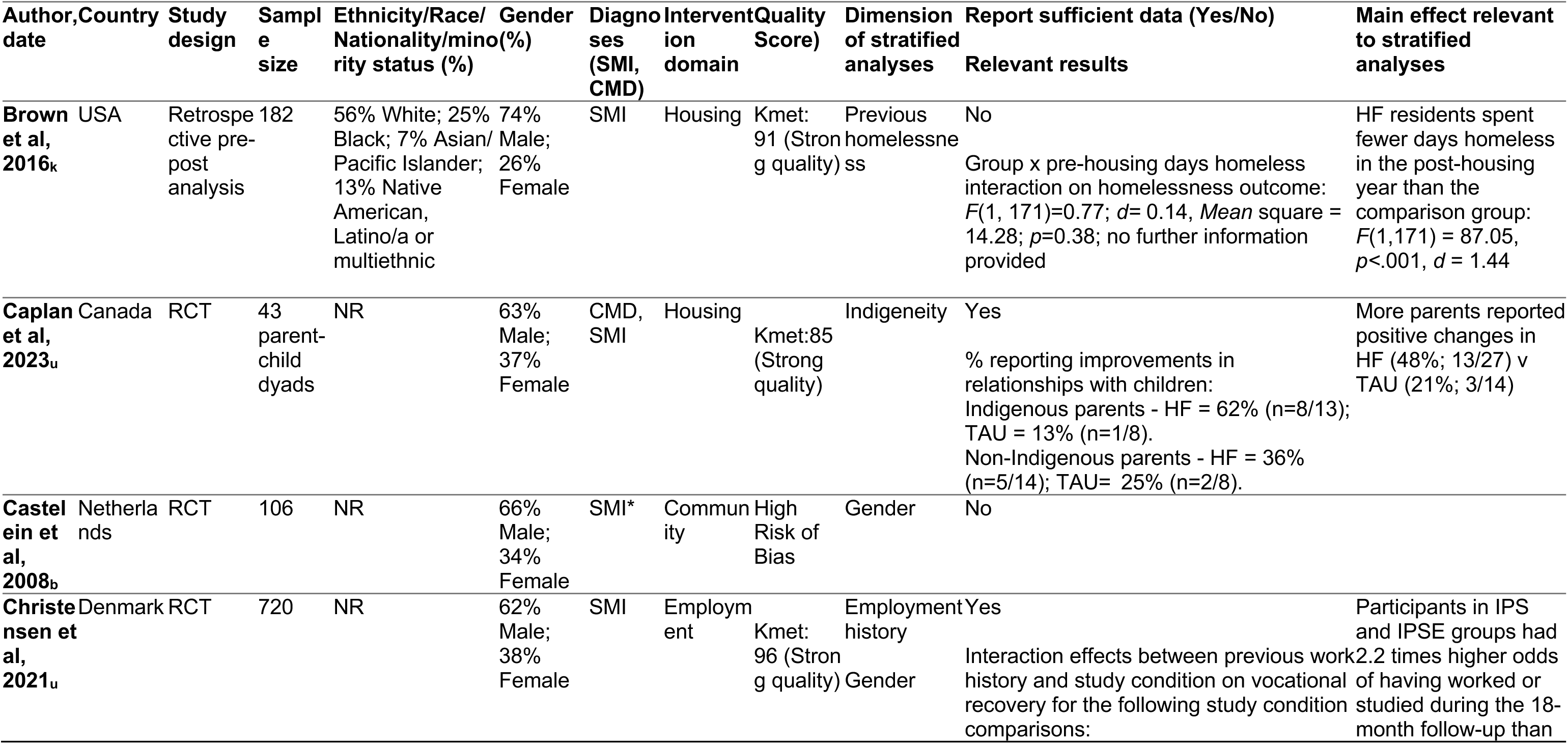
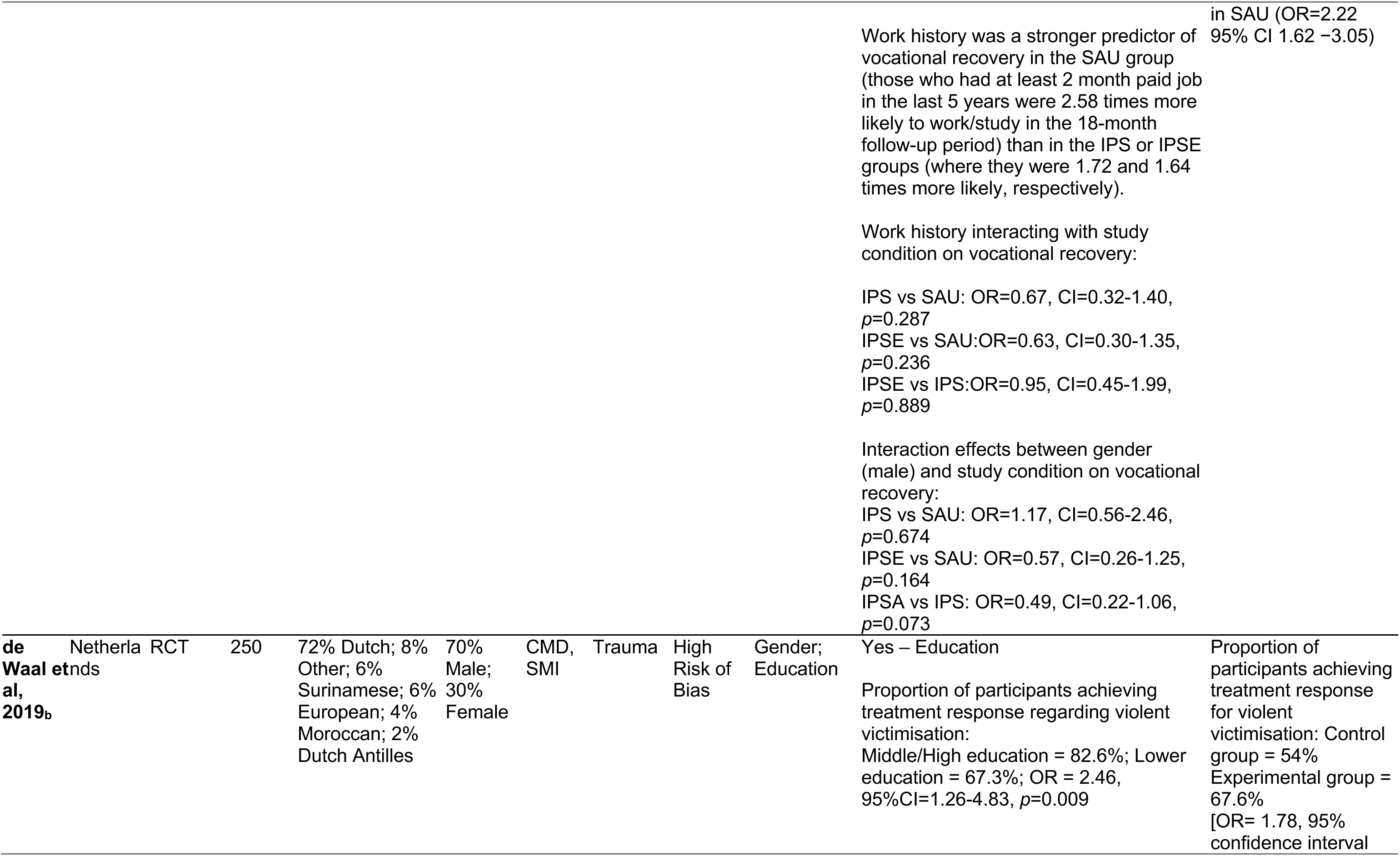
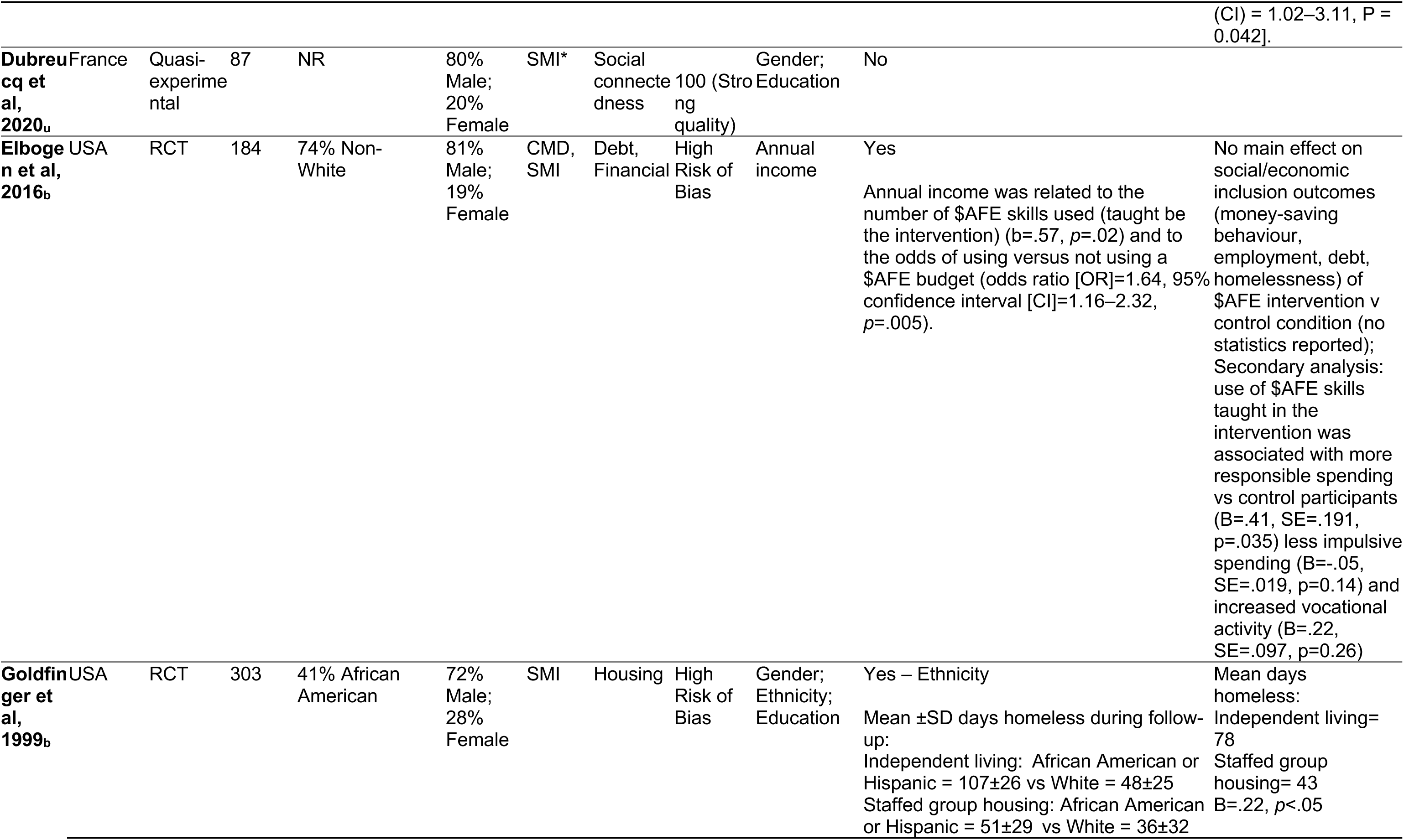
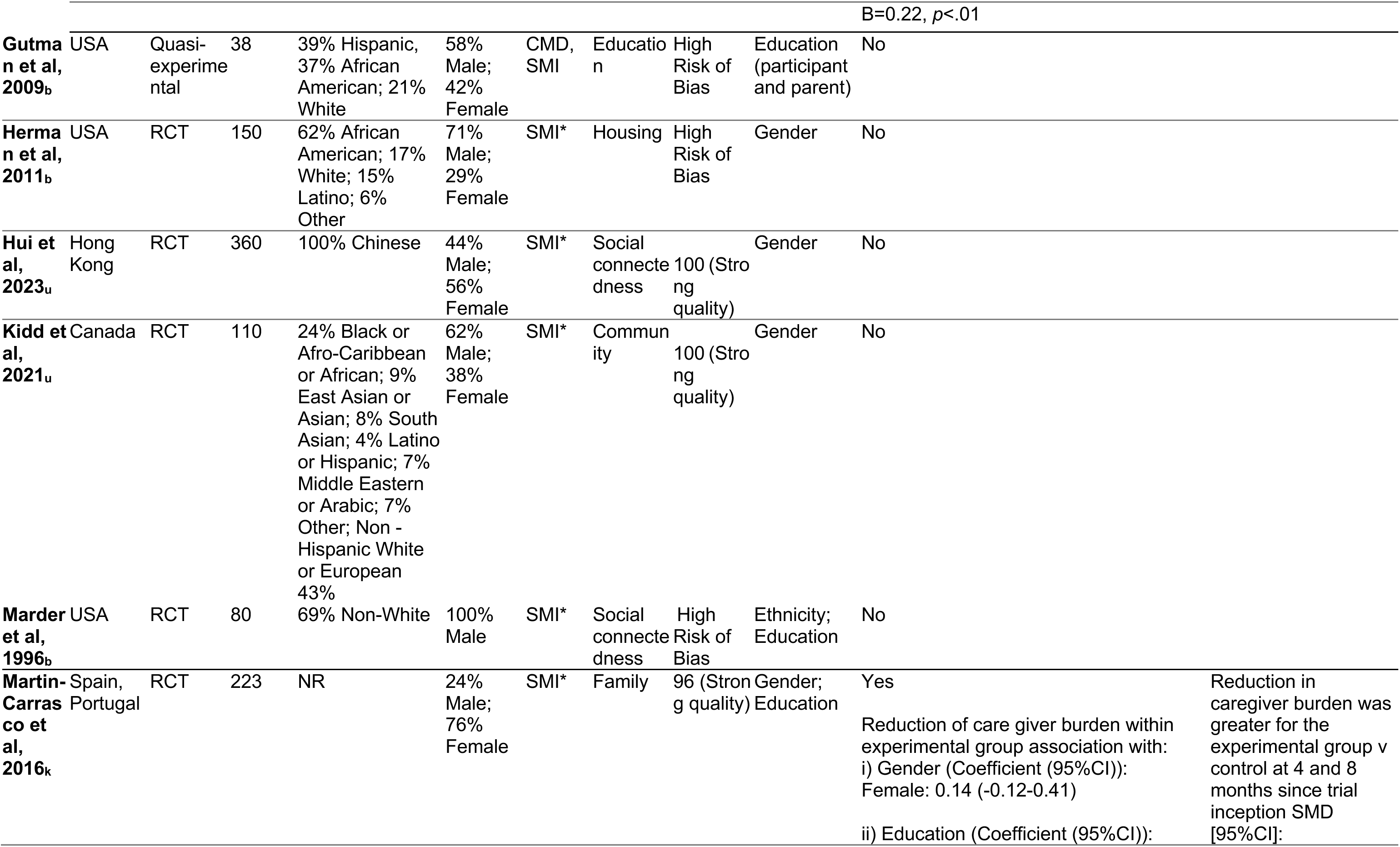
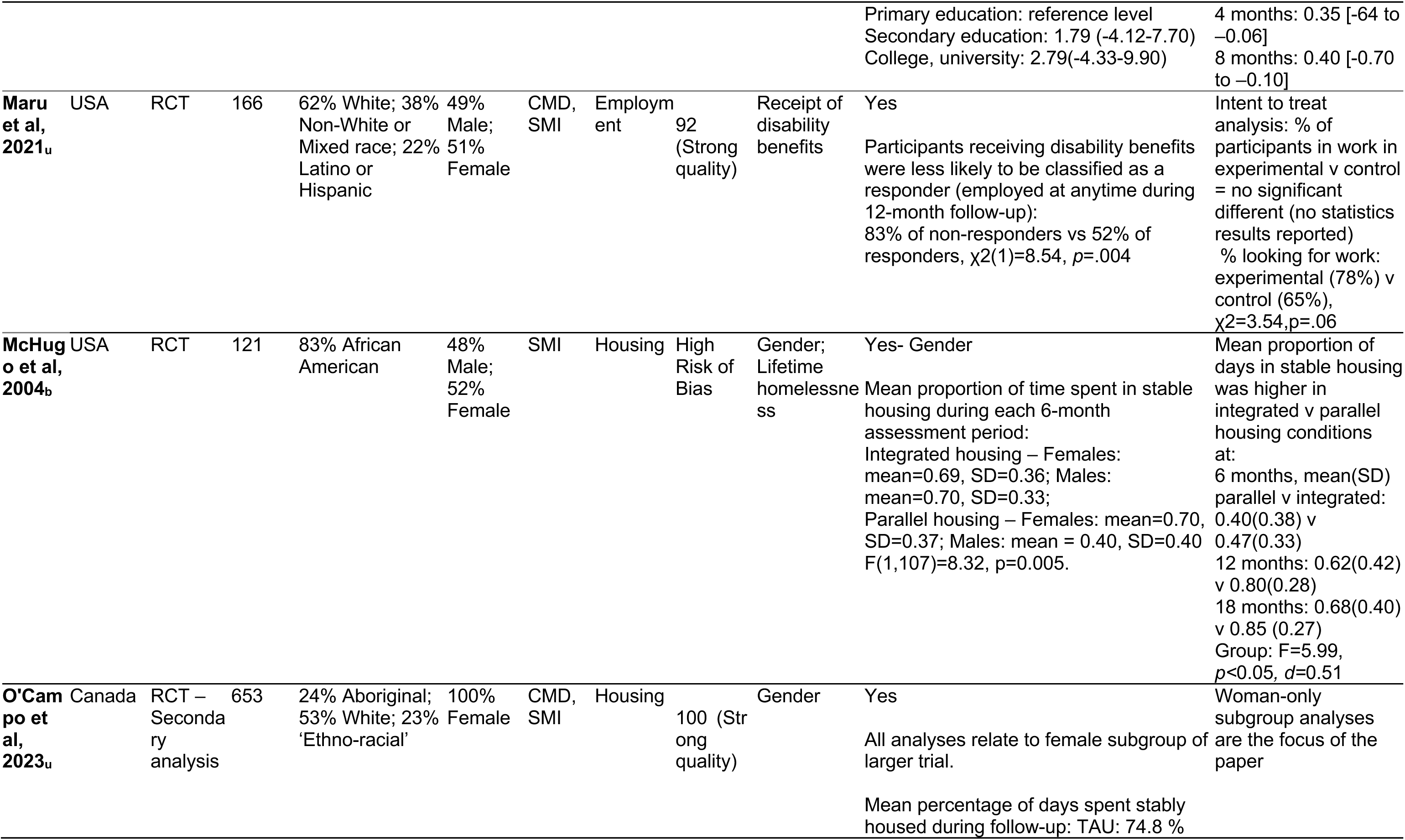
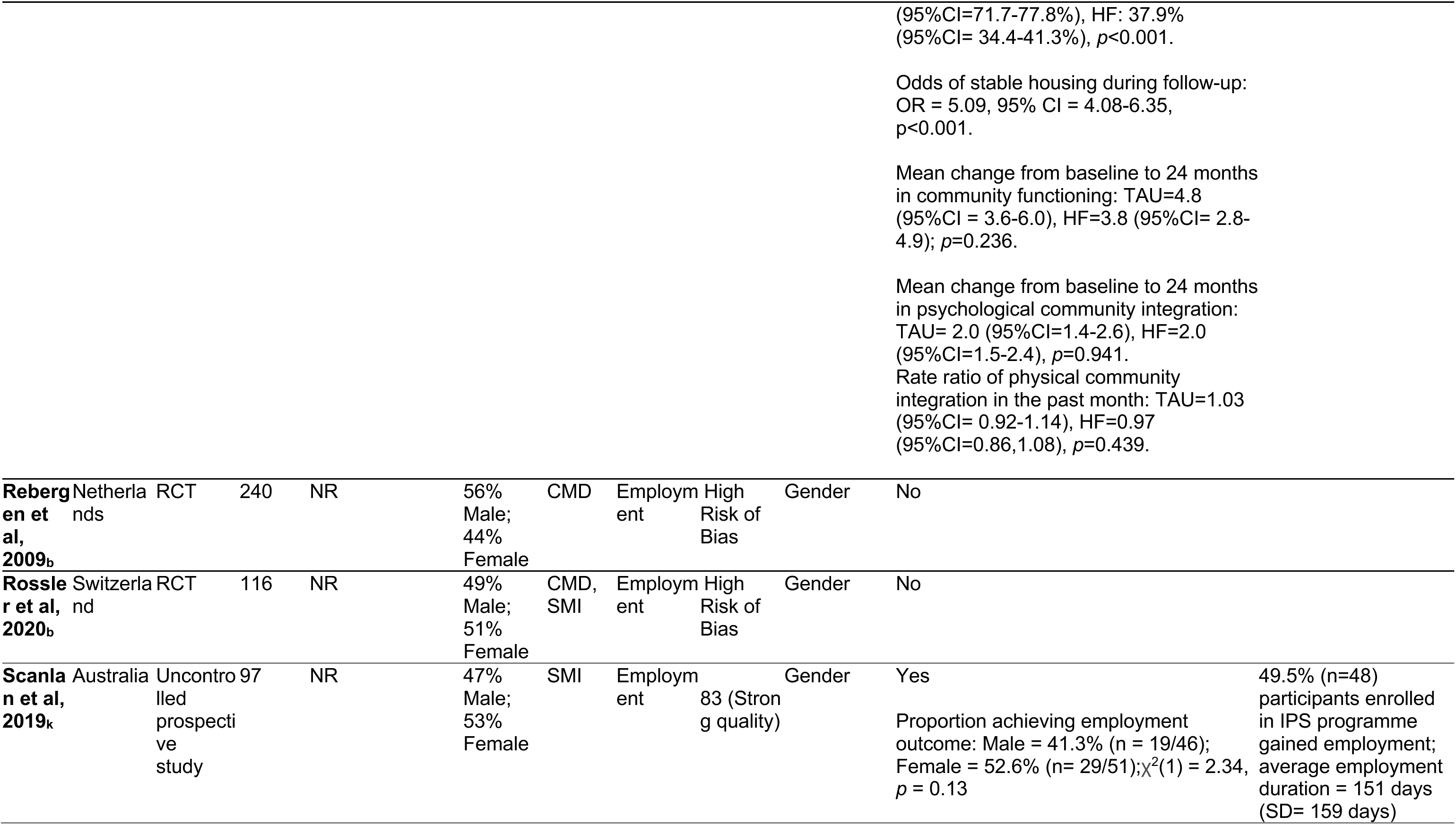
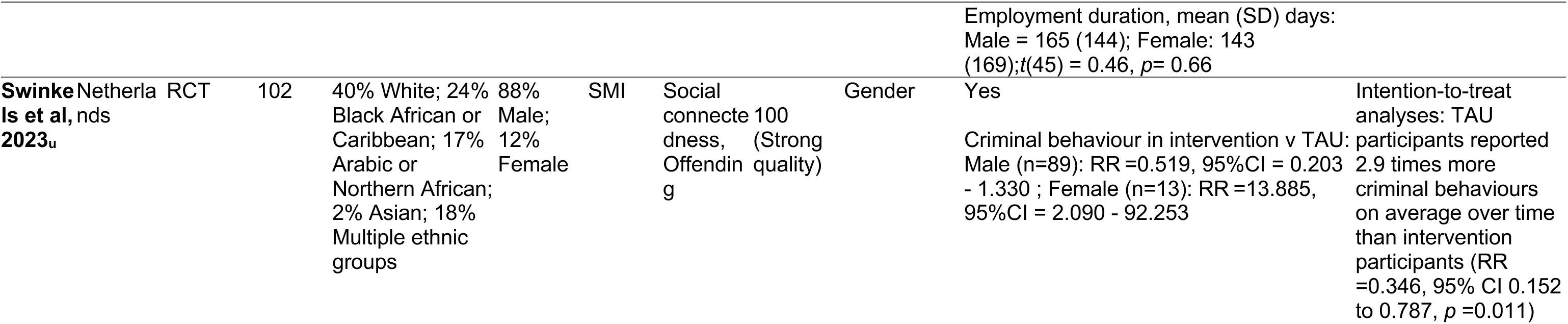
Characteristics and results of studies conducting stratified analyses on the basis of gender, ethnicity/race/Indigeneity, and socio-economic indicators. Each study was classified as comprising “Sufficient data” where they included effect size and associated data (e.g. mean scores by group) relevant to at least one stratified analyses on the basis of each indicator. *= studies exclusively including participants with psychosis spectrum diagnoses. u= identified in updated search; k = identified in Killaspy et al (2022); b = identified in Barnett et al., (2022). QA kmet scores reported for studies included from the updated search and Killaspy et al., whereas Cochrane Risk of Bias scores are reported for studies included from Barnett et al., 2022, obtained via communication with the authors. See SI Table 6 for further details on stratified analyses.

## DISCUSSION

Our review shows that research on interventions seeking to improve social and economic circumstances of people with mental ill-health gives little consideration to the social contexts in which interventions are tested and the social groups included. When relevant information is included, variations in reporting and data missingness obstructs a complete understanding of inclusion of marginalised groups. There is similarly a lack of data regarding people who experience multiple intersecting forms of marginalisation. In the rare instances where analyses are stratified by social group, some evidence suggests that people from marginalised socioeconomic positions may benefit least. Overall, we are very far from understanding which interventions work for those who need them most.

There are some limitations that should be considered when interpreting our findings. Due to the slight discrepancy in search strategies between the two reviews we updated, coverage of the literature was less thorough for the period covered by the two previous reviews compared with the updated review period. For example, non-RCTs recruiting people with common mental disorder diagnoses would have been missed by the two previous reviews as Barnett et al (2022) only included RCTs and Killaspy et al (2022) only included studies of severe mental illnesses. Further, as we screened for samples with a diagnosed mental disorder or who had accessed mental health services, we may have missed social care studies not based on diagnostic frameworks. Related to this, as we restricted our search to articles published in peer-reviewed journals, we overlooked social interventions evaluated in the grey literature. Additionally, we only included English language papers and may have missed studies conducted in non-English-speaking countries.

### Social intervention research domains, contexts and participants

Most research we identified was conducted in the Global North, with most in the USA, and in urban settings. We observed a marked increase in research on interventions to improve social and economic circumstances since approximately 2010; however, this has clustered across a few specific domains. Most interventions focused on housing, employment, social connectedness and community participation. There are areas of social and economic need that are largely neglected, notably interventions addressing debt, finances, and victimisation. We did not identify any studies testing the impact of primary prevention interventions, such as Universal Basic Income, which would also address the socioeconomic needs of those with mental ill-health. More work is needed given that debt, poverty and need for social security are very high for people with mental ill-health and can prevent recovery. Equally, mental ill-health can impact a person’s ability to manage finances and navigate complex social security systems (50–52).

Data on ethnicity and associated concepts were often poorly or incompletely reported. Inadequate reporting sometimes stemmed from national policy and legal frameworks. Estimating pooled statistics for representation was beyond the scope of this review. Nevertheless, our data highlights that intervention research may not be generalisable to contexts where the service users are not of majority White ethnicity: for example, White participants comprised the majority (70.85%) reported in UK samples, and as such may not apply to many services and contexts in the UK such as London where populations are more ethnically diverse. Socioeconomic status and sex/gender were more consistently reported; however, only 4% of studies reported any data on non-binary genders.

Studies were also unrepresentative of people with co-occurring conditions, substance use and physical ill-health. Approximately 1 in 5 studies excluded participants with substance abuse or dependence problems, which limits our ability to draw conclusions about intervention effectiveness for some of the most vulnerable in society. For studies including such populations, average comorbidity with a substance use disorder was high (55%). Even fewer studies reported the prevalence of physical health conditions in their samples. This is problematic given interactions between such factors and mental ill-health, which worsen outcomes (53–55). Further research is also required to understand the extent to which social intervention research has included people with neurodevelopmental conditions, such as autism, who are at high risk of developing mental ill-health (56), yet are marginalised within mental health care systems (57,58) and experience heightened social exclusion (59).

We found current reporting standards in social intervention research prevent a nuanced understanding of how forms of marginalisation may impact outcomes. Most papers do not report sociodemographic characteristics, and this precludes any consideration of variation by, and at the intersection of, marginalised groups. This is problematic given that mental health outcomes are worse for people in multiply marginalised groups (60). This reflects the dominant conceptualisation of social adversities in mental health research, whereby problems are located within the individual, as distinct risk factors (e.g. socioeconomic status, ethnicity) without consideration of how such factors may interact, or of the processes and social structures contributing to poor outcomes (61).

### What works for whom?

Our synthesis shows that we do not know what social interventions work best for whom. Under 10% of studies investigated the effectiveness of interventions by socioeconomic and sociodemographic groups. No studies or analyses were replicated.

Only 45% (n=13) of the stratified analyses reported adequate data for us to interpret their findings. Most inferred effectiveness based on statistical significance testing against arbitrary *p*-values (i.e. *p*≤.05), and did not report any data for analyses which were classed as “non-significant”. This approach is problematic (62–65), not least as *p* values are largely influenced by sample size. This is relevant here as many included studies were likely underpowered – as shown by our results regarding underrepresentation of marginalised groups – to detect differences between groups. Indeed, primary studies are often only powered to detect effects across all included participants and may struggle to recruit sufficient samples to investigate associations in subgroups. One way to address this is to foster open science initiatives whereby anonymised trial datasets are made available to other researchers to harmonise with other datasets and test outcomes for specific groups across multiple studies. Equally, improved attempts to pre-specify *a priori* subgroups for subgroup analyses are needed, particularly given the persistent problem of trials reporting unplanned subgroup analyses after finding statistically significant effects (66).

Within this context, some evidence suggested that effectiveness of social interventions may be lower for people in lower socioeconomic groups. Most RCT studies reporting sufficient data (4 out of 5 RCT studies) found this, including interventions to prevent victimisation (45), assist with budgeting skills (46), provide vocational peer support (47), and reduce caregiver burden (67). Further research is needed to firmly draw conclusions regarding socioeconomic position due to the limited number of studies.

The paucity of stratified analyses was most striking with respect to ethnicity. This is particularly problematic given the high inequalities in mental healthcare access and outcomes for minoritised ethnic groups (68–70). Further, these studies were constrained to the domains of housing and social skills, but we know people with mental health problems from minoritised ethnic groups experience heightened adversity across other domains, including social isolation (17,18) and unemployment (71), among others.

## CONCLUSIONS

In sum, we do not know whether the effectiveness of existing social interventions varies for different groups, although there was some indication that interventions risk reproducing existing inequalities due to lower effectiveness for those from more disadvantaged socioeconomic groups. The lack of stratified analyses prevents us from assessing whether findings from intervention research are translatable to local populations in practice. More nuanced research trials, open science efforts, and more representative recruitment practices are required.

## Data Availability

Data produced in the present study are available upon reasonable request to the authors

## ACKNOWLEDGEMENTS

This paper forms part of the ENRICHED project on social inclusion and mental health. We would like to thank Professor Helen Killaspy and Dr Phoebe Barnett for their support and advice in conducting this research, and, along with their co-authors, for conducting the reviews upon which this research was based. We would also like to thank Professor Claire Henderson, Katie Chamberlain and Madison Wempe for their contributions to this research.

## AUTHOR APPROVAL

All authors have seen and approved the manuscript

## SUPPLEMENTARY DATA

Supplementary information is available online at medRxiv

## References

1. Olesen SC, Butterworth P, Leach LS, Kelaher M, Pirkis J. Mental health affects future employment as job loss affects mental health: findings from a longitudinal population study. BMC Psychiatry. 2013 Dec;13(1):144.

2. Rinaldi M, Killackey E, Smith J, Shepherd G, Singh SP, Craig T. First episode psychosis and employment: A review. Int Rev Psychiatry. 2010 Jan;22(2):148– 62.

3. Harvey C, Killackey E, Groves A, Herrman H. A place to live: Housing needs for people with psychotic disorders identified in the second Australian national survey of psychosis. Aust N Z J Psychiatry. 2012 Sep;46(9):840–50.

4. Jones AA, Gicas KM, Seyedin S, Willi TS, Leonova O, Vila-Rodriguez F, et al. Associations of substance use, psychosis, and mortality among people living in precarious housing or homelessness: A longitudinal, community-based study in Vancouver, Canada. Tsai AC, editor. PLOS Med. 2020 Jul 6;17(7):e1003172.

5. Pevalin DJ, Reeves A, Baker E, Bentley R. The impact of persistent poor housing conditions on mental health: A longitudinal population-based study. Prev Med. 2017 Dec;105:304–10.

6. Sareen J, Afifi TO, McMillan KA, Asmundson GJG. Relationship Between Household Income and Mental Disorders: Findings From a Population-Based Longitudinal Study. Arch Gen Psychiatry. 2011 Apr 1;68(4):419.

7. Topor A, Stefansson CG, Denhov A, Bülow P, Andersson G. Recovery and economy; salary and allowances: a 10-year follow-up of income for persons diagnosed with first-time psychosis. Soc Psychiatry Psychiatr Epidemiol. 2019 Aug;54(8):919–26.

8. Nuyen J, Tuithof M, De Graaf R, Van Dorsselaer S, Kleinjan M, Have MT. The bidirectional relationship between loneliness and common mental disorders in adults: findings from a longitudinal population-based cohort study. Soc Psychiatry Psychiatr Epidemiol. 2020 Oct;55(10):1297–310.

9. Stain HJ, Galletly CA, Clark S, Wilson J, Killen EA, Anthes L, et al. Understanding the social costs of psychosis: The experience of adults affected by psychosis identified within the second Australian national survey of psychosis. Aust N Z J Psychiatry. 2012 Sep;46(9):879–89.

10. Honings S, Drukker M, Ten Have M, de Graaf R, van Dorsselaer S, van Os J. The interplay of psychosis and victimisation across the life course: a prospective study in the general population. Soc Psychiatry Psychiatr Epidemiol. 2017 Nov;52(11):1363–74.

11. Morgan C, Gayer-Anderson C. Childhood adversities and psychosis: evidence, challenges, implications. World Psychiatry. 2016 Jun;15(2):93–102.

12. Varese F, Smeets F, Drukker M, Lieverse R, Lataster T, Viechtbauer W, et al. Childhood Adversities Increase the Risk of Psychosis: A Meta-analysis of Patient-Control, Prospective- and Cross-sectional Cohort Studies. Schizophr Bull. 2012 Jun 18;38(4):661–71.

13. Phillips DM, Finkel D, Petkus AJ, Muñoz E, Pahlen S, Johnson W, et al. Longitudinal analyses indicate bidirectional associations between loneliness and health. Aging Ment Health. 2023 Jun 3;27(6):1217–25.

14. Barnett P, Oshinowo I, Cooper C, Taylor C, Smith S, Pilling S. The association between social class and the impact of treatment for mental health problems: a systematic review and narrative synthesis. Soc Psychiatry Psychiatr Epidemiol. 2023 Apr;58(4):581–603.

15. Lambri M, Chakraborty A, Leavey G, King M. Quality of Life and Unmet Need in People with Psychosis in the London Borough of Haringey, UK. Sci World J. 2012;2012:1–10.

16. Chilman N, Laporte D, Dorrington S, Hatch SL, Morgan C, Okoroji C, et al. Understanding social and clinical associations with unemployment for people with schizophrenia and bipolar disorders: large-scale health records study. Soc Psychiatry Psychiatr Epidemiol [Internet]. 2024 Feb 20 [cited 2024 Jul 27]; Available from: 10.1007/s00127-024-02620-6

17. Morgan C, Kirkbride J, Hutchinson G, Craig T, Morgan K, Dazzan P, et al. Cumulative social disadvantage, ethnicity and first-episode psychosis: a case-control study. Psychol Med. 2008 Dec;38(12):1701–15.

18. Morgan C, Fearon P, Lappin J, Heslin M, Donoghue K, Lomas B, et al. Ethnicity and long-term course and outcome of psychotic disorders in a UK sample: the ÆSOP-10 study. Br J Psychiatry. 2017;211(2):88–94.

19. Das-Munshi J, Bhugra D, Crawford MJ. Ethnic minority inequalities in access to treatments for schizophrenia and schizoaffective disorders: findings from a nationally representative cross-sectional study. BMC Med. 2018 Apr 18;16(1):55.

20. Harwood H, Rhead R, Chui Z, Bakolis I, Connor L, Gazard B, et al. Variations by ethnicity in referral and treatment pathways for IAPT service users in South London. Psychol Med. 2023 Feb;53(3):1084–95.

21. Schlief M, Rich N, Rains LS, Baldwin H, Rojas-Garcia A, Nyikavaranda P, et al. Ethnic differences in receipt of psychological interventions in Early Intervention in Psychosis services in England – a cross-sectional study. Psychiatry Res. 2023 Dec;330:115529.

22. Boardman J, Killaspy H, Mezey G. Social Inclusion and Mental Health: Understanding Poverty, Inequality and Social Exclusion [Internet]. 2nd ed. Cambridge University Press; 2023 [cited 2024 Oct 8]. Available from: https://www.cambridge.org/core/product/identifier/9781911623601/type/book

23. Barnett P, Steare T, Dedat Z, Pilling S, McCrone P, Knapp M, et al. Interventions to improve social circumstances of people with mental health conditions: a rapid evidence synthesis. BMC Psychiatry. 2022 Dec;22(1):302.

24. Killaspy H, Harvey C, Brasier C, Brophy L, Ennals P, Fletcher J, et al. Community-based social interventions for people with severe mental illness: a systematic review and narrative synthesis of recent evidence. World Psychiatry. 2022 Feb;21(1):96–123.

25. Kmet, Leanne M. ; Cook, Linda S. ; Lee, Robert C. Standard Quality Assessment Criteria for Evaluating Primary Research Papers from a Variety of Fields [Internet]. University of Alberta Libraries; 2004 [cited 2024 Jun 3]. Available from: https://era.library.ualberta.ca/files/9s1619324

26. Higgins JPT, Altman DG, Gøtzsche PC, Jüni P, Moher D, Oxman AD, et al. The Cochrane Collaboration’s tool for assessing risk of bias in randomised trials. BMJ. 2011 Oct 18;343:d5928.

27. Appleton R, Barnett P, Chipp B, Clark M, Goldblatt P, Jeffreys S, et al. Development of a conceptual framework to guide description and evaluation of social interventions for people with serious mental health conditions. SSM Ment Health. 2023;4.

28. Bhopal R, Rafnsson S. Global inequalities in assessment of migrant and ethnic variations in health. Public Health. 2012 Mar 1;126(3):241–4.

29. Flanagin A, Frey T, Christiansen SL, AMA Manual of Style Committee. Updated Guidance on the Reporting of Race and Ethnicity in Medical and Science Journals. JAMA. 2021 Aug 17;326(7):621–7.

30. Lu C, Ahmed R, Lamri A, Anand SS. Use of race, ethnicity, and ancestry data in health research. PLOS Glob Public Health. 2022 Sep 15;2(9):e0001060.

31. Bhopal R, Gruer L, Agyemang C, Davidovitch N, de-Graft Aikins A, Krasnik A, et al. The Global Society on Migration, Ethnicity, Race and Health: why race can’t be ignored even if it causes discomfort. Eur J Public Health. 2021 Feb 1;31(1):3– 4.

32. Galobardes B, Shaw M, Lawlor DA, Lynch JW, Smith GD. Indicators of socioeconomic position (part 1). J Epidemiol Community Health. 2006 Jan 1;60(1):7–12.

33. Scanlan JN, Feder K, Ennals P, Hancock N. Outcomes of an individual placement and support programme incorporating principles of the collaborative recovery model. Aust Occup Ther J. 2019 Aug;66(4):519–29.

34. Dubreucq J, Gabayet F, Ycart B, Faraldo M, Melis F, Lucas T, et al. Improving social function with real-world social-cognitive remediation in schizophrenia: Results from the RemedRugby quasi-experimental trial. Eur Psychiatry. 2020;63(1):e41.

35. McHugo GJ, Bebout RR, Harris M, Cleghorn S, Herring G, Xie H. A randomized controlled trial of integrated versus parallel housing services for homeless adults with severe mental illness. Schizophr Bull. 2004;30(4):969–82.

36. Swinkels LTA, Van Der Pol TM, Twisk J, Ter Harmsel JF, Dekker JJM, Popma A. The effectiveness of an additive informal social network intervention for forensic psychiatric outpatients: results of a randomized controlled trial. Front Psychiatry. 2023 May 24;14:1129492.

37. O’Campo P, Nisenbaum R, Crocker AG, Nicholls T, Eiboff F, Adair CE. Women experiencing homelessness and mental illness in a Housing First multi-site trial: Looking beyond housing to social outcomes and well-being. Rosenbaum JE, editor. PLOS ONE. 2023 Feb 10;18(2):e0277074.

38. Christensen TN, Wallstrøm IG, Bojesen AB, Nordentoft M, Eplov LF. Predictors of work and education among people with severe mental illness who participated in the Danish individual placement and support study: findings from a randomized clinical trial. Soc Psychiatry Psychiatr Epidemiol. 2021 Sep;56(9):1669–77.

39. Martín-Carrasco M, Fernández-Catalina P, Domínguez-Panchón AI, Gonçalves-Pereira M, González-Fraile E, Muñoz-Hermoso P, et al. A randomized trial to assess the efficacy of a psychoeducational intervention on caregiver burden in schizophrenia. Eur Psychiatry. 2016;33(1):9–17.

40. Caplan RA, Nelson G, Distasio J, Isaak C, Edel B, Macnaughton E, et al. Parent– child relationship outcomes in a randomized controlled trial of housing first for indigenous and non-Indigenous parents experiencing homelessness, mental illness, and separation from their children. Psychiatr Rehabil J. 2023 Dec;46(4):335–42.

41. Goldfinger SM, Schutt RK, Tolomiczenko GS, Seidman L, Penk WE, Turner W, et al. Housing Placement and Subsequent Days Homeless Among Formerly Homeless Adults With Mental Illness. Psychiatr Serv. 1999 May;50(5):674–9.

42. Marder SR, Wirshing WC, Mintz J, McKenzie J, Johnston K, Eckman TA, et al. Two-year outcome of social skills training and group psychotherapy for outpatients with schizophrenia. Am J Psychiatry. 1996 Dec;153(12):1585–92.

43. Brown M, Jason L, D M. Housing first as an effective model for community stabilization among vulnerable individuals with chronic and nonchronic homelessness histories. J Community Psychol. 2016;44:384–90.

44. Gutman SA, Kerner R, Zombek I, Dulek J, Ramsey CA. Supported education for adults with psychiatric disabilities: effectiveness of an occupational therapy program. Am J Occup Ther. 2009;63(3):245–54.

45. De Waal MM, Dekker JJM, Kikkert MJ, Christ C, Chmielewska J, Staats MWM, et al. Self-wise, Other-wise, Streetwise (SOS) training, an intervention to prevent victimization in dual-diagnosis patients: results from a randomized clinical trial. Addiction. 2019 Apr;114(4):730–40.

46. Elbogen EB, Hamer RM, Swanson JW, Swartz MS. A Randomized Clinical Trial of a Money Management Intervention for Veterans With Psychiatric Disabilities. Psychiatr Serv Wash DC. 2016 Oct 1;67(10):1142–5.

47. Maru M, Rogers ES, Nicolellis D, Legere L, Placencio-Castro M, Magee C, et al. Vocational peer support for adults with psychiatric disabilities: Results of a randomized trial. Psychiatr Rehabil J. 2021 Dec;44(4):327–36.

48. Kirkbride JB, Anglin DM, Colman I, Dykxhoorn J, Jones PB, Patalay P, et al. The social determinants of mental health and disorder: evidence, prevention and recommendations. World Psychiatry. 2024 Feb;23(1):58–90.

49. Wilson N, McDaid S. The mental health effects of a Universal Basic Income: A synthesis of the evidence from previous pilots. Soc Sci Med. 2021 Oct;287:114374.

50. Baekgaard M, Nielsen SA, Rosholm M, Svarer M. Long-term employment and health effects of active labor market programs. Proc Natl Acad Sci. 2024 Dec 10;121(50):e2411439121.

51. Simpson J, Albani V, Bell Z, Bambra C, Brown H. Effects of social security policy reforms on mental health and inequalities: A systematic review of observational studies in high-income countries. Soc Sci Med 1982. 2021 Mar;272:113717.

52. Wickham S, Bentley L, Rose T, Whitehead M, Taylor-Robinson D, Barr B. Effects on mental health of a UK welfare reform, Universal Credit: a longitudinal controlled study. Lancet Public Health. 2020 Mar;5(3):e157–64.

53. Bhui K, Halvorsrud K, Mooney R, Hosang GM. Is psychosis a syndemic manifestation of historical and contemporary adversity? Findings from UK Biobank. Br J Psychiatry. 2021;219(6):686–94.

54. Coid J, Zhang Y, Bebbington P, Ullrich S, De Stavola B, Bhui K, et al. A syndemic of psychiatric morbidity, substance misuse, violence, and poor physical health among young Scottish men with reduced life expectancy. SSM - Popul Health. 2021 Sep;15:100858.

55. Mendenhall E, Kohrt BA, Norris SA, Ndetei D, Prabhakaran D. Non-communicable disease syndemics: poverty, depression, and diabetes among low-income populations. The Lancet. 2017 Mar 4;389(10072):951–63.

56. Lai MC, Kassee C, Besney R, Bonato S, Hull L, Mandy W, et al. Prevalence of co-occurring mental health diagnoses in the autism population: a systematic review and meta-analysis. Lancet Psychiatry. 2019 Oct 1;6(10):819–29.

57. Loizou S, Pemovska T, Stefanidou T, Foye U, Cooper R, Kular A, et al. Approaches to improving mental healthcare for autistic people: systematic review. BJPsych Open. 2024 Jul;10(4):e128.

58. Mandy W. Six ideas about how to address the autism mental health crisis. Autism. 2022 Feb 1;26(2):289–92.

59. Gray KM, Keating CM, Taffe JR, Brereton AV, Einfeld SL, Reardon TC, et al. Adult Outcomes in Autism: Community Inclusion and Living Skills. J Autism Dev Disord. 2014 Dec 1;44(12):3006–15.

60. Moreno-Agostino D, Woodhead C, Ploubidis GB, Das-Munshi J. A quantitative approach to the intersectional study of mental health inequalities during the COVID-19 pandemic in UK young adults. Soc Psychiatry Psychiatr Epidemiol. 2024 Mar;59(3):417–29.

61. Bemme D, Béhague D. Theorising the social in mental health research and action: a call for more inclusivity and accountability. Soc Psychiatry Psychiatr Epidemiol. 2024 Mar;59(3):403–8.

62. Amrhein V, Greenland S, McShane B. Scientists rise up against statistical significance. Nature. 2019 Mar;567(7748):305–7.

63. Greenland S. Connecting simple and precise P-values to complex and ambiguous realities (includes rejoinder to comments on “Divergence vs. decision P-values”). Scand J Stat. 2023;50(3):899–914.

64. Greenland S, Senn SJ, Rothman KJ, Carlin JB, Poole C, Goodman SN, et al. Statistical tests, P values, confidence intervals, and power: a guide to misinterpretations. Eur J Epidemiol. 2016 Apr 1;31(4):337–50.

65. Savitz DA, Wise LA, Bond JC, Hatch EE, Ncube CN, Wesselink AK, et al. Responding to Reviewers and Editors About Statistical Significance Testing. Ann Intern Med. 2024 Mar 19;177(3):385–6.

66. Kasenda B, Schandelmaier S, Sun X, Elm E von, You J, Blümle A, et al. Subgroup analyses in randomised controlled trials: cohort study on trial protocols and journal publications. BMJ. 2014 Jul 16;349:g4539.

67. Martin-Carrasco M, Fernandez-Catalina P, A DP. A randomized trial to assess the efficacy of a psychoeducational intervention on caregiver burden in schizophrenia. Eur Psychiatry. 2016;33:9–17.

68. Bansal N, Karlsen S, Sashidharan SP, Cohen R, Chew-Graham CA, Malpass A. Understanding ethnic inequalities in mental healthcare in the UK: A meta-ethnography. PLOS Med. 2022 Dec 13;19(12):e1004139.

69. Barnett P, Mackay E, Matthews H, Gate R, Greenwood H, Ariyo K, et al. Ethnic variations in compulsory detention under the Mental Health Act: a systematic review and meta-analysis of international data. Lancet Psychiatry. 2019 Apr;6(4):305–17.

70. Chorlton E, McKenzie K, Morgan C, Doody G. Course and outcome of psychosis in black Caribbean populations and other ethnic groups living in the UK: A systematic review. Int J Soc Psychiatry. 2012 Jul;58(4):400–8.

71. Henderson M. Race Inequality in the Workforce: Exploring Connections between Work, Ethnicity and Mental Health [Internet]. Carnegie UK; Carnegie UK Trust, UCL Centre for Longitudinal Studies; Operation Black Vote: London, UK. London, UK: Carnegie UK; Carnegie UK Trust, UCL Centre for Longitudinal Studies; Operation Black Vote; 2020 Feb [cited 2024 Jul 29]. Available from: https://www.carnegieuktrust.org.uk/publications/race-inequality-in-the-workforce/

