## Supplementary_Material for "What works for whom: a systematic review of inequalities in inclusion and effectiveness of social interventions for mental ill-health"

### Supplementary Information

#### Database searches – further details

Database searches, data extraction and synthesis were conducted according to a pre-defined protocol. Database records were imported, de-duplicated, and screened on Rayyan software according to a set of inclusion criteria (see Supplementary Materials) based upon original criteria set out in the two reviews (Barnett et al., 2022; Killaspy et al., 2022). Where there were minor discrepancies between the two sets of inclusion criteria for each review, we adopted the more broad or inclusive criteria, with the exception of the current exclusion of any secondary research, such as meta-analyses and systematic reviews. The Killaspy et al (2022) searches were replicated in MEDLINE via Ovid, and the Barnett et al (2022) searches (see below) were replicated in Web of Science (SciELO database), PsycINFO via Ovid and the Cochrane Central Register of Controlled Trials (CENTRAL) to identify literature published between January 2020 and February 2024.

#### Table 1. Inclusion and exclusion criteria for Killaspy et al (2022) and Barnett et al., (2022)

| | Barnett | Killaspy |
|---|---|---|
| Study design and details | • Systematic reviews and RCTs<br>• RCTs included feasibility and pilot trials<br>• English language<br>• High income countries<br>• RCTs: 2000-aug 2020<br>• SRs: inception-feb 2020 | • Peer-reviewed papers reporting primary empirical data<br>• Published between Jan 2016-July 2020<br>• English<br>• Full text availability |
| | Exclusions: | Exclusions:<br>• Publications that did not report primary empirical data<br>• Reviews, editorials and commentaries |
| Participants | • Adults 18+<br>• Any mental health condition or diagnosis of personality disorder established through clinical diagnosis, meeting threshold criteria on an established diagnostic screening tool or symptom severity measure; or users of specialist MH services (minimum 80% sample) | • Adults ages 18-65 years with severe and persistent mental illness<br>• People with SMI, defined as a primary diagnosis of schizophrenia, schizoaffective disorder, bipolar disorder, or other severe and enduring psychotic disorder. |
| | Exclusions:<br>Intellectual/learning disability, dementia or other organic mental disorder, neurodevelopmental disorder or acquired cognitive impairment, anti-social personality disorder, adjustment disorder, substance use disorder (in the absence of any mental illness or personality disorder) | Exclusions:<br>Studies that focused on individuals with a primary diagnosis of personality, depressive or anxiety disorder, substance use disorder, acquired brain injury, intellectual disability, or trauma due to natural disasters or military service; studies where fewer than 50% of the sample met our SMI diagnostic inclusion criteria |
| Intervention | • Non-pharmacological interventions designed to improve social circumstances in any of the included life domains where this was the primary outcome or otherwise described in the paper as an explicit, direct focus of the intervention.<br>• Interventions designed to improve more than one life domain, e.g. through helping people access available services, groups or community resources were also included where improving overall social circumstances was the primary aim of the programme.<br>• Social inclusion domains: housing/homelessness; money | • Models of care or interventions (group or individual) that are carried out in the community<br>• Interventions that aimed to improve social inclusion<br>• Social inclusion domains: supported accommodation, supported education, supported employment, community participation interventions, family interventions, peer-supported/developed/led interventions; social skills training interventions)<br>• Models of care or interventions that also comprised a peer component were included within the relevant category. The separate peer-led/supported interventions category included studies where the peer component was not delivered as part of |

| | | |
|---|---|---|
| | and basic needs; work and education; social isolation and connectedness; family, intimate and caring relationships; victimisation and exploitation; offending; rights, inclusion and citizenship [see outcomes for full list of relevant social circumstances to each domain]<br>• For social isolation and family/relationship domains: we limited included outcomes for individual relationships to the maintenance or gain of social roles (e.g. retention of partner relationship, parental contact, carer role) | one of the other included models of care or interventions. |
| | Exclusions: Interventions focussing on subjective measures of social isolation/family relationships | Exclusions: studies conducted in environments other than the community, for example inpatient units or prisons |
| Comparator | Comparators of routine care, no support or an active intervention were all included. | Any/none |
| Outcomes | Studies needed to report at least one outcome specifically relating to the social circumstances:<br><br>Housing: homelessness; housing instability; housing quality<br><br>Money/basic needs: poverty/income: financial barriers; debt; money management<br><br>Work/education: unemployment; achieving and sustaining paid employment; precarious work; lack of access/completion of educational goals; lack of meaningful activity; length of illness absence<br><br>Social isolation/connectedness: subjective social isolation/loneliness; objective SI and social network; social capital<br><br>Family: partner/sexual relationships; maintaining parenting roles/ contact with children; maintaining contact/cohabitation with family members; caring responsibilities<br><br>Victimisation: victim of crime; sexual or physical assault; domestic | • Any indicator of improved social or economic participation. For example -<br>• Supported accommodation: housing stability or progression to more independent accommodation;<br>• Supported employment or supported education: outcomes related to gaining or sustaining employment in a competitive, paid or unpaid post, or engagement in mainstream or supported study or volunteering.<br>• family interventions: outcomes relating to family functioning such as expressed emotion and carer burden (not measured at the individual level)<br>• For other interventions, outcomes included measures of social skills, social functioning, engagement in community-based activities, social connection, self-efficacy, hope and empowerment. |

| | | |
|---|---|---|
| | violence/coercive control; exploitation, harassment, safeguarding concerns<br><br>Offending: risk of offending; transition from prison to community; reoffending<br><br>Rights/inclusion/citizenship: social exclusion and participation; access to public services; immigration status; privacy/dignity resulting from social circumstances | |
| | Exclusions:<br>In social isolation and family, intimate and caring relationships domains, excluded subjective social; relationship outcomes: i) individual perceived relationship quality including parent–child attachment and partner relationship or parenting quality, ii) family relationship quality including expressed emotion, and iii) experienced or self-stigma. | Exclusions: studies that did not report on any relevant social outcomes |

#### Table 2. Full inclusion and exclusion criteria for the updated search

These criteria are taken from Barnett et al., (2022) and Killaspy et al., (2022). Where there were minor discrepancies between the two sets of inclusion criteria for each review, we adopted the more broad or inclusive criteria, with the exception of the current exclusion of any secondary research, such as meta-analyses and systematic reviews

| | **Inclusion criteria** | **Exclusion criteria** |
|---|---|---|
| **Study design and details** | • Peer-reviewed papers reporting primary empirical data (including RCTs, feasibility and pilot trials, qualitative and mixed-method evaluations)<br>• Published between July 2020 and February 2024<br>• English language<br>• Full-text availability | • Publications that did not report primary empirical data (but relevant reviews will be labelled for reference searching)<br>• Editorials and commentaries<br>• Protocols (but relevant protocols will be labelled to identify associated publications) |
| **Participants** | • Adults 18+<br>• Any mental health condition or diagnosis of personality disorder established through clinical diagnosis, meeting threshold criteria on an established diagnostic screening tool or symptom severity measure; or users of specialist MH services (minimum 80% sample) | • Children and adolescents under the age of 18<br>• Intellectual/learning disability, dementia or other organic mental disorder, neurodevelopmental disorder or acquired cognitive impairment, anti-social personality disorder, adjustment |

| | | | |
|---|---|---|---|
| | | | disorder, substance use disorder (in the absence of any mental illness or personality disorder) |
| **Intervention** | • Non-pharmacological interventions **designed** to improve social circumstances in any of the included life domains **where this was the primary outcome or otherwise described in the paper as an explicit, direct focus of the intervention.**<br>• Interventions designed to improve more than one life domain, e.g. through helping people access available services, groups or community resources were also included where improving overall social circumstances was the primary aim of the programme.<br>• Social inclusion domains: housing/homelessness; money and basic needs; work and education; social isolation and connectedness; family, intimate and caring relationships; victimisation and exploitation; offending; rights, inclusion and citizenship<br>• Models of care or interventions that also comprised a peer component are to be included within the relevant category. | | |
| **Comparator** | • Any comparators can be included (or no comparator at all) | | |
| **Outcomes** | Studies need to report at least one outcome specifically relating to the social/economic circumstances:<br>• Housing: homelessness; housing instability; housing quality<br>• Money/basic needs: poverty/income: financial barriers; debt; money management<br>• Work/education: unemployment; achieving and sustaining paid employment; precarious work; lack of access/completion of educational goals; lack of meaningful activity; length of illness absence<br>• Social isolation/connectedness: subjective social isolation/loneliness; objective SI and social network; social capital<br>• Social skills, social functioning, engagement in community-based activities, social connection, self-efficacy, hope and empowerment.<br>• Family: partner/sexual relationships; maintaining parenting roles/ contact with children; maintaining contact/cohabitation with family members; caring responsibilities; outcomes relating to family functioning such as expressed emotion and carer burden<br>• Victimisation: victim of crime; sexual or physical assault; domestic violence/coercive | | • Instances where the only social outcomes included are those only measured through social cognition tasks e.g. emotion recognition tasks |

| | |
|---|---|
| | control; exploitation, harassment, safeguarding concerns<br>• Offending: risk of offending; transition from prison to community; reoffending<br>• Rights/ inclusion /citizenship: social exclusion and participation; access to public services; immigration status; privacy/dignity resulting from social circumstances<br>• Or fidelity and acceptability assessments of interventions which directly aim to improve the social or economic circumstances of people living with a mental health condition. |

#### Figure 1. A PRISMA Diagram demonstrating the flow of records throughout the review process.

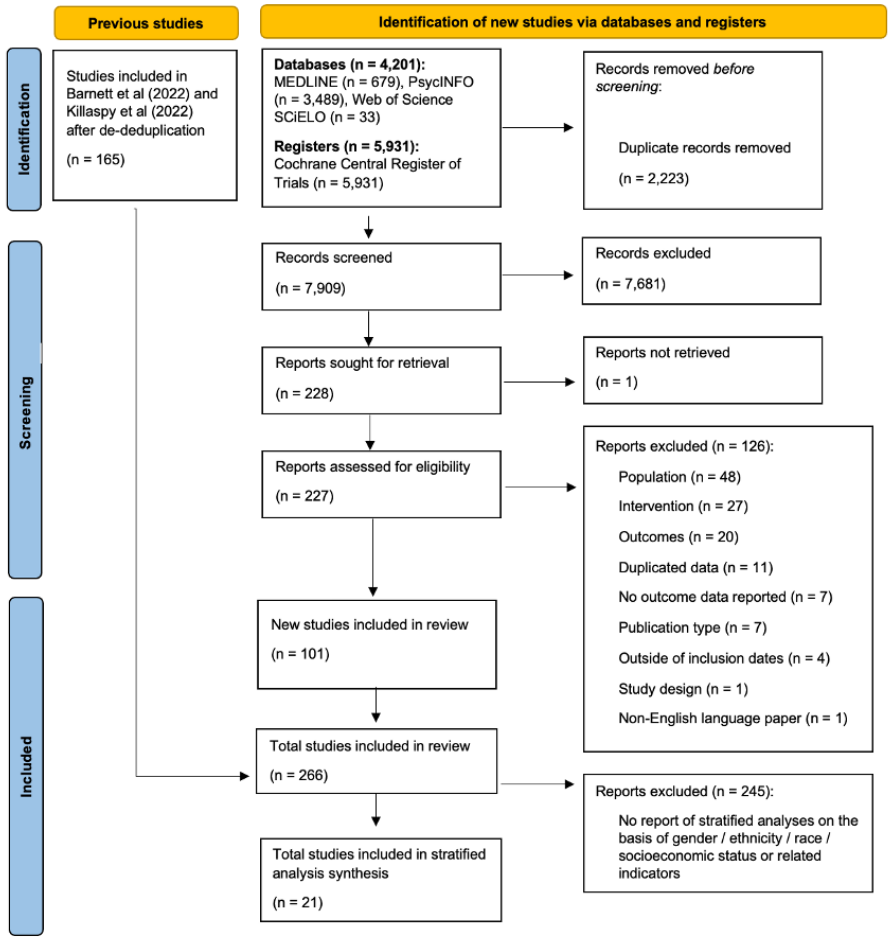

#### Data extraction - further details

Data were extracted using a fit-for-purpose data extraction sheet according to: article information (authors; date; source [Barnett review, Killaspy review, Updated searches]; study design; country in which the research was conducted; whether the research took place in an urban, rural or mixed environment; and whether there was any lived experience in the design or conduct of the research), characteristics of the sample (sample size; diagnostic profile of the sample; whether the sample comprised only people with psychosis; proportion of the sample with substance abuse problems noted; proportion of the sample with physical health morbidities noted; ethnicity (and related data including race, country of birth, nationality, indigeneity); gender; age; indicators of participant socioeconomic status; refugee status), the intervention tested (intervention name; intervention domain; comparator conditions, if applicable; setting in which the intervention was tested), any applicable outcomes (social and/or economic participation outcome measures; clinical outcome measures; any other outcome measures; effectiveness results in social or economic inclusion domains), and whether analyses pertaining to social and/or economic inclusion outcome domains were stratified by gender, ethnicity or socioeconomic status. We note that data relating to social and/or participation outcomes were only extracted where social and/or economic participation measures could be explicitly separated (i.e. in a subscale), rather than when these factors formed part of a composite score with non-social or economic outcomes.

Data extractions were conducted by one researcher and all data extractions were checked by a second member of the team. Where it was clear that the study sample was identical across more than one paper, we only extracted this data once, to avoid double counting. Ethnicity data was extracted twice and checked for coherence (HB, AG). In practice, effectiveness data was double extracted during the results synthesis process. Any conflicts were resolved through team discussion.

#### Quality appraisal results – further details

For all studies which were rated using Kmet tool (ie studies derived from the updated search and the Killaspy et al., 2022 review), Kmet quality appraisal scores ranged from 69%- 100% for quantitative studies, and 65%-95% for qualitative studies. Overall, for papers included from the updated search, 96 studies were regarded as strong (score of >80%), eight as good (70-80%), three as adequate (50-70%), and none as limited (<50%), following precedent in previous research (Giusti et al., 2020; Lee et al., 2008). Inter-rater agreement on a random selection of 10% of double-appraised papers was 81% (quantitative) and 95% (qualitative), respectively. Mixed methods papers were appraised twice, against both quantitative and qualitative criteria. Discrepancies were resolved through discussion with the wider review team.

#### Table 3. A summary the key characteristics of the included studies within the systematic review

| Author, date | Country | Study design | Sample size | Ethnicity/Race/ Nationality/minority status (%) | Gender (%) | Diagnoses (SMI, CMD) | Intervention life domain | Quality Score |
|---|---|---|---|---|---|---|---|---|
| **Adamus et al, 2022**[u] | Switzerland | RCT/ Observational cohort study | 141 | 74% Swiss national; 26% Foreign national | 53% Male; 47% Female | SMI | Housing | 92 |
| **Agrest et al, 2019**[b] | Chile | Qualitative interviews | 15 | NR | 67% Male; 33% Female | SMI | Social connectedness | See Barnett et al, 2022 |
| **Agudelo-Hernandez et al, 2024**[u] | Colombia | Pre-post quasi-experimental | 131 | 94% stated that they did not belong to any specific ethnic group; 6% identified as belonging to the Embera people. | 40% Male; 60% Female | SMI, CMD | Community, Social connectedness | 77 |
| **Aivalioti et al, 2023**[u] | Greece | RCT | 30 | NR | 60% Male; 40% Female | SMI | Family | 77 |
| **Albers et al, 2021**[u] | Netherlands | Cluster RCT | 400 | NR | 61% Male; 39% Female | SMI | Trauma | 100 |
| **Al-HadiHasan et al, 2017**[k] | Jordan | Qualitative process evaluation | 17 | NR | 35% Male; 65% Female | SMI | Education | Quality score reported in Killaspy et al, 2022 |
| **Ali et al, 2021**[u] | UK | Pilot RCT | 16 | 50% White; 50% Other | 44% Male; 56% Female | SMI, CMD | Social connectedness | 92 |
| **Aubry et al, 2016**[b] | Canada | RCT | 950 | 21% Member of a racial or ethnic minority group; 19% Aboriginal | 68% Male; 32% Female | SMI | Housing | See Barnett et al, 2022 |
| **Aubry et al, 2019**[k] | Canada | RCT | 201 | 4% Indigenous; 81% White; 15% Other | 65% Male; 35% Female; >1% Transgender | SMI | Housing | Quality score reported in Killaspy et al, 2022 |

| | | | | | | | | |
|---|---|---|---|---|---|---|---|---|
| **Baller et al, 2020**[u] | USA | RCT (follow-up using administrative data) | 2,160 | 62% White, 27% Black, 11% Other | 47% Male; 53% Female | SMI, CMD | Employment | 100 |
| **Battle et al, 2023**[u] | USA | Open pilot trial | 32 (16 couples) | 88% White Non-Hispanic; 9% Latina or Hispanic; 3% Missing | 100% Female (and 100% Male partners) | CMD | Family | 86 |
| **Beaven et al, 2017**[k] | Australia | Survey | 29 | NR | 43% Male; 57% Female | SMI | Community | Quality score reported in Killaspy et al, 2022 |
| **Bejerholm et al, 2017**[b] | Sweden | RCT | 63 | 92% Native; 8% Immigrant | 28% Male; 72% Female | CMD, SMI | Employment | See Barnett et al, 2022 |
| **Bell et al, 1993**[b] | USA | RCT | 100 | 76% White; 21% Black; 3% Hispanic | 94% Male; 6% Female | SMI | Employment | See Barnett et al, 2022 |
| **Bell et al, 2003**[b] | USA | RCT | 63 | 62% Caucasian; 30% African American; 8% Hispanic; 0% Asian | 100% Male | SMI | Employment | See Barnett et al, 2022 |
| **Bell et al, 2005**[b] | USA | RCT | 145 | 63% Caucasian; 32% African American; 3% Hispanic; 3% Asian | 80% Male; 20% Female | SMI | Employment | See Barnett et al, 2022 |
| **Bell et al, 2008**[b] | USA | RCT | 77 | 47% Caucasian; 23% African American; 4% Hispanic; 1% Asian | 54% Male; 46% Female | SMI | Employment | See Barnett et al, 2022 |
| **Bell et al, 2018**[b] | USA | RCT | 77 | 94% Non-Hispanic/ Latino; 4% Hispanic/ Latino; 2% Unknown ethnicity; 3% American or Alaskan Native; 47% Black or African American; 44% White; 7% Unknown race | 90% Male; 10% Female | SMI | Employment | See Barnett et al, 2022 |
| **Beutel et al, 2005**[b] | Germany | RCT | 329 | NR | 42% Male; 58% Female | CMD | Employment | See Barnett et al, 2022 |
| **Bitter et al, 2017**[k] | Netherlands | RCT | 263 | 85% Born in the Netherlands; 15% Other | 65% Male; 35% Female | SMI | Social connectedness | Quality score reported in |

| | | | | | | | | |
|---|---|---|---|---|---|---|---|---|
| | | | | | | | | Killaspy et al, 2022 |
| **Bjorkedal et al, 2023**[u] | Denmark | RCT | 139 | NR | 49% Male; 51% Female | CMD, SMI | Community | 100 |
| **Blajeski et al, 2023*a***[u] | USA | Mixed-methods evaluation of an RCT | 59 | 64% African American Black; 24% White; 7% Latinx; 5% More than one race | 46% Male; 54% Female | CMD, SMI | Employment | 73 |
| **Blajeski et al, 2023*b***[u] | USA | Qualitative evaluation of an RCT | 16 recipients and 22 staff | Recipients: 50% Black; 38% White; 0% Asian; 0% Latinx; 13% More than one race | NR | SMI | Employment | 90 |
| **Boevink et al, 2016**[b] | Netherlands | RCT | 163 | NR | 51% Male; 49% Female | SMI | Social connectedness | See Barnett et al, 2022 |
| **Brown et al, 2016**[k] | USA | Retrospective pre-post analysis | 182 | 56% White; 25% Black; 7% Asian/ Pacific Islander; 13% Native American, Latino/a or multiethnic | 74% Male; 26% Female | SMI | Housing | Quality score reported in Killaspy et al, 2022 |
| **Burnam et al, 1996**[b] | USA | RCT | 276 | 58% White; 28% Black; 14% Other | 84% Male; 16% Female | SMI | Housing | See Barnett et al, 2022 |
| **Burns et al, 2023**[u] | Canada | RCT | 97 | 71% White; 21% Asian; 4% Black; 4% Other, including Hispanic | 57% Male; 43% Female | SMI | Employment | 100 |
| **Caplan et al, 2023**[u] | Canada | RCT | 43 parent-child dyads | NR | 63% Male; 37% Female | CMD, SMI | Housing | 85 |
| **Castelein et al, 2008**[b] | Netherlands | RCT | 106 | NR | 66% Male; 34% Female | SMI | Community | See Barnett et al, 2022 |
| **Cella et al, 2023**[u] | UK | Proof-of-concept trial | 48 | NR | 56% Male; 44% Female | SMI | Social connectedness | 92 |
| **Cervello et al, 2021**[u] | France | RCT | 53 | NR | 17% Male; 83% Female | SMI | Employment | 75 |

| | | | | | | | | |
|---|---|---|---|---|---|---|---|---|
| **Chandler et al, 2006[b]** | USA | RCT | 182 | 66% African American; 21% White; 3% Hispanic; 3% Other | 80% Male; 20% Female | SMI | Offending | See Barnett et al, 2022 |
| **Chaudhry et al, 2023[u]** | Pakistan | Feasibility RCT | 26 | NR | 100% Female | CMD | Family | 100 (quant) 90 (qual) |
| **Chen et al, 2020[k]** | China | Pilot RCT | 51 | 100% Native Chinese | 77% Male; 33% Female | SMI | Community | Quality score reported in Killaspy et al, 2022 |
| **Christensen et al, 2019[b]** | Denmark | RCT | 477 | NR | 62% Male; 38% Female | SMI | Employment | See Barnett et al, 2022 |
| **Christensen et al, 2021[u]** | Denmark | RCT | 720 | NR | 62% Male; 38% Female | SMI | Employment | 96 |
| **Compton et al, 2023[u]** | USA | RCT | 240 | 48% Black African American; 48% White Caucasian; 4% Other | 65% Male; 35% Female | SMI | Community | 92 |
| **Conoley et al, 1985[b]** | USA | RCT | 38 | NR | 100% Female | CMD | Social connectedness | See Barnett et al, 2022 |
| **Cook et al, 2008[b]** | USA | RCT | 1273 | 50% White; 50% Other | 47% Male; 53% Female | SMI | Employment | See Barnett et al, 2022 |
| **Cook et al, 2016[k]** | USA | Controlled trial | 449 | 52% White non-Hispanic; 30% Black; 18% Hispanic or Latino | 52% Male; 48% Female | SMI | Employment | Quality score reported in Killaspy et al, 2022 |
| **Cosden et al, 2005[b]** | USA | RCT | 235 | 71% European American; 17% Hispanic; 8% African American; 4% Other | 49% Male; 51% Female | SMI | Offending | See Barnett et al, 2022 |
| **Creech et al, 2022[u]** | USA | Pilot trial | 21 | 57% White; 10% Black; 34% Multiracial or Other | 43% Male; 57% Female | SMI | Family | 86 |

| | | | | | | | | |
|---|---|---|---|---|---|---|---|---|
| **Cusack et al, 2010**[b] | USA | RCT | 134 | 63% Caucasian; 22% Hispanic; 8% African American | 59% Male; 41% Female | SMI | Offending | See Barnett et al, 2022 |
| **Dabit et al, 2021**[u] | USA, Canada, India, France, UK | RCT pilot trial | 31 | NR | 35% Male; 52% Female; 13% Non-binary | SMI | Social connectedness | 96 |
| **Danielsson et al, 2020**[u] | Sweden | Pilot trial | 42 | NR | 21% Male; 79% Female | CMD | Employment | 100 |
| **Dark et al, 2020**[u] | Australia | RCT | 120 | 7% Aboriginal/ Torres Strait Islander | 72% Male; 28% Female | CMD | Social connectedness | 100 |
| **Davidson et al, 2004**[b] | USA | RCT | 260 | 82% White; 11% African American; 2% Hispanic or Latino; 1% Asian/ Pacific Islander | 43% Male; 57% Female | CMD, SMI | Social connectedness | See Barnett et al, 2022 |
| **Davis et al, 2012**[b] | USA | RCT | 85 | 72% African American; 27% Caucasian; 1% Native American | 88% Male; 12% Female | SMI | Employment | See Barnett et al, 2022 |
| **Davis et al, 2015**[b] | USA | Feasibility RCT | 34 | 62% African American; 38% White | 97% Male; 3% Female | SMI | Employment | See Barnett et al, 2022 |
| **Davis et al, 2018**[b] | USA | RCT | 541 | 51% White; 42% African American; 17% Hispanic, Spanish or Latino | 82% Male; 18% Female | PTSD | Employment | See Barnett et al, 2022 |
| **Davis et al, 2022**[u] | USA | RCT | 119 | 73% Black; 26% White; 2% Hispanic; 1% Other | 82% Male; 18% Female | CMD, SMI | Employment | 88 |
| **de Waal et al, 2019**[b] | Netherlands | RCT | 250 | 72% Dutch; 8% Other; 6% Surinamese; 6% European; 4% Moroccan; 2% Dutch Antilles | 70% Male; 30% Female | CMD, SMI | Trauma | See Barnett et al, 2022 |
| **de Waal et al, 2021**[u] | Netherlands | Economic evaluation | 250 | NR | 70% Male; 30% Female | CMD, SMI | Trauma | 100 |
| **de Weerd et al, 2016**[b] | Netherlands | RCT | 60 | NR | 53% Male; 47% Female | CMD | Employment | See Barnett et al, 2022 |

| | | | | | | | | |
|---|---|---|---|---|---|---|---|---|
| **de Winter et al, 2020**[k] | Netherlands | Program fidelity study | 27 IPS Programs | NR | NR | SMI | Employment | |
| **Dehn et al, 2022**[u] | Germany | Quasi-experimental | 334 | NR | 62% Male; 38% Female | SMI | Housing | 100 |
| **Dogu et al, 2021**[u] | Turkey | RCT | 60 | NR | 62% Male; 38% Female | SMI | Social connectedness, Community | 92 |
| **Doré-Gauthier et al, 2020**[u] | Canada | Prospective longitudinal study | 50 | 68% 'Visible minority' | 92% Male; 8% Female | SMI | Housing | 91 |
| **Dubreucq et al, 2020**[u] | France | Quasi-experimental | 87 | NR | 80% Male; 20% Female | SMI | Social connectedness | 100 |
| **Easter et al, 2021**[k] | USA | RCT | 145 | 28% White; 67% Black; 6% Other Ethnicity; 2% Latino | 57% Male; 43% Female | SMI | Community | Quality score reported in Killaspy et al, 2022 |
| **Ebrahim et al, 2018**[k] | UK | Mixed-methods evaluation | 56 | NR | 34% Male; 66% Female | NR | Education | Quality score reported in Killaspy et al, 2022 |
| **Edge et al, 2018**[k] | UK | Feasibility/ acceptability cohort study | 31 | 55% African-Caribbean; 19% Mixed White and African-Caribbean; 13% Black British; 10% Mixed background; 3% Other Black background | 68% Male; 32% Female | SMI | Family | Quality score reported in Killaspy et al, 2022 |
| **Elbogen et al, 2016**[b] | USA | RCT | 184 | 74% Non-White | 81% Male; 19% Female | CMD, SMI | Debt, Financial | See Barnett et al, 2022 |
| **Ellison et al, 2020**[b] | USA | RCT | 166 | 47% Minority race | 93% Male; 7% Female | SMI | Housing | See Barnett et al, 2022 |

| | | | | | | | | |
|---|---|---|---|---|---|---|---|---|
| **Erickson et al, 2021**[b] | Canada | RCT | 109 | NR | 78% Male; 21% Female | SMI | Employment | See Barnett et al, 2022 |
| **Favrod et al, 2019**[k] | France | Pre-post implementation | 21 | NR | 76% Male; 24% Female | SMI | Social connectedness | Quality score reported in Killaspy et al, 2022 |
| **Finnes et al, 2022**[u] | Sweden | RCT | 264 | NR | 21% Male; 79% Female | CMD | Employment | 100 |
| **Fitzpatrick et al, 2021**[u] | Canada | Pilot pre-post interventional design | 14 (7 couples) | 100% White/ Caucasian/ European origin | 86% Male; 14% Female | SMI | Family | 90 |
| **Fletcher et al, 2008**[b] | USA | RCT | 191 | 71% Minority ethnicity, primarily African American; 28% Caucasian | 80% Male; 20% Female | SMI | Housing, Social connectedness | See Barnett et al, 2022 |
| **Fowler et al, 2019**[b] | UK | RCT | 77 | 86% White | 71% Male; 29% Female | SMI | Social connectedness | See Barnett et al, 2022 |
| **Gammelgaard et al, 2017**[k] | Denmark | Qualitative interviews | 12 | NR | 75% Male; 25% Female | SMI | Employment | Quality score reported in Killaspy et al, 2022 |
| **Gelkopf et al, 1994**[b] | Israel | RCT | 34 | NR | 82% Male; 18% Female | SMI | Social connectedness | See Barnett et al, 2022 |
| **Gjengedal et al, 2020**[u] | Norway | Quasi-experimental | 410 | NR | 25% Male; 75% Female | CMD | Employment | 92 |
| **Glynn et al, 2004**[b] | USA | RCT | 63 | 44% Caucasian; 40% African American; 13% Hispanic; 3% Asian | 92% Male; 8% Female | SMI | Social connectedness | See Barnett et al, 2022 |
| **Glynn et al, 2017**[k] | USA | RCT | 107 | 77% White; 2% Latino; 3% Asian; 15% Black; 3% Other | 82% Male; 18% Female | SMI | Employment | Quality score reported in Killaspy et al, 2022 |

| | | | | | | | | |
|---|---|---|---|---|---|---|---|---|
| **Goldfinger et al, 1999**[b] | USA | RCT | 303 | 41% African American | 72% Male; 28% Female | SMI | Housing | See Barnett et al, 2022 |
| **Granholm et al, 2005**[b] | USA | RCT | 76 | 79% Caucasian | 74% Male; 26% Female | SMI | Social connectedness | See Barnett et al, 2022 |
| **Gutman et al, 2009**[b] | USA | Quasi-experimental | 38 | 39% Hispanic, 37% African American; 21% White | 58% Male; 42% Female | CMD, SMI | Education | See Barnett et al, 2022 |
| **Gutman et al, 2017**[k] | USA | Non-randomised controlled trial | 15 | 13% White; 53% African American; 34% Hispanic | 100% Male | CMD, SMI | Housing | Quality score reported in Killaspy et al, 2022 |
| **Hall et al, 2018**[k] | Australia | Mixed-methods evaluation | 51 | NR | 17% Male; 82% Female; 1% Gender-free | NR | Education | Quality score reported in Killaspy et al, 2022 |
| **Hansen et al, 2023**[u] | Denmark | RCT follow-up | 547 | NR | 48% Male; 512% Female | SMI | Social connectedness, Family | 96 |
| **Hanssen et al, 2020**[u] | Netherlands | RCT | 50 | NR | 64% Male; 36% Female | SMI | Social connectedness | 81 |
| **Harris et al, 2017**[b] | Australia | RCT | 86 | NR | 64% Male; 36% Female | SMI | Employment | See Barnett et al, 2022 |
| **Haslam et al, 2019**[b] | Australia | RCT | 120 | 74% Caucasian; 69% Australian nationality | 36% Male; 64% Female | CMD, SMI | Social connectedness | See Barnett et al, 2022 |
| **Hasson-Ohayon et al, 2014**[b] | Israel | RCT | 55 | NR | 56% Male; 44% Female | SMI | Social connectedness | See Barnett et al, 2022 |
| **Hasson-Ohayon et al, 2019**[k] | Israel | RCT | 158 | NR | 59% Male; 41% Female | SMI | Social connectedness | Quality score reported in Killaspy et al, 2022 |

| | | | | | | | | |
|---|---|---|---|---|---|---|---|---|
| **Heatherington et al, 2019[k]** | USA | Non-controlled pre-post implementation | 259 | 95% Caucasian, non-Latinx; 1% Latinx; 3% African American; 1% Asian American; 1% Mixed race; 2% Other | 68% Male; 32% Female | CMD, SMI | Community | Quality score reported in Killaspy et al, 2022 |
| **Hees et al, 2013[b]** | Netherlands | RCT | 117 | NR | 49% Male; 51% Female | SMI | Employment | See Barnett et al, 2022 |
| **Hellstrom et al, 2017[b]** | Denmark | RCT | 326 | NR | 32% Male; 68% Female | CMD, SMI | Employment | See Barnett et al, 2022 |
| **Henderson et al, 2013[b]** | UK | RCT | 80 | 47% Black; 38% White; 11% Other; 4% Asian | 52% Male; 48% Female | CMD, SMI | Employment | See Barnett et al, 2022 |
| **Herman et al, 2011[b]** | USA | RCT | 150 | 62% African American; 17% White; 15% Latino; 6% Other | 71% Male; 29% Female | SMI | Housing | See Barnett et al, 2022 |
| **Higgins et al, 2018[k]** | Ireland | Mixed-methods pre-post evaluation | 128 (40 service users; 24 family members; 63 clinicians) | NR | 20% Male; 80% Female | SMI | Education | Quality score reported in Killaspy et al, 2022 |
| **Higgins et al, 2019[k]** | Ireland | Mixed-methods pre-post survey | 168 (paired analyses on 86 service users and family members) | NR | 29% Male; 71% Female | SMI | Education | Quality score reported in Killaspy et al, 2022 |
| **Himle et al, 2014[b]** | USA | RCT | 58 | 86% African American; 10% White; 4% Multiracial | 67% Male; 33% Female | CMD, SMI | Employment | See Barnett et al, 2022 |
| **Holmas et al, 2021[u]** | Norway | RCT | 327 | NR | 50% Male; 50% Female | NR | Employment | 88 |
| **Holmes et al, 2017[k]** | Australia | Quasi-experimental | 162 | NR | 78% Male; 22% Female | SMI | Housing | Quality score reported in |

| | | | | | | | | |
|---|---|---|---|---|---|---|---|---|
| | | prospective cohort study | | | | | | Killaspy et al, 2022 |
| **Holt et al, 2021**[u] | Australia | RCT | 77 | 68% Born in Australia | 100% Female | CMD | Family | 100 |
| **Horan et al, 2018**[k] | USA | RCT | 139 | 58% White; 33% African American; 3% Asian; 3% Hawaiian/ Pacific Islander; 3% Mixed race; 78% Non-Hispanic; 22% Hispanic | 70% Male; 30% Female | SMI | Social connectedness | Quality score reported in Killaspy et al, 2022 |
| **Hui et al, 2023**[u] | Hong Kong | RCT | 360 | 100% Chinese | 44% Male; 56% Female | SMI | Social connectedness | 100 |
| **Hurlburt et al, 1996**[b] | USA | RCT | 361 | 63% White; 20% Black; 13% Hispanic; 5% Other | 67% Male; 33% Female | SMI | Housing | See Barnett et al, 2022 |
| **Hutchinson et al, 2018**[k] | UK | Mixed-methods evaluation | 6 sites (including 235 staff and 65 service users) | NR | NR | NR | Employment | Quality score reported in Killaspy et al, 2022 |
| **Inman et al, 2021**[u] | UK | Feasibility study | 16 | NR | 75% Male; 25% Female | SMI | Community | 95 (qual and quant) |
| **Johnson et al, 2020**[u] | USA | RCT | 172 | 47% White; 44% Black African American; 9% Multiracial; 4% Hispanic or Latina ; 96% Not Hispanic or Latina | 100% Female | CMD, SMI | Trauma, Victimisation | 89 |
| **Karasz et al, 2021**[u] | Bangladesh | Pilot RCT | 48 | NR | 100% Female | NR | Other – Financial | 88 |
| **Kayo et al, 2020**[k] | Brazil | RCT | 62 | NR | 71% Male; 29% Female | SMI | Social connectedness | Quality score reported in Killaspy et al, 2022 |
| **Kerman et al, 2020**[u] | Canada | RCT | 2111 | 21% Indigenous | 67% Male; 33% Female | SMI | Housing | 88 |

| | | | | | | | | |
|---|---|---|---|---|---|---|---|---|
| **Kern et al, 2018[b]** | USA | RCT | 58 | 65% White | 84% Male; 16% Female | SMI | Employment | See Barnett et al, 2022 |
| **Kern et al, 2022[u]** | USA | RCT | 91 | 81% Non-White; 19% White | 90% Male; 10% Female | SMI | Employment | 92 |
| **Khalifa et al, 2020[u]** | UK | Feasibility cluster RCT | 18 | 72% White British; 17% Black; 11% Mixed | 89% Male; 11% Female | SMI | Employment | 88 |
| **Kidd et al, 2021[u]** | Canada | RCT | 110 | 24% Black or Afro-Caribbean or African; 9% East Asian or Asian; 8% South Asian; 4% Latino or Hispanic; 7% Middle Eastern or Arabic; 7% Other; Non -Hispanic White or European 43% | 62% Male; 38% Female | SMI | Community | 100 |
| **Killackey et al, 2019[b]** | Australia | RCT | 146 | NR | 69% Male; 31% Female | SMI | Employment | See Barnett et al, 2022 |
| **Killaspy et al, 2016[k]** | UK | National survey | 619 | 81% White | 66% Male; 34% Female | SMI | Housing | Quality score reported in Killaspy et al, 2022 |
| **Killaspy et al, 2020[k]** | UK | National prospective cohort study | 586 | See Killaspy et al 2016 | NR | SMI | Housing | Quality score reported in Killaspy et al, 2022 |
| **Kingston et al, 2018[b]** | Canada | RCT | 101 | 88% Caucasian; 11% Aboriginal; 9% Asian; 6% Black | 100% Male | SMI | Offending | See Barnett et al, 2022 |
| **Kirst et al, 2020[u]** | Canada | RCT | Qualitative n = 150 Quantitative n = 2132 | 'Visible minority' 40%; Country of birth: Canada 81%; Outside of Canada 19% | 67% Male; 33% Female | NR | Housing | 88 |
| **Knight et al, 2021[u]** | Australia | RCT | 112 | NR | 34% Male; 66% Female | SMI | Social connectedness | 88 |

| | | | | | | | | |
|---|---|---|---|---|---|---|---|---|
| **Korr & Joseph, 1995**[b] | USA | RCT | 95 | 43% Black; 49% White; 8% Other | 80% Male; 20% Female | SMI | Housing | See Barnett et al, 2022 |
| **Kukla et al, 2018**[b] | USA | RCT | 75 | 57% African American; 41% White; 1% Hispanic American | 93% Male; 7% Female | SMI | Employment | See Barnett et al, 2022 |
| **Kumar et al, 2020**[k] | India | Single-blind RCT | 66 service users (& their relatives) | NR | NR | SMI | Family, Education | Quality score reported in Killaspy et al, 2022 |
| **Lachaud et al, 2021**[u] | Canada | RCT | 543 | 59% Non-White; 41% White | 68% Male; 32% Female | SMI | Housing | 88 |
| **Lamberti et al, 2017**[b] | USA | RCT | 70 | 73% African American; 19% Caucasian; 8% Hispanic | 61% Male; 39% Female | SMI | Offending | See Barnett et al, 2022 |
| **Latimer et al, 2020**[u] | Canada | RCT | 950 | NR | 68% Male; 31% Female | SMI | Housing | 96 |
| **Laurila et al, 2024**[u] | Finland | RCT | 326 | NR | 49% Male; 51% Female | CMD, SMI | Housing | 81 |
| **Lecomte et al, 2020**[b] | Canada | RCT | 164 | 63% Caucasian | 61% Male; 39% Female | CMD | Employment | See Barnett et al, 2022 |
| **Lehman et al, 1997**[b] | USA | RCT | 152 | 72% African American; 24% White | 67% Male; 33% Female | SMI | Housing | See Barnett et al, 2022 |
| **Lemoine et al, 2021**[u] | France | RCT | 704 | NR | 83% Male; 17% Female | SMI | Housing | 95 |
| **Lerner et al, 2020**[u] | USA | RCT | 253 | 53% White | 86% Male; 14% Female | SMI | Employment | 100 |
| **Liang et al, 2023**[u] | China | Quasi-experimental | 104 | NR | 44% Male; 56% Female | SMI | Community, Social connectedness | 83 |

| | | | | | | | | |
|---|---|---|---|---|---|---|---|---|
| **Lindenmeyer et al, 2008**[b] | USA | RCT | 85 | 58% Black; 27% Hispanic; 13% White; 3% Asian | 89% Male; 11% Female | SMI | Employment | See Barnett et al, 2022 |
| **Lipton et al, 1988**[b] | USA | RCT | 49 | NR | 65% Male; 35% Female | SMI | Housing | See Barnett et al, 2022 |
| **Lloyd-Evans et al, 2020**[b] | UK | Feasibility RCT | 129 | 64% White; 13% Black African/ Caribbean or Black British; 10% Asian/ Asian British; 8% Multiple ethnic groups; 5% Other ethnic groups | 27% Male; 73% Female | CMD | Social connectedness | See Barnett et al, 2022 |
| **Lobban et al, 2020a**[k] | UK | Online RCT | 800 | 91% White British; 1% White Irish; 4% Other White background; 2% Mixed ethnic background; 2% Asian or Asian British; 1% Other ethnic group; <0.5% Prefer not to say | 19% Male; 81% Female; <1% Not applicable | SMI | Family | Quality score reported in Killaspy et al, 2022 |
| **Loubiere et al, 2022**[u] | France | RCT follow-up | 703 | 82% French nationality | 82% Male; 18% Female | SMI | Housing | 100 |
| **Lysaker et al, 2005**[b] | USA | RCT | 50 | 56% African American; 42% Caucasian; 2% Latino | 100% Male | SMI | Employment | See Barnett et al, 2022 |
| **Lystad et al, 2017**[k] | Norway | Unblinded RCT | 131 | NR | 70% Male; 30% Female | SMI | Employment | Quality score reported in Killaspy et al, 2022 |
| **Macnaughton et al, 2018**[k] | Canada | Process evaluation | NA | 'Indigenous people constitute the majority of people experiencing homelessness [in the research location]' | NA | SMI | Housing | Quality score reported in Killaspy et al, 2022 |
| **Mahlke et al, 2017**[k] | Germany | RCT | 216 | NR | 50% Male; 50% Female | SMI | Community | Quality score reported in Killaspy et al, 2022 |

| | | | | | | | | |
|---|---|---|---|---|---|---|---|---|
| **Marder et al, 1996**[b] | USA | RCT | 80 | 69% Non-White | 100% Male | SMI | Social connectedness | See Barnett et al, 2022 |
| **Martin-Carrasco et al, 2016**[k] | Spain, Portugal | RCT | 223 | NR | 24% Male; 76% Female | SMI | Family | Quality score reported in Killaspy et al, 2022 |
| **Martini et al, 2022**[u] | Italy | RCT | 311 | NR | 62% Male; 38% Female | SMI | Employment | 69 |
| **Maru et al, 2021**[u] | USA | RCT | 166 | 62% White; 38% Non-White or Mixed race; 22% Latino or Hispanic | 49% Male; 51% Female | CMD, SMI | Employment | 92 |
| **McGurk et al, 2016**[b] | USA | RCT | 54 | 24% White; 61% African American; 15% Multi-racial<br>19% Hispanic/ Latino; 81% Non-Hispanic/ Latino | 70% Male; 30% Female | SMI | Employment | See Barnett et al, 2022 |
| **McGurk et al, 2007**[b] | USA | RCT | 44 | 68% African American; 16% Hispanic; 14% Caucasian; 2% Asian | 55% Male; 45% Female | SMI | Employment | See Barnett et al, 2022 |
| **McGurk et al, 2015**[b] | USA | RCT | 107 | 86% White; 10% Black or African American; 2% Asian; 2% Mixed | 65% Male; 35% Female | SMI | Employment | See Barnett et al, 2022 |
| **McGurk et al, 2017**[k] | USA | Feasibility RCT | 83 | 53% White; 42% Non-White; 4% Missing | 58% Male; 42% Female | SMI | Employment | Quality score reported in Killaspy et al, 2022 |
| **McHugo et al, 2004**[b] | USA | RCT | 121 | 83% African American | 48% Male; 52% Female | SMI | Housing | See Barnett et al, 2022 |
| **Mejia-Lancheros et al, 2020**[u] | Canada | RCT | 381 | 43% White; 57% Non-White | 68% Male; 32% Female | SMI | Housing | 96 |
| **Mervis et al, 2017**[b] | USA | RCT | 64 | NR | 61% Male; 39% Female | SMI | Employment | See Barnett et al, 2022 |

| | | | | | | | | |
|---|---|---|---|---|---|---|---|---|
| **Metts et al, 2023**[u] | USA | RCT | 250 | 41% Black. African American; 36% Non-Hispanic White/ Caucasian; 9% Multi-racial | 41% Male; 59% Female | CMD | Employment | 88 |
| **Milligan-Saville et al, 2017**[b] | Australia | Cluster RCT | 85 | NR | 100% Male | NR | Employment | See Barnett et al, 2022 |
| **Minor et al, 2022**[u] | USA | RCT | 20 | 75% African American; 25% Caucasian | 45% Male; 55% Female | SMI | Social connectedness | 88 |
| **Mirsepassi et al, 2018**[k] | Iran | Pilot implementation | 301 | NR | 57% Male; 43% Female | SMI | Family | Quality score reported in Killaspy et al, 2022 |
| **Monson et al, 2022**[u] | Canada | Uncontrolled trial | 17 couples | 91% White Caucasian; 9% Aboriginal/ First Nations/ Metis/ Inuit; | 47% Male; 53% Female; 0% Non-binary | SMI | Social connectedness | 86 |
| **Morse et al, 1992**[b] | USA | RCT | 116 | 53% Non-White | 58% Male; 42% Female | SMI | Housing | See Barnett et al, 2022 |
| **Morse et al, 1997**[b] | USA | RCT | 165 | 45% Caucasian; 55% African American | 58% Male; 42% Female | SMI | Housing | See Barnett et al, 2022 |
| **Morse et al, 2006**[b] | USA | RCT | 149 | 73% African American; 25% Caucasian; 2% Other minority group | 80% Male; 20% Female | SMI | Housing | See Barnett et al, 2022 |
| **Motteli et al, 2022**[u] | Switzerland | Pragmatic RCT | 58 | 62% Swiss nationals | 38% Male; 62% Female | SMI | Housing | 92 |
| **Moxham et al, 2017**[k] | Australia | Qualitative evaluation | 27 | NR | 63% Male; 37% Female | CMD, SMI | Social connectedness | Quality score reported in Killaspy et al, 2022 |
| **Mueller et al, 2023**[u] | USA | RCT | 54 | 78% Caucasian; 13% Black or African American; 6% Asian; 22% Hispanic or Latino; 76% Not Hispanic or Latino | 15% Male; 85% Female | CMD | Social connectedness | 96 |
| **Mueser et al, 2005**[b] | USA | RCT | 35 | 97% Non-Hispanic White; 3% Asian | 80% Male; 20% Female | SMI | Employment | See Barnett et al, 2022 |

| | | | | | | | | |
|---|---|---|---|---|---|---|---|---|
| **Mustafa et al, 2022**[u] | Canda | Single-blind RCT | 220 | NR | 69% Male; 31% Female | SMI | Employment, Education | 92 |
| **Nezafat et al, 2023**[u] | Iran | RCT | 41 | NR | NR | CMD | Social connectedness | 73 |
| **Nguyen et al, 2020**[k] | Vietnam | Pilot proof-of-concept trial | 68 | NR | 37% Male; 63% Female | SMI | Community | Quality score reported in Killaspy et al, 2022 |
| **Niedermoser et al, 2020**[u] | Switzerland | RCT | 27 | NR | 48% Male; 52% Female | SMI | Employment | 92 |
| **Nijman et al, 2023**[u] | Netherlands | Single-blind RCT | 81 | NR | 70% Male; 30% Female | SMI | Social connectedness | 100 |
| **Noordik et al, 2013**[b] | Netherlands | Cluster RCT | 35 occupational physicians and 158 clients | NR | 34% Male; 66% Female | CMD | Employment | See Barnett et al, 2022 |
| **Nuechterlein et al, 2019**[b] | USA | RCT | 69 | 26% White; 12% Asian; 22% African American; 38% Mixed race Ethnicity: 26% Hispanic/ Latino | 67% Male; 33% Female | SMI | Employment | See Barnett et al, 2022 |
| **O'Connell et al, 2018**[k] | USA | RCT | 76 | 59% White; 26% Black; 25% Other | 50% Male; 50% Female | SMI | Community | Quality score reported in Killaspy et al, 2022 |
| **O'Campo et al, 2023**[u] | Canada | RCT – Secondary analysis | 653 | 24% Aboriginal; 53% White; 23% 'Ethno-racial' | 100% Female | CMD, SMI | Housing | 100 |
| **O'Connell et al, 2023**[u] | USA | RCT – Secondary analysis | 272 | 31% White; 62% Black; 4% Hispanic or Latino | 94% Male; 6% Female | CMD, SMI | Housing | 83 |

| | | | | | | | | |
|---|---|---|---|---|---|---|---|---|
| **Ohki et al, 2021**[u] | Japan | Retrospective cohort study | 323 | NR | 80% Male; 20% Female | SMI | Employment | 100 |
| **Okpaku et al, 1997**[b] | USA | RCT | 152 | 60% White | 59% Male; 41% Female | CMD, SMI | Employment | See Barnett et al, 2022 |
| **Overland et al, 2018**[b] | Norway | RCT | 1193 | NR | 33% Male; 67% Female | CMD | Employment | See Barnett et al, 2022 |
| **Oxford et al, 2021**[u] | USA | RCT | 252 | 65% White; 17% Black; 10% Mixed race; 7% Unknown; 53% Hispanic or Latino | 100% Female | CMD | Family | 100 |
| **Padamaker et al, 2020**[k] | India | Mixed-methods evaluation | 11 | NR | 100% Female | NR | Housing | Quality score reported in Killaspy et al, 2022 |
| **Perkins et al, 2023**[u] | UK | Unblinded RCT | 89 | 67% White; 8% Asian; 6% Black; 2% Arab; 5% Other; 1% Prefer not to say | 100% Female | CMD | Family | 100 |
| **Perlick et al, 2018**[k] | USA | RCT | 43 | NR | 16% Male; 84% Female | SMI | Family | Quality score reported in Killaspy et al, 2022 |
| **Perrez-Corrales et al, 2019**[k] | Spain | Qualitative evaluation | 23 | NR | 70% Male; 30% Female | SMI | Community | Quality score reported in Killaspy et al, 2022 |
| **Pichler et al, 2021**[u] | Germany | RCT follow-up | 114 | NR | 46% Male; 54% Female | CMD, SMI | Employment | 90 |
| **Pos et al, 2019**[b] | Netherlands | RCT | 99 | 60% Ethnic minority | 81% Male; 19% Female | SMI | Social connectedness | See Barnett et al, 2022 |
| **Pot-Kolder et al, 2018**[b] | Netherlands | Single-blind RCT | 116 | 34% Non-Dutch origin | 71% Male; 29% Female | SMI | Social connectedness | See Barnett et al, 2022 |

| | | | | | | | | |
|---|---|---|---|---|---|---|---|---|
| **Priebe et al, 2020**[b] | UK | RCT | 124 | 15% White; 2% Arab; 2% White Other; 15% Black Caribbean; 19% Black African; 6% Black Other; 4% Indian Pakistani; 19% Bangladeshi; 4% Asian Other; 2% Mixed or Multiple; 9% Unspecified | 65% Male; 35% Female | SMI | Social connectedness | See Barnett et al, 2022 |
| **Prince et al, 2018**[k] | USA | Qualitative interviews | 20 | NR | NR | SMI | Social connectedness | Quality score reported in Killaspy et al, 2022 |
| **Puig et al, 2016**[k] | USA | RCT | 40 | NR | 60% Male; 40% Female | SMI | Employment | Quality score reported in Killaspy et al, 2022 |
| **Rajji et al, 2022**[u] | Canada | RCT | 63 | NR | 51% Male; 49% Female | SMI | Social connectedness | 100 |
| **Raven et al, 2020**[u] | USA | RCT | 423 | 65% White; 25% Hispanic; 14% Black; 21% Other | 71% Male; 29% Female | SMI | Housing | 96 |
| **Rebergen et al, 2009**[b] | Netherlands | RCT | 240 | NR | 56% Male; 44% Female | CMD | Employment | See Barnett et al, 2022 |
| **Reme et al, 2019**[b] | Norway | RCT | 410 | NR | 51% Male; 49% Female | CMD, SMI | Employment | See Barnett et al, 2022 |
| **Rhenter et al, 2018**[k] | France | Qualitative evaluation of an RCT | 13 | NR | NR | SMI | Housing | Quality score reported in Killaspy et al, 2022 |
| **Rivera et al, 2007**[b] | USA | RCT | 203 | 29% Caucasian; 17% African American; 31% Hispanic | 51% Male; 49% Female | CMD, SMI | Community | See Barnett et al, 2022 |
| **Roberts et al, 2014**[b] | USA | RCT | 66 | 64% Caucasian; 36% African American<br>Ethnicity: 6% Hispanic or Latino | 67% Male; 33% Female | SMI | Social connectedness | See Barnett et al, 2022 |

| | | | | | | | | |
|---|---|---|---|---|---|---|---|---|
| **Rodríguez Pulido et al, 2019[b]** | Spain | RCT | 57 | NR | 68% Male; 32% Female | SMI | Employment | See Barnett et al, 2022 |
| **Rogers et al, 2006[b]** | USA | RCT | 135 | 30% African American; 58% Caucasian; 10% Other | 54% Male; 46% Female | SMI | Employment | See Barnett et al, 2022 |
| **Roos et al, 2016[k]** | Norway | Qualitative evaluation | 14 | NR | NR | SMI | Housing | Quality score reported in Killaspy et al, 2022 |
| **Rossler et al, 2020[b]** | Switzerland | RCT | 116 | NR | 49% Male; 51% Female | CMD, SMI | Employment | See Barnett et al, 2022 |
| **Rouse et al, 2017[k]** | Canada | Interviews | NR | NR | NR | SMI | Community | |
| **Rowe et al, 2007[b]** | USA | RCT | 114 | 58% African American; 31% White; 15% Hispanic; 9% Other; 3% Native American | 68% Male; 32% Female | SMI | Offending | See Barnett et al, 2022 |
| **Ruiz-Comellas et al, 2022[u]** | Spain | RCT | 90 | NR | 23% Male; 77% Female | CMD | Social connectedness | 92 |
| **Russinova et al, 2018[b]** | USA | RCT | 51 | 65% Non-Hispanic White; 35% Minority ethnic group | 39% Male; 61% Female | CMD, SMI | Employment | See Barnett et al, 2022 |
| **Russinova et al, 2023[u]** | USA | RCT | 185 | 54% White; 19% Black; 5% American Indian or Alaskan Native; 1% Asian; 1% Asian/ Pacific Islander; 19% Two or more races<br>Ethnicity: 19% Hispanic or Latino | 38% Male; 61% Female; 1% Unknown | CMD, SMI | Community | 85 |
| **Saavedra et al, 2018[k]** | Spain | Qualitative interviews | 19 (11 service users and 8 service providers) | NR | 64% Male; 36% Female (service users) | SMI | Community | Quality score reported in Killaspy et al, 2022 |

| | | | | | | | | |
|---|---|---|---|---|---|---|---|---|
| **Sacks et al, 2004**[b] | USA | RCT | 185 | 49% Caucasian; 30% Black; 17% Hispanic; 4% Other | 100% Male | SMI | Offending | See Barnett et al, 2022 |
| **Sacks et al, 2012**[b] | USA | RCT | 127 | 56% White; 17% Hispanic; 17% Other or Mixed; 10% Black | 100% Male | SMI | Offending | See Barnett et al, 2022 |
| **Salomonsson et al, 2020**[u] | Sweden | RCT | 59 (‘DepAnx’ group only) | NR | 29% Male; 71% Female | CMD | Employment | 91 |
| **Salzer et al, 2016**[b] | USA | RCT | 100 | 75% Black; 21% White; 4% Latin or Hispanic; 2% Native American; 2% Other race; 1% Asian | 53% Male; 46% Female; 1% Trans-gender | SMI | Community | See Barnett et al, 2022 |
| **Sanches et al, 2020**[b] | Netherlands | RCT | 188 | NR | 58% Male; 42% Female | SMI | Employment | See Barnett et al, 2022 |
| **Sanches et al, 2022**[u] | Netherlands | RCT | 188 | NR | 58% Male; 42% Female | CMD, SMI | Social connectedness | 100 |
| **Scanlan et al, 2019**[k] | Australia | Uncontrolled prospective study | 97 | NR | 47% Male; 53% Female | SMI | Employment | Quality score reported in Killaspy et al, 2022 |
| **Schene et al, 2007**[b] | Netherlands | RCT | 62 | NR | 48% Male; 52% Female | SMI | Employment | See Barnett et al, 2022 |
| **Schneider et al, 2016**[k] | UK | Feasibility study | 74 | 68% White British; 4% Other White; 17% Black British; 11% Other ethnic group | 70% Male; 30% Female | SMI | Employment | Quality score reported in Killaspy et al, 2022 |
| **Schramm et al, 2020**[u] | Germany | Pilot RCT | 28 | NR | 21% Male; 79% Female | CMD | Employment | 92 |
| **Segal et al, 2010**[b] | USA | RCT | 505 | 36% White; 34% African American; 30% Other | 53% Male; 47% Female | SMI | Community | See Barnett et al, 2022 |
| **Seoane-Bouzas et al, 2021**[u] | Spain | Pilot RCT | 16 | NR | 63% Male; 37% Female | SMI | Social connectedness | 96 |

| | | | | | | | | |
|---|---|---|---|---|---|---|---|---|
| **Shen et al, 2022**[u] | China | Pilot RCT | 87 | NR | 66% Male; 34% Female | SMI | Social connectedness | 100 |
| **Sheridan et al, 2015**[b] | Ireland | RCT | 107 | NR | 48% Male; 52% Female | SMI | Social connectedness | See Barnett et al, 2022 |
| **Shern et al, 2000**[b] | USA | RCT | 168 | 61% Black; 29% White; 10% Hispanic | 66% Male; 34% Female | SMI | Housing | See Barnett et al, 2022 |
| **Shih et al, 2023**[u] | Taiwan | RCT | 90 | NR | 50% Male; 50% Female | SMI | Social connectedness | 100 |
| **Shimada et al, 2022**[u] | Japan | RCT | 102 | NR | 54% Male; 46% Female | SMI | Social connectedness | 100 |
| **Sikira et al, 2021**[u] | Bosnia and Herzegovina | Exploratory RCT | 65 | NR | 42% Male; 58% Female | SMI | Social connectedness | 96 |
| **Silverman et al, 2014**[b] | USA | RCT | 45 | 71% Caucasian American; 13% African American; 9% Other; 4% Hispanic American | 49% Male; 51% Female | SMI | Social connectedness | See Barnett et al, 2022 |
| **Smidl et al, 2017**[k] | USA | Mixed-methods evaluation | 20 | NR | NR | SMI | Community | Quality score reported in Killaspy et al, 2022 |
| **Smith et al, 2022**[u] | USA | RCT | 90 | 66% Black African American; 26% White; 8% Latinx; 1% More than one race | 57% Male; 43% Female | SMI | Employment | 96 |
| **Snethen et al, 2024**[u] | USA | RCT | 74 | 78% Black African American; 18% White; 9% American Indian or Alaska Native; 3% Asian; 8% Other Ethnicity: 5% Hispanic or Latino | 57% Male; 43% Female | SMI | Community | 83 |
| **Solar et al, 2023**[u] | Australia | Single-blind RCT | 51 | NR | 67% Male; 33% Female | SMI | Employment | 79 |

| | | | | | | | | |
|---|---|---|---|---|---|---|---|---|
| **Somers et al, 2017**[k] | USA | Unblinded RCT | 297 | 15% Aboriginal; 57% White; 28% Mixed or Other | 73% Male; 23% Female | SMI | Housing | Quality score reported in Killaspy et al, 2022 |
| **Sommer et al, 2019**[k] | Australia | Observational study | 64 | NR | 27% Male; 73% Female | NR | Education, Community | Quality score reported in Killaspy et al, 2022 |
| **Sroosh et al, 2023**[u] | Iran | Quas-experimental | 60 | NR | NR | CMD | Family | 72 |
| **Stanhope et al, 2016**[k] | USA | Longitudinal qualitative study | NA | NA | NA | SMI | Housing | Quality score reported in Killaspy et al, 2022 |
| **Stergiopoulos et al, 2015**[b] | Canada | Unblinded RCT | 1198 | 24% Aboriginal; 28% Ethno-racial; 48% White | 66% Male; 33% Female; 1% Other | SMI | Housing | See Barnett et al, 2022 |
| **Stergiopoulos et al, 2016*a***[k] | Canada | Unblinded RCT | 237 | 22% Black Caribbean; 18% Black African; 11% Mixed; 10% South Asian; 25% Other | 66% Male; 34% Female | SMI | Housing | Quality score reported in Killaspy et al, 2022 |
| **Stergiopoulos et al, 2016*b***[k] | Canada | Qualitative fidelity programme assessment | 19 | NR | NR | SMI | Housing | Quality score reported in Killaspy et al, 2022 |
| **Stroupe et al, 2021**[u] | USA | RCT | 541 | 50% White; 42% African American; 13% Other; 17% Hispanic | 82% Male; 18% Female | SMI | Employment | 100 |
| **Susser et al, 1997**[b] | USA | RCT | 96 | 64% African American; 26% Other | 100% Male | SMI | Housing | See Barnett et al, 2022 |
| **Sutton et al, 2019**[k] | UK | Questionnaire | 108 | 90% White; 10% Other | 36% Male; 64% Female | NR | Employment | Quality score reported in Killaspy et al, 2022 |

| | | | | | | | | |
|---|---|---|---|---|---|---|---|---|
| **Swinkels et al, 2023**[u] | Netherlands | RCT | 102 | 40% White; 24% Black African or Caribbean; 17% Arabic or Northern African; 2% Asian; 18% Multiple ethnic groups | 88% Male; 12% Female | SMI | Social connectedness, Offending | 100 |
| **Talbot et al, 2018**[k] | UK | Feasibility RCT | 4 clusters | NR | NR | NR | Employment | Quality score reported in Killaspy et al, 2022 |
| **Taylor et al, 2020**[u] | USA | RCT | 29 | 71% White; 21% Asian; 4% Native American; 4% Pacific Islander; 21% Hispanic | 39% Male; 61% Female | CMD | Social connectedness | 85 |
| **Terzian et al, 2013**[b] | Italy | RCT | 357 | NR | 70% Male; 30% Female | SMI | Social connectedness | See Barnett et al, 2022 |
| **Thomas et al, 2018**[k] | USA | Secondary analysis of an RCT | 46 | 50% White; 41% Black; 7% Latino; 4% Native American; 2% Asian; 7% Other | 30% Male; 70% Female | SMI | Community | Quality score reported in Killaspy et al, 2022 |
| **Thompson et al, 2020**[u] | UK | Single arm proof-of-concept trial | 19 | NR | 26% Male; 74% Female | SMI | Social connectedness | 100 |
| **Tinland et al, 2020**[b] | France | RCT | 703 | 85% French nationality; 15% Other | 82% Male; 18% Female | SMI | Housing | See Barnett et al, 2022 |
| **Tjaden et al, 2021**[u] | Netherlands | RCT | 158 | 59% Dutch; 21% Western; 29% Non-Western; <1% Unknown | 59% Male; 41% Female | SMI | Social connectedness | 100 |
| **Tse et al, 2021**[u] | Hong Kong | RCT | 24 (invited from a larger RCT of N=176) | 100% Native Chinese | 25% Male; 75% Female | SMI | Community | 85 |
| **Tsemberis et al, 2004**[b] | USA | RCT | 206 | 27% White non-Hispanic; 41% Black non-Hispanic; 15% Hispanic; 18% Mixed or Other | 79% Male; 21% Female | SMI | Housing | See Barnett et al, 2022 |

| | | | | | | | | |
|---|---|---|---|---|---|---|---|---|
| **Twamley et al, 2019**[b] | USA | RCT | 153 | 38% racial or ethnic minority | 57% Male; 43% Female | SMI | Employment | See Barnett et al, 2022 |
| **Valentine et al, 2020**[u] | Australia | Qualitative evaluation of an RCT | 12 | NR | 42% Male; 58% Female | SMI | Social connectedness | 90 |
| **Valls et al, 2021**[u] | Spain | RCT | 65 | NR | 48% Male; 52% Female | SMI | Family, Social connectedness | 96 |
| **van Beurden et al, 2017**[b] | Netherlands | RCT | 3379 workers and 66 occupa-tional physicians | NR | 29% Male; 71% Female | CMD | Employment | See Barnett et al, 2022 |
| **van der Stouwe et al, 2022**[u] | Netherlands | RCT | 105 | NR | 60% Male; 40% Female | SMI | Trauma, Victimisation | 85 |
| **Van Lieshout et al, 2021**[u] | Canada | RCT | 403 | 72% White | 100% Female | CMD | Social connectedness, Family | 100 |
| **van Veen et al, 2021**[u] | Netherlands | RCT | 93 service users & 56 clinicians | 77% Dutch; 14% Other; 9% Missing | 30% Male; 70% Female | CMD | Social connectedness | 100 |
| **Varga et al, 2018**[k] | Hungary | RCT | 75 | NR | 51% Male; 49% Female | SMI | Community | Quality score reported in Killaspy et al, 2022 |
| **Vauth et al, 2005**[b] | Germany | RCT | 138 | NR | 35% Male; 65% Female | SMI | Employment | See Barnett et al, 2022 |
| **Vlasveld et al, 2013**[b] | Netherlands | RCT | 126 | NR | 46% Male; 54% Female | SMI | Employment | See Barnett et al, 2022 |

| | | | | | | | | |
|---|---|---|---|---|---|---|---|---|
| **Vogel et al, 2023**[u] | Netherlands | RCT | 43 | NR | 81% Male; 19% Female | SMI | Social connectedness | 100 |
| **Volker et al, 2015**[b] | Netherlands | RCT | 220 | 98% Dutch Nationality | 41% Male; 59% Female | CMD | Employment | See Barnett et al, 2022 |
| **Webber et al, 2021**[u] | UK | Non-randomised pre-post controlled trial | 151 service users across 5 NHS Trusts | 100% White (96% White; 1% White Irish; 3% White Other) | 30% Male; 70% Female | NR | Social connectedness | 92 |
| **Whitley et al, 2021**[k] | Canada | Qualitative interviews | 20 | NR | 60% Male; 40% Female | SMI | Social connectedness | Quality score reported in Killaspy et al, 2022 |
| **Wilson et al, 2019**[k] | UK | Mixed-methods evaluation | 101 | 92% White British; 4% Indian; 4% British Muslim | 44% Male; 56% Female | NR | Community, Education | Quality score reported in Killaspy et al, 2022 |
| **Winter et al, 2020**[u] | Germany | Feasibility study | 20 | NR | 25% Male; 75% Female | CMD | Employment | 65 |
| **Wong et al, 2023**[u] | Hong Kong | RCT | 127 | NR | 40% Male; 60% Female | SMI | Community, Social connectedness | 100 |
| **Worton et al, 2018**[k] | Canada | Case study evaluation | 6 sites | NR | NR | SMI | Housing | Quality score reported in Killaspy et al, 2022 |
| **Yamaguchi et al, 2017**[b] | Japan | RCT | 111 | NR | 68% Male; 32% Female | CMD, SMI | Employment | See Barnett et al, 2022 |
| **Yu et al, 2016**[k] | China | Qualitative interviews | 15 participants and 15 caregivers | NR | 67% Male; 33% Female | SMI | Employment | Quality score reported in Killaspy et al, 2022 |

| | | | | | | | | |
|---|---|---|---|---|---|---|---|---|
| **Zhang et al, 2017**[k] | China | RCT | 162 | NR | 42% Male; 58% Female | SMI | Employment | Quality score reported in Killaspy et al, 2022 |
| **Zhu et al, 2020**[u] | China | RCT | 157 | 100% Native Chinese | 54% Male; 46% Female | SMI | Social connectedness | 96 |

*nb. Gender and ethnicity percentages do not always equal one hundred due to either missing participant data or participants identifying with multiple race and/or ethnicity categories.*

NR = Not reported; [b] = Sourced from Barnett et al., 2022; [k] = Sourced from Killaspy et al., 2022; [u] = Sourced from updated searches; SMI = Severe Mental Illness; CMD = Common Mental Disorders.

#### Table 4: Example interventions for each domain

| Domain | Authors, date | Intervention name | Intervention description and aims |
|---|---|---|---|
| **Community support** | Russinova et al., 2023 | Bridging Community Gaps Photovoice | A peer-led photovoice-based intervention aiming to enhance community participation in individuals with a diagnosis of mental illness living in USA. It comprised a 6-month manualised peer-delivered intervention with parallel group and individual components. |
| **Debt and finance** | Karasz et al., 2021 | ASHA (Hope) Project | An integrated depression treatment and economic strengthening intervention for low-income women living in rural Bangladesh, aiming to improve their financial situation, status, autonomy and social relationships; and reduce depression. The intervention was delivered by peer health workers and comprised a 6-month group-based fortnightly depression management and financial literacy intervention, followed by a cash-transfer at 12 months' follow-up. |
| **Education** | Ebrahim et al., 2018 | Recovery College | A Recovery College in the UK, which serves as an education-based mental health resources, drawing on practioner and lived experience expertise to offer courses to people with mental ill-health. Core principles of Recovery Colleges include co-production, a focus on adult learning, open access, peer support and self-management. The purpose of this Recovery College is described as facilitating recovery through education that inspires hope, control over mental health problems and access to community connections |
| **Employment** | Killackey et al., 2019 | Individual Placement and Support (IPS) | This IPS intervention in Australia aimed to support young people with first episode psychosis into employment. It followed IPS principles: intensive, individual support provided by a vocational specialist who was embedded as a member of the clinical team; and a rapid job search followed by placement in paid employment. Participants received 6 months of the IPS intervention. |
| **Family relationships** | Battle et al., 2023 | Family treatment for postpartum depression | A structured, short-term family intervention aiming to improve family functioning in dimensions that typically need to be strengthened in families where postpartum depression occurs – for example, improving communication and adjusting to role changes. The intervention was delivered in the USA and comprised 10-12 1-hour sessions of a 16-week period. |
| **Housing** | Stergiopoulos et al., 2016 | Housing First (HF) | A Housing First intervention in Canada. HF is designed for adults experiencing chronic homeless and mental ill-health, providing permanent housing and clinical support without requiring treatment compliance as a prerequisite for housing access. The implementation of this intervention was part of the larger At Home/Chez Soi project, a multi-site field trial of HF. |
| **Offending** | Lamberti et al., 2017 | Forensic Assertive Community Treatment Model (FACT) | FACT is an adaptation of the assertive community treatment model specifically for justice-involved adults with severe mental illness. The model involves four components: high-fidelity ACT provided by a team of staff experienced with criminal justice system, identification and targeting of criminogenic risk factors, use of legal authority to promote engagement in necessary interventions, and mental health-criminal justice collaboration to promote effective problem solving. This FACT intervention was implemented for one year in the USA and |

| | | | |
|---|---|---|---|
| | | | aimed to reduce crime recidivism, reduce hospital use and promote engagement with outpatient mental health services. |
| **Social connectedness and social skills** | Ali et al., 2021 | Befriending | A one-to-one befriending intervention in the UK for people with intellectual disability and possible depression, aiming to provide social support and promote engagement in community activities. Participants were matched to a volunteer on the basis of shared interests and availability. The pairs had access to a resource booklet of local activities and were expected to meet once a week for about one hour, over 6 months, and to spend at least half their sessions in the community. Expenses were reimbursed. |
| **Trauma and victimisation** | De Waal et al., 2021 | Self, Otherwise, Streetwise (SOS) training | The SOS training intervention aimed to reduce victimisation in people in the Netherlands with co-occurring substance use disorders and mental ill-health. The intervention is a manualised group training, delivered in 12 twice-weekly 90-minute sessions. It comprises three modules (Self-wise, Other-wise, Streetwise), each consisting of 4 sessions. The Self module involves emotion regulation skills training, the Other-wise module involves conflict resolution skills training, and the Streetwise module involves street skills training. |

#### Table 5: Pooled statistics for inclusion of minority and majority groups on the basis of ethnicity, race, nationality, heritage, indigeneity and related factors

We aimed to broadly estimate the inclusion of individuals from groups which are marginalised on the basis of ethnicity, race, nationality, heritage, indigeneity or related factors in research. Where data on more than one of these indicators was available, we focussed on ethnicity. We categorised samples into "minority" or "majority" groups according to the demographics of the country the study was carried out in. If studies only reported inclusion of a majority group, we inferred the corresponding inclusion of participants from minority groups. Similarly, if the reported percentages were not complete, yet a majority group percentage was reported, we estimated % of minority group participants to be the remainder of participants.

Studies not reporting any relevant data were excluded from these calculations. In fact, only 53% of total studies included in the review reported this data. This is problematic for our estimates as data is likely to be missing not at random as studies not including any minority groups may be more likely to omit reporting the according demographics of their samples.

As such, the percentages we provide are based on estimates. This is a crude and limited strategy, necessitated by the poor reporting standards in research, and cannot be used to infer the experience of specific groups.

| Country | N studies reporting data;<br>Most common categorisation | Majority group pooled estimates (%) | Minority group pooled estimates (%) |
|---|---|---|---|
| **Australia** | 3<br>Nationality | Mean(SD) = 78(13)<br>Median=74<br>Min=68<br>Max=93<br>IQR=13 | Mean(SD) = 21(13)<br>Median=26<br>Min=7<br>Max=32<br>IQR=13 |
| **Canada** | 17<br>Ethnic or cultural origins | Mean(SD) = 60(25)<br>Median=60<br>Min=0<br>Max=100<br>IQR=36 | Mean(SD)=40(25)<br>Median=40<br>Min=0<br>Max=100<br>IQR=36 |
| **Columbia** | 1<br>Ethnicity | 93% | 7% |
| **China** | 2<br>Ethnicity | Mean (SD) =100(0)<br>Median=100<br>Min=100<br>Max=100<br>IQR=0 | Mean(SD) =0<br>Median=0<br>Min=0<br>Max=0<br>IQR=0 |
| **France** | 2<br>Nationality | Mean (SD) = 83 (2)<br>Median =83 | Mean(SD) = 17(2)<br>Median = 17 |

| | | | |
|---|---|---|---|
| | | Min=82<br>Max=85<br>IQR=2 | Min = 15<br>Max = 18<br>IQR = 2 |
| **Hong Kong** | 2<br>Ethnicity | Mean(SD) =100(0)<br>Median=100<br>Min=100<br>Max=100<br>IQR=0 | Mean(SD) =0<br>Median=0<br>Min=0<br>Max=0<br>IQR=0 |
| **Netherlands** | 8<br>Nationality | Mean (SD) = 68(21)<br>Median=69<br>Min=40<br>Max=98<br>IQR=31 | Mean (SD)=32(21)<br>Median=30<br>Min=2<br>Max=60<br>IQR=31 |
| **Sweden** | 1<br>Immigration status | 92% | 8% |
| **Switzerland** | 2<br>Nationality | Mean(SD) =64(15)<br>Median=64<br>Min=53<br>Max=74<br>IQR=11 | Mean(SD) =37(15)<br>Median=37<br>Min=26<br>Max=47<br>IQR=11 |
| **UK** | 14<br>Ethnicity | Mean(SD)=66(30)<br>Median=70<br>Min=0<br>Max=100<br>IQR=35 | Mean(SD)=34(30)<br>Median=30<br>Min=0<br>Max=100<br>IQR=35 |
| **USA** | 90<br>Ethnicity and Race | Mean(SD)=47(21)<br>Median=49<br>Min=10<br>Max=97<br>IQR=35 | Mean(SD)=53(21)<br>Median=51<br>Min=3<br>Max=90<br>IQR=35 |

#### Table 6: Stratified analyses results by paper

| Author (date) | Study design | Stratified analysis of socio-economic outcomes | Stratified effect |
|---|---|---|---|
| **Stratification: gender/sex** | | | |
| Castelein et al (2008) | RCT | Compared participants who improved at least 0.2 SD on overall quality of life at follow-up with the remaining participants. | No relevant statistics reported. Authors reported that gender did not predict overall quality of life. |
| Christensen et al., (2021) | RCT | Multiple regression analyses were conducted with interaction terms between study condition (IPS vs IPSE vs SAU)* and significant baseline predictors of competitive employment or education at follow-up.<br><br>*IPS = individual placement and support<br>IPSE = enhanced IPS with cognitive remediation and work-related social skills training | Interaction effects be tween gender(male) and study condition on vocational recovery:<br><br>IPS vs SAU: OR=1.17, CI=0.56-2.46, p=0.674<br><br>IPSE vs SAU: OR=0.57, CI=0.26-1.25, p=0.164<br><br>IPSA vs IPS: OR=0.49, CI=0.22-1.06, p=0.073 |
| de Waal et al (2019) | RCT | Logistic regression analyses fitted adjusted for predictors of treatment response for total victimisation, violent victimisation and property victimisation. | No relevant statistics reported for any victimisation outcome. Authors reported that gender did not predict treatment response for any victimisation outcome. |
| Dubreucq et al (2020) | Quasi-experimental | Multivariate analysis included gender as one of ten covariates to investigate improvements in Personal and Social Performance Scale (PSP) pre-post intervention (as well as 3 clinical variables). | No relevant effect size data reported. Authors reported that gender did not predict improvement in PSP ($p$=0.104). |
| Goldfinger et al (1999) | RCT | Multiple regression analysis included interaction between gender and housing type on number of days homeless. | No relevant data reported. Authors reported that effect of housing type on days homeless did not vary with gender. |

| | | | |
|---|---|---|---|
| Herman et al (2011) | RCT | Logistic regression models with homelessness during last three intervals as the outcome were repeated on men and women subgroups. | No data by gender reported. Authors reported that similar results were found for men and women. |
| Hui et al (2023) | RCT | The main analyses, linear mixed-effects models, were repeated within each sex subgroup to assess subgroup differences on the treatment x time interaction effect found in the main analyses on social functioning outcomes. | No data reported for this analysis. Authors reported that no subgroup differences on the treatment x time interaction were found for the sex subgroup. |
| Kidd et al (2021) | RCT | A linear model including an interaction effect between treatment group and gender on the Multnomah Community Ability Scale score was used. | No data reported for this analysis. Authors reported that gender was not a consistent moderator. |
| Martin-Carrasco et al (2016) | RCT | Subgroup analysis of the experimental group was conducted to identify if sex was associated with reduced caregiver burden. Reported results of associations among change scores since baseline of the measure of caregiver burden from multiple linear regression analysis where Male is the reference level for the sex variable. | Coefficient (95%CI) -<br>Female: 0.14 (-0.12-0.41) |
| McHugo et al (2004) | RCT | Mixed-effect regression models were used to evaluate treatment group differences over time with gender as a possible moderator variable. | Mean proportion of time spent in stable housing during each 6-month assessment period:<br>Integrated housing –<br>Females: mean=0.69, SD=0.36<br>Males: mean=0.70, SD=0.33;<br><br>Parallel housing –<br>Females: mean=0.70, SD=0.37<br>Males: mean = 0.40, SD=0.40<br><br>$F(1,107)=8.32$, $p=0.005$. |

| | | | |
|---|---|---|---|
| | | | No treatment group main effects were found in satisfaction with housing or victimisation so no data on gender interaction effects were available. |
| O'Campo et al (2023) | RCT | This paper comprised a post-hoc subgroup exploratory analysis of the impact of a Housing First intervention on housing stability, community functioning, psychological community integration and physical community integration in self-identified women using linear mixed models and a generalised estimating equation. | Mean percentage of days spent stably housed during follow-up:<br>TAU: 74.8 % (95%CI=71.7-77.8%), HF: 37.9% (95%CI= 34.4-41.3%),<br>*p*<0.001.<br><br>Odds of stable housing during follow-up: OR = 5.09, 95% CI = 4.08-6.35, p<0.001.<br><br>Mean change from baseline to 24 months in community functioning:<br>TAU=4.8 (95%CI = 3.6-6.0), HF=3.8 (95%CI= 2.8-4.9); *p*=0.236.<br><br>Mean change from baseline to 24 months in psychological community integration:<br>TAU= 2.0 (95%CI=1.4-2.6), HF=2.0 (95%CI=1.5-2.4), *p*=0.941.<br><br>Rate ratio of physical community integration in the past month:<br>TAU=1.03 (95%CI= 0.92-1.14), HF=0.97 (95%CI=0.86,1.08),<br>*p*=0.439. |
| Rebergen et al (2009) | RCT | Gender was considered as a potential effect modifier and tested as an interaction effect in a Cox proportional hazards model for return to work. | No data reported for this analysis. Authors report no modifying effect of gender. |
| Rossler et al (2020) | RCT | Gender was included in a multivariable Cox regression model to test potential moderator effect. | No data regarding this analysis were reported. Authors reported that there was no moderator effect of gender. |
| Scanlan et al (2019) | Uncontroll ed | Proportion achieving employment outcome and employment duration were reported for each gender, where between-group differences were assessed with | Proportion achieving employment outcome:<br>Male = 41.3% (n = 19/46)<br>Female = 52.6% (n= 29/51) |

| | | | |
|---|---|---|---|
| | prospective study | Chi-square analyses and *t*-tests for employment outcome and duration, respectively. | $\chi^2(1) = 2.34$, $p = 0.13$<br><br>Employment duration, mean (SD) days:<br>Male = 165 (144)<br>Female: 143 (169)<br>$t(45) = 0.46$, $p= 0.66$ |
| Swinkels et al (2023) | RCT | Linear Mixed Models with interaction terms between sex and treatment condition were conducted to explore the potential moderating effect of sex on outcomes including criminal recidivism, social network size and quality as well as social support and loneliness. | Criminal behaviour in intervention v TAU:<br>Male: RR =0.519, 95%CI = 0.203 - 1.330<br>Female: RR =13.885, 95%CI = 2.090 - 92.253 [i.e. negative effect]).<br><br>No other data on sex moderating effects reported. |
| **Stratification: ethnicity/race** | | | |
| Caplan et al (2023) | RCT | Analysis of qualitative data comparing reported progress in relationships with children in Housing First vs TAU conditions. | Proportion reporting improvements in relationships with children:<br>Indigenous parents -<br>HF = 62% (n=8/13); TAU = 13% (n=1/8).<br>Non-Indigenous parents-<br>HF = 36% (n=5/14); TAU= 25% (n=2/8). |
| Goldfinger et al (1999) | RCT | Multiple regression analysis included interaction between ethnicity and housing type on number of days homeless. | Mean ±SD days homeless during follow-up:<br>Independent living: African American or Hispanic = 107±26 vs White = 48±25<br><br>Staffed group housing: African American or Hispanic = 51±29 vs White = 36±32<br><br>B=0.22, $p<.01$ |
| Marder et al (1996) | RCT | General linear mixed model analysis was used to study interactions between ethnicity and psychosocial treatment modality. | No data on comparisons by ethnicity were reported. Authors reported that no interaction effect between ethnicity and psychosocial treatment modality on social adjustment was found. |
| **Stratification: socio-economic status indicators** | | | |

| | | | |
|---|---|---|---|
| Brown et al (2016) | Retrospective pre-post records analysis | Analyses of covariance were conducted for the homelessness outcome included an interaction of group (Housing First vs comparison) and pre-housing days homeless. | Group x pre-housing days homeless interaction:<br>*F*(1, 182)=0.77; Mean square = 14.28; *p*=0.38.<br>No further data regarding this comparison reported. |
| Christensen et al (2021) | RCT | Multiple regression analyses were conducted with interaction terms between study condition (IPS vs IPSE vs SAU)* and significant baseline predictors of competitive employment or education at follow-up.<br><br>*IPS = individual placement and support<br>IPSE = enhanced IPS with cognitive remediation and work-related social skills training | Work history was a stronger predictor of vocational recovery in the SAU group (those who had at least 2 month paid job in the last 5 years were 2.58 times more likely to work/study in the 18-month follow-up period) than in the IPS or IPSE groups (where they were 1.72 and 1.64 times more likely, respectively).<br><br>Interaction effects between previous work history and study condition on vocational recovery :<br>IPS vs SAU:<br>OR=0.67, CI=0.32-1.40, *p*=0.287<br><br>IPSE vs SAU:<br>OR=0.63, CI=0.30-1.35, *p*=0.236<br><br>IPSE vs IPS:<br>OR=0.95, CI=0.45-1.99, *p*=0.889 |
| De Waal et al (2019) | RCT | Logistic regression analyses fitted adjusted for predictors of treatment response for total victimisation, violent victimisation and property victimisation. | Proportion of participants achieving treatment response:<br>Middle/High education = 82.6%<br>Lower education = 67.3%<br>OR = 2.46, 95%CI=1.26-4.83, *p*=0.009.<br><br>No data reported with regards to treatment response on the basis of education for total victimisation or property victimisation. |
| Dubreucq et al (2020) | Quasi-experimen | Multivariate analysis included education as one of ten covariates to investigate improvements in Personal | No relevant effect size data reported. Authors reported that education did not predict improvement in PSP (*p*=0.946). |

| | | | |
|---|---|---|---|
| | tal controlled design | and Social Performance Scale (PSP) pre-post intervention (as well as 3 clinical variables). | |
| Elbogen et al (2016) | RCT | Post-hoc analyses examining the association between annual income and use of skills taught in the intervention condition. | Annual income was related to the number of $AFE skills used (b=.57, *p*=.02) and to the odds of using versus not using a $AFE budget (odds ratio [OR]=1.64, 95% confidence interval [CI]=1.16–2.32, *p*=.005). |
| Goldfinger et al (1999) | RCT | Multiple regression analysis included interaction between education and housing type on number of days homeless. | No data regarding this analysis reported. The authors reported that effect of housing type on days homeless did not vary with education. |
| Gutman et al (2009) | Quasi-experimental design | Examined correlations between success completing the program and enrolling in further employment or education, and educational level of participants or their parents. | No data regarding this analysis reported. The authors reported that there was no significant correlation between success and educational level of participants or their parents. |
| Marder et al (1996) | RCT | General linear mixed model analysis was used to study interactions between education and psychosocial treatment modality. | No data on outcome comparisons by education were reported. Authors report that there was no interaction effect between education and psychosocial treatment modality on social adjustment. |
| Martin-Carrasco et al (2016) | RCT | Multiple regression analysis of the experimental group data was conducted to identify if educational level was associated with reduced caregiver burden. | Coefficient (95%CI) -<br><br>Primary education: reference level<br>Secondary education: 1.79 (-4.12-7.70)<br>College, university: 2.79(-4.33-9.90) |
| Maru et al (2021) | RCT | Responder analyses were conducted on participants in the experimental condition to determine demographic differences in participants who achieved positive vocational outcomes a) for 6 months during the 12-month follow-up period, and b) had become employed at any time during the follow-up, versus those who did not. | No data regarding socioeconomic characteristics and 6-month employment outcomes as authors report no difference on the basis of significance testing.<br><br>Participants receiving disability benefits were less likely to be classified as a responder (employed at anytime during 12 month follow-up): 83% of non-responders vs 52% of responders, $\chi2(1)=8.54$, *p*=.004. |

| | | | |
|---|---|---|---|
| McHugo et al (2004) | RCT | Mixed-effect regression models were used to evaluate treatment group differences over time with lifetime homelessness as a possible moderator variable. | No associated data reported. Authors report that lifetime homelessness did not interact with the effect of group on stable housing. |

#### Studies exclusively including participants with psychosis spectrum disorder diagnoses - Results

Seven of the 72 studies including exclusively participants with psychosis reported conducting stratified analyses according to gender, socioeconomic status or ethnicity/race of participants (see * symbol, Table 1). These studies were all RCTs except one which followed a quasi-experimental design (Dubreuqc et al., 2020). The majority of the RCTs were two-armed, and two RCTs were three-armed (n=110; n=360). Sample sizes ranged from 80 to 360 participants (median = 110, mean = 159).

Six of these studies stratified results on the basis of gender (Castelein et al., 2008; Dubreucq et al., 2020; Herman et al., 2011; Hui et al., 2023; Kidd et al., 2021; Martín-Carrasco et al., 2016) however only one of these reported relevant effect size data pertaining to this analysis (Martín-Carrasco et al., 2016). This two-arm RCT study found that a psychoeducational intervention to reduce caregiver burden may be more effective for women than men, as being female was moderately associated with burden reduction; however, data were statistically consistent with parameter values ranging from a considerable level of reduced risk to a considerable level of increased risk of burden reduction.

One of these studies stratified results on the basis of ethnicity regarding a social skills intervention. However, they did not report sufficient data concerning how changes in social adjustment varied between different ethnic groups (Marder et al., 1996).

Three of these studies performed stratified analyses on the basis of socio-economic indicators (Dubreucq et al., 2020; Marder et al., 1996; Martín-Carrasco et al., 2016). Again, Martin-Carrasco et al (2016) was the only study to report relevant effect size data concerning this analysis, finding a large effect size indicating that being college/university- or secondary school-educated was associated with greater response to the psychoeducational carer intervention (compared to having a primary school education alone), however data were statistically consistent with parameter values ranging from a considerable level of reduced risk to a considerable level of increased risk of burden reduction.

#### Studies exclusively including participants with psychosis spectrum disorder diagnoses - Discussion

No studies that exclusively included participants with psychosis spectrum diagnoses conclusively found evidence for differential treatment effects based on gender, socioeconomic status, or ethnicity of participants. The research in this area was extremely limited as only seven studies included such an analysis. This may in part be due to small samples limiting the power to run stratified analyses. Only one of these reported relevant effect size data to interpret the results of their stratified analysis, finding that women may benefit more than men from a care burden reduction intervention, however these results were not conclusive. Despite the increased social adversity faced by people with psychosis from minoritised Black ethnicities, only one of the studies recruiting participants with psychosis investigated ethnicity as a moderator of intervention effects, and did not report effect size or related data regarding this analysis.

#### Replicated Killaspy et al., (2022) Search Strings for January 2020-February 2024

Ovid MEDLINE(R) ALL <1946 to February 26, 2024>

1 *Mental Disorders/ 143705

2 exp Mentally Ill Persons/ 6451

3 "Diagnosis, Dual (Psychiatry)"/ 3742

4 ("mental illness*" or "mental distress" or "mental disturbance*" or "mental disorder*" or "mentally ill").tw. 97826

5 or/1-4 212155

6 Psychotic Disorders/ 53383

7 Schizophrenia/ 112566

8 Affective Disorders, Psychotic/ 2319

9 Bipolar Disorder/ 45856

10 Psychoses, Substance-Induced/ 5392

11 (psychotic or psychosis or psychoses or schizophrenia or bipolar or "schizoaffective affective disorder*" or "delusional disorder*").tw. 239531

12 ("complex trauma" or "serious mental illness*").tw. 5471

13 or/6-12 297478

14 (severe or chronic or enduring or persistent or persisting or complex or "serious and continuing").tw. 4194181

15 ("high level" and (disabilit* or need*)).tw. 16968

16 ("psychosocial disabilit*" or "psycho-social disabilit*").tw. 384

17 or/14-16 4208099

18 5 and 17 35490

19 13 or 18 322714

20 exp Mental Health Services/ 106854

21 exp Community Psychiatry/ 2091

22 community health services/ or exp community mental health services/ or patient participation/ 81603

23 Residential Facilities/ 5760

24 Residential Treatment/ 3345

25 Health Services for Persons with Disabilities/ 149

26 Preventive Health Services/ 14514

27 "inpatient mental health".mp. 698

28 "psychological service*".mp. 1132

29 "Delivery of Health Care"/ 121510

30 "Delivery of Health Care, Integrated"/ 14434

31 Patient Care Management/ 4749

32 Program Evaluation/ 67493

33 "Continuity of Patient Care"/ 20811

34 "Referral and Consultation"/ 77161

35 "model* of care".tw. or "care model*".mp. 21391

36 "service delivery model*".tw. 1557

37 institutionali*.tw. 18419

38 de-institutionali*.tw. 178

39 transition*.tw. 535805

40 discharge.tw. 250147

41 "stepped care".tw. 1711
42 "shared care".tw. 1641
43 "collaborative care".tw. 3129
44 "community care".tw. 5523
45 "integrated care".tw. 6568
46 "integrated model*".tw. 4447
47 "case management".tw. 12712
48 Rehabilitation/ 18716
49 ("psychosocial rehabilitation" or "psycho-social rehabilitation" or recovery-based or "recovery based" or recovery-orient* or "recovery orient*" or "psychosocial support*" or "psycho-social support*").tw. 8369
50 Home Care Services/ 36707
51 ("home based outreach" or "outreach service*" or "assertive community treatment").tw. 1862
52 social support/ 79644
53 Employment, Supported/ 1380
54 ("supported employment" or "individual placement and support" or "competitive employment" or "peer supported work" or "peer workforce" or "vocational rehabilitation" or "employment assistance" or "supported education").tw. 3902
55 ("volunteering" or "voluntary work").tw. 2952
56 Housing/ 20455
57 ("support* housing" or "housing first" or "staffed housing" or "residential support" or "staffed residen*" or "24 hour supported community" or "twenty-four hour supported community" or "residen* rehabilitation" or "support* accommodation").tw. 1783
58 Occupational Health Services/ 10771
59 "occupation based lifestyle intervention*".tw. 4
60 "occupational time use intervention*".tw. 1
61 "Activity based intervention*".tw. 146
62 ((remotivation or re-motivation) adj2 process*).tw. 2
63 Family/ 85586
64 "family involvement".kw. 135
65 ("family psycho-education" or "family psychoeducation" or "family education" or "multiple family groups" or "family involvement" or "family support").tw. 11153
66 Cognitive Behavioral Therapy/ 31001
67 Psychotherapy/ 58435
68 Behavior Therapy/ 30772
69 ("cognitive behavioral therapy" or "cognitive behavioural therapy" or CBT or "cognitive remediation").tw. 25574
70 Self Care/ 36385
71 (psychoeducation or "psycho-education" or "self-management" or "self management" or "personal* support*" or "person centered" or "person centred" or "self-help group" or "personal* assistance" or "peer support*" or "peer group" or "peer led" or "peer-let" or "care co-ordinators" or care coordinators).tw. 54352
72 Social Skills/ 2768
73 Social Adjustment/ 23707
74 ("social skills" or "social functioning" or "social network" or "social skills training" or "social cognition training" or "skills training").tw. 40572
75 ("social connection" or "social support*").tw. 56723
76 "social interventions".tw. 935
77 "Social prescribing".tw. 382
78 social participation/ or "Social participation".tw. 6774

79 Harm Reduction/ 4280
80 ("harm reduction" or "motivational interviewing").tw. 12259
81 ("early intervention" or "priority intervention" or "mental health triage" or "illness management" or "relapse prevention" or "wellness recovery action plan" or ("wellness plan*" or "safety plan*" or "recovery plan*")).tw. 30791
82 "shared decision making".tw. 14095
83 partnership*.tw. 48776
84 collaborative.tw. 77325
85 "dispersed power".tw,kw. 3
86 "trauma informed".mp. 3041
87 "group work".tw. 1382
88 "evidence informed intervention*".mp. 139
89 "evidence based intervention*".mp. 7226
90 ("psycho-social intervention*" or "psychosocial intervention*").mp. 8739
91 (NDIS or "national disability insurance scheme").mp. 446
92 ("step-down" or secure).tw. 38039
93 ("co-design" or codesign or "co-production" or coproduction).tw. 6097
94 ("open dialogue" or "open dialog").tw. 632
95 ("needs based model*" or "needs-based model").tw. 48
96 "network* model*".tw. 19773
97 or/20-96 1934518
98 Quality of Life/ or "quality of life".tw. 461603
99 Patient Readmission/ or Patient Admission/ or readmission.tw. or readmit*.tw. or admission.tw. or admit*.tw. 508452
100 Activities of Daily Living/ or activities of daily living.tw. 89239
101 Homeless Persons/ or homeless.tw. 13800
102 Occupations/ or Return to Work/ or vocation.tw. or return to work.tw. or return-to-work.tw. or back to work.tw. or back-to-work.tw. or "work participation".tw. or "vocation* outcome*".mp. 39889
103 Health Behavior/ or Risk Reduction Behavior/ or Life Style/ or exercise.tw. or tobacco.tw. or diet.tw. 920637
104 Patient Satisfaction/ or "patient satisfaction".tw. or "consumer satisfaction".tw. or "consumer recovery".tw. or "consumer outcome*".tw. 116784
105 "Carer satisfaction".tw. 98
106 (family relationship* or family connection*).tw. 5268
107 "restrictive intervention*".tw. 162
108 seclusion.tw. 1594
109 restraint*.tw. 29920
110 ("forced medication" or "forced sedation" or coercion).tw. 4291
111 (compulsion or "restrictive practice*" or adherence).tw. 165262
112 (recovery adj3 measur*).mp. 6205
113 connection.tw. 110744
114 inclusion.tw. 347253
115 "consumer experience of care".tw. 1
116 "family experience of care".tw. 21
117 (Choice or Connectedness or Hope or Optimism or Meaning or empower* or Agency or Autonomy or Growth or Wellbeing or well-being or Belonging).tw. 2606185
118 qualitative.tw. 335760
119 "mixed method*".tw. 45479
120 evidence based practice/ 12203
121 ("consumer led" or "consumer academic" or "lived experience academic").tw. 127

122 ("stigma reduction" or "stigma and discrimination").tw. 3673
123 mutuality.tw. 899
124 "personal journey".tw. 298
125 isolation.tw. 299723
126 carer.tw. 6258
127 supporters.tw. 3225
128 "significant other*".tw. 4922
129 engagement.tw.107402
130 "relational recovery".tw. 4
131 ("therapeutic alliance*" or "therapeutic relationship*").tw. 7348
132 or/98-131 5529052
133 19 and 97 and 132 19990
134 19 and 91 12
135 133 or 134 19995
136 limit 135 to (english language and yr="2020 -Current") 4247
137 19 and 97 51754
138 limit 137 to randomized controlled trial 3192
139 Randomized Controlled Trial/ 609629
140 Randomized Controlled Trials as Topic/ 167234
141 ("randomi#ed controlled trial*" or RCT or "randomi#ed controlled study*" or "randomi#ed trial*or double blind*").mp. 900102
142 139 or 140 or 141 900102
143 137 and 142 4995
144 138 or 143 4995
145 limit 137 to (meta analysis or "systematic review") 1567
146 Meta-Analysis/ 195913
147 Meta-Analysis as Topic/ 23784
148 ("meta analys*" or metaanalys* or "systematic review*").mp. 503098
149 146 or 147 or 148 503098
150 137 and 149 2239
151 145 or 150 2239
152 144 or 151 6539
153 152 not 135 3009
154 limit 153 to (english language and yr="2020 -Current") 679

#### Replicated Barnett et al., (2022) Search Strings for January 2020-February 2024

**APA PsycInfo** <1806 to February Week 4 2024>

1 clinical trials.sh. 12305

2 (randomi#ed or randomi#ation or randomi#ing).ti,ab,id. 113208

3 (RCT or at random or (random* adj3 (administ* or allocat* or assign* or class* or control* or crossover or cross-over or determine* or divide* or division or distribut* or expose* or fashion or number* or place* or recruit* or split or subsitut* or treat*))).ti,ab,id. 130161

4 ((control* adj5 (trial or study or group?)) and (placebo or waitlist* or wait* list* or ((treatment or care) adj2 usual))).ti,ab,id,hw. 26911

5 trial.ti,id. 45596

6 controlled trial.ab. 26277

7 or/1-6 190648

8 ((children? or kids or adolescen* or teens or teenagers or schools or school based) not (adult* or family or families or mother? or woman* or women* or female? or father? or men or mens or male? or relations or former)).ti. 364986

9 prevalence.ti. 23709

10 7 not (8 or 9) 175755

11 limit 10 to yr="2020 -Current" 35795

12 BIPOLAR DISORDER/ or bipolar i disorder/ or bipolar ii disorder/ or cyclothymic disorder/ or mania/ or hypomania/ or exp dissociative disorders/ or PERSONALITY DISORDERS/ or antisocial personality disorder/ or avoidant personality disorder/ or borderline personality disorder/ or dependent personality disorder/ or histrionic personality disorder/ or narcissistic personality disorder/ or obsessive compulsive personality disorder/ or paranoid personality disorder/ or passive aggressive personality disorder/ or schizoid personality disorder/ or schizotypal personality disorder/ or PSYCHOSIS/ or exp acute psychosis/ or affective psychosis/ or exp alcoholic psychosis/ or capgras syndrome/ or chronic psychosis/ or postpartum psychosis/ or reactive psychosis/ or schizophrenia/ or acute schizophrenia/ or catatonic schizophrenia/ or paranoid schizophrenia/ or process schizophrenia/ or schizoaffective disorder/ or schizophreniform disorder/ or undifferentiated schizophrenia/ or delusions/ or schizotypy/ or toxic psychoses/ 193544

13 (bipolar or cyclothymi* or mania or manic or hypermani* or rapid cycling or conversion disorder* or (dissociative adj (amnesi* or fugue* or disorder*)) or borderline state? or catatoni* or character disorder* or delusion* or capgras syndrom* or diogenes syndrom* or depersonalization or depersonalisation or de-personalization or de-personalisation or perceptual disorder* or personality disorder* or BPD or paranoi* or psychiatr* or psychopathol* or psycho-pathol* or psychotic* or psychosis* or psychoses* or schizo* or hebephreni* or serious* mental* or SMI).ti,ab,id. 548961

14 12 or 13 556617

15 11 and 14 4199

16 MENTAL DISORDERS/ or anhedonia/ or neurosis/ or ACUTE STRESS DISORDER/ or adjustment disorders/ or attachment disorders/ or disinhibited social engagement disorder/ or posttraumatic stress disorder/ or complex ptsd/ or desnos/ or acute stress disorder/ or post-traumatic stress/ or traumatic neurosis/ or *emotional trauma/ or AFFECTIVE DISORDERS/ or disruptive mood dysregulation disorder/ or dysthymic disorder/ or seasonal affective disorder/ or major depression/ or anaclitic depression/ or endogenous depression/ or late life depression/ or postpartum depression/ or reactive depression/ or recurrent depression/ or treatment resistant depression/ or premenstrual dysphoric disorder/ or ANXIETY DISORDERS/ or generalized anxiety disorder/ or exp obsessive compulsive disorder/ or panic attack/ or panic disorder/ or exp phobias/ or trichotillomania/ or phobias/ or acrophobia/ or agoraphobia/ or claustrophobia/ or ophidiophobia/ or social phobia/ or separation anxiety disorder/ or mutism/ or elective mutism/ or EATING

DISORDERS/ or anorexia nervosa/ or binge eating disorder/ or bulimia/ or SELF-INJURIOUS BEHAVIOR/ or self-destructive behavior/ or self-inflicted wounds/ or self-mutilation/ or self-poisoning/ or suicide/ or attempted suicide/ or suicidality/ or suicidal ideation/ or suicide prevention/ or SOMATOFORM DISORDERS/ or body dysmorphic disorder/ or exp conversion disorder/ or exp factitious disorders/ or hypochondriasis/ or exp hysteria/ or neurasthenia/ or neurodermatitis/ or somatization disorder/ or somatoform pain disorder/ or munchausen syndrome/ or munchausen syndrome by proxy/ 459895

17 "Depression (Emotion)"/ 27158

18 "Stress and trauma related disorders"/ 56

19 Mental Health/ or (mental* adj2 (health* or ill*)).ti,ab,id. 300112

20 Mental Health Services/ or Community Mental Health Services/ or Community Counseling/ or Community Psychiatry/ 48950

21 Mental Health Program Evaluation/ 2457

22 (acute stress or adjustment disorder* or ADNOS or affective disorder* or agoraphobi* or anorexia nervosa or anxiety or astheni* or attachment disorder* or binge eat* or binging or body dysmorphi* or bulimi* or combat disorder* or obsessive or compulsi* or OCD or depression or depressed or depressive or dyssomni* or dyspareunia* or dysphori* or dysthymi* or dystoni* or eating disorder* or EDNOS or emotional trauma or fear or health anxiety or hoarding or hyperactivity or hypochondri* or hysteri* or medically unexplained or malingering or MDD or common mental or (mental* adj2 (health or well*)) or mood or moods or munchausen or MUPS or mutism or neurastheni* or neurotic or neuros* or panic or phobi* or PND or ((post-trauma* or posttrauma*) adj stress*) or psychogenic or psychosomatic or PTSD or (self adj (injur* or harm or mutilat*)) or psychosexual or (psychological adj3 sexual adj3 dysfunction*) or social* anxi* or somati* or somatoform or suicid* or parasuicid* or trichotillomani*).ti,ab,id. 1065011

23 or/16-22 1146616

24 11 and 23 13736

25 social prescribing.mp. 134

26 ((chang* or develop* or enhanc* or initiative? or intervention? or program* or mitigat* or address* or improv* or target*) adj3 (community or living or social) adj3 (condition? or circumstance?)).ti,ab,id. 1498

27 ((communit* or social) adj (connect* or engagement? or link* or referral? or intervention? or wellbeing)).ti,ab,id. 15946

28 "sense of belonging".ti,ab,id. 4477

29 or/26-28 21709

30 15 and 29 26

31 24 and 29 135

32 housing/ or assisted living/ or group homes/ or shelters/ 10482

33 residential facilit*.ti,id. 772

34 homeless/ or homeless mentally ill/ or deinstitutionalization/ 11259

35 ((chang* or develop* or enhanc* or initiative? or intervention? or program* or mitigat* or address* or improv* or target*) adj3 (housing or neighbo?rhood?)).ti,ab,id. 4789

36 homeless*.ti,ab,id. 13421

37 ((housing adj (first or stability or instability)) or permanent housing).ti,ab,id. 1283

38 housing.ti. or ((housing adj (strateg* or polic* or project* or program* or quality)) or new* buil* or social housing*).ti,ab,id. 5863

39 independent living programs/ or living arrangements/ or residential care institutions/ 15199

40 halfway houses/ or independent living programs/ or living arrangements/ or residential care institutions/ 15507

41 poverty areas/ or social environments/ 9810

42 ((autonomous or assisted or sheltered or support*) adj3 (housing or accommodation or dwelling?)).ti,ab,id. 2363
43 (((clubhouse or club house) adj model?) or ((autonomous or independent or assisted) adj living)).ti,ab,id. 5114
44 (community residences or group homes or community living or supervised apartments).ti,ab,id. 3471
45 therapeutic social clubs/ 157
46 ((independ* or assist* or support* or secur* or sustain* or maint*) adj3 (tenanc* or tenure?)).mp. [mp=title, abstract, heading word, table of contents, key concepts, original title, tests & measures, mesh word] 248
47 ((halfway or satellite) adj (dwelling? or home? or house?)).ti,ab,id. 500
48 (neighbo?rhood? adj (characteristic* or intervention* or program*)).ti,ab,id. 1460
49 ((environment* or housing or neighbo?rhood?) and infrastructure).ti,ab,id. 2510
50 built environment/ or urban planning/ 2275
51 or/32-50 64561
52 15 and 51 48
53 24 and 51 159
54 MONEY.ti,id. 4028
55 socioeconomic status/ or "income (economic)"/ or budgets/ or economic security/ or financial strain/ 39984
56 exp employee benefits/ 6927
57 *disadvantaged/ or *social deprivation/ 7035
58 ((access* or improv* or manag* or supplement*) adj2 (cash or money or financ* or income? or savings)).ti,ab,id. 4775
59 exp income level/ 20363
60 ((financial adj (autonomy or security or insecurity)) or loans or borrowing or budgeting or microcredit or microfinance or social fund*).ti,ab,id. 5378
61 high poverty.ti,ab,id. or poverty.ti. 6309
62 ((address* or escap* or improv* or support* or target*) adj2 (depriv* or poor or poverty)).ti,ab,id. 2624
63 "out of poverty".ab. 223
64 (((food or fuel) adj poverty) or food bank?).ti,ab,id. 189
65 ((alleviat* or ease or manag* or prevent* or reduc* or stop*) adj2 (poverty or ((economic or financial) adj hardship?))).ti,ab,id. 1637
66 ((alleviat* or eas* or manag* or prevent* or reduc* or relief or stop*) adj1 debt?).ti,ab,id. 115
67 debt?.ti,id. 1141
68 (austerity or recession?).ti,ab,id.3723
69 (((basic or minimum) adj3 (wage? or income?)) or zero hours).ti,ab,id. 705
70 paid work.ti,ab,id. 1813
71 "dealing with money".ab. 17
72 (family adj (income? or tax credit?)).ti,ab,id. 4661
73 "welfare services (government)"/ or community welfare services/ or medicaid/ or welfare reform/ 9000
74 welfare benefit?.ti,ab,id. 484
75 or/54-74 106448
76 15 and 75 48
77 24 and 75 213
78 employment status/ or employability/ or occupational tenure/ or occupational status/ or job security/ or job search/ or supported employment/ or vocational rehabilitation/ or vocational evaluation/ or work adjustment training/ or sheltered workshops/ 33919

79 unemployment/ or personnel termination/ or employee layoffs/ 6046
80 ((chang* or develop* or enhanc* or initiative? or intervention? or program* or address* or improv* or target*) adj3 (employment or unemployment or unemploy*)).ti,ab,id. 5738
81 (support* adj3 (employment? or work or vocational or occupation*)).ti,ab,id. 13943
82 ((job? or work* or employment* or employee? or occupation*) adj5 skills adj5 train*).ti,ab,id. 1373
83 (paid adj (job? or employment or work or occupation*)).ti,ab,id. 3133
84 (employment or unemployment or occupation*).ti. 32126
85 individual placement?.ti,ab,id. 479
86 ((finding or gaining or obtaining or keeping or sustaining) adj3 (work or job? or employment or occupation*)).ti,ab,id. 3165
87 (social firms or (sheltered adj (employment or work or occupation*))).ti,ab,id. 241
88 (precar* adj1 (job? or employment or work or occupation*)).ti,ab,id. 539
89 (voluntary work or volunteering).ti,ab,id. 3871
90 (meaningful adj (activit* or job? or employment or work or occupation*)).ti,ab,id. 2364
91 (((return or back) adj2 work) or absenteeism).ti,ab,id. 8541
92 ((alleviat* or ease or manag* or prevent* or reduc* or stop*) adj ((employment or work* or occupation*) adj disabilit*)).ti,ab,id. 47
93 (return* adj2 education).ab. 207
94 ((education or learning or training) adj3 (access* or takeup or take up)).ti,ab,id. 5567
95 ((labo?r force or employment or unemployment or occupation*) adj status).ti,ab,id. 9823
96 or/78-95 99361
97 15 and 96 86
98 limit 97 to yr="2020 -Current" 86
99 24 and 96 247
100 family relations/ or intergenerational relations/ or exp marital relations/ 59388
101 family conflict/ or marital conflict/ 6855
102 home environment/ or living alone/ 12193
103 ((family or families or intergenerat* or inter-generat*) adj (relation* or conflict?)).ti,ab,id. 22811
104 ((sexual or intimate or partner? or marital) adj (relation* or conflict?)).ti,ab,id. 21458
105 ((develop* or enhanc* or initiative? or intervention? or program* or address* or improv* or promot* or target*) adj2 relationship?).ti,ab,id. 21676
106 ((carer? or partner or relationship? or marital) adj support*).ti,ab,id. 1892
107 (child* adj2 (access or contact or custody or maintenance)).ti,ab,id. 5865
108 child custody/ or joint custody/ or child visitation/ or divorce/ or family reunification/ or living arrangements/ 15108
109 ((care proceeding? or family court? or child removal or fostercare or foster care) and (parent* or mother? or father?)).ti,ab,id. 4146
110 parenting/ or parental involvement/ or parental role/ 29491
111 *parents/ or parent* outcome?.ti,ab,id. 29595
112 or/100-111 188026
113 15 and 112 74
114 24 and 112 457
115 parent* mental health.ti,ab,id. 1359
116 114 or 115 1793
117 (VICTIMIZATION or victimisation or revictimi#ation).ti,ab,id,hw. 36020
118 crime victims/ 5792
119 (crime victim? or revictimi*).ti,ab,id. 2263

120 ((victim* or crime?) and survivor*).ti,ab,id,hw. 5794
121 domestic violence/ or battered females/ or exposure to violence/ or intimate partner violence/ or physical abuse/ or exp sexual abuse/ or shelters/ 54588
122 elder abuse/ 2066
123 ((domestic or partner? or spouse?) adj3 (abus* or violen*)).ti,ab,id. 27276
124 ((domestic or marital or partner? or spous*) adj3 (rape or sex* assault*)).ti,ab,id,hw. 819
125 (intimate partner adj2 abus*).ti,ab,id. 760
126 interpersonal control/ or coercion/ 15642
127 coercive control.ti,ab,id.520
128 ((female? or women?) adj (refuge? or shelter?)).ti,ab,id. 401
129 (exploitation or safe guarding or safeguarding).ti,ab,id. 10155
130 slavery/ or human trafficking/ or *freedom/ 4248
131 or/117-130 118723
132 exp *criminal offenders/ 17329
133 Recidivism.ti,id,sh. 7740
134 ((crime? or criminal* or offend* or offence? or recidiv*) adj3 (initiative? or intervention? or program* or mitigat* or address* or rehabilitat*)).ti,ab,id. 6364
135 ((crime? or criminal* or offend* or offence? or recidiv*) adj3 (diver* or prevent*)).ti,ab,id. 3999
136 prisoners/ or criminal rehabilitation/ or reintegration/ 16469
137 ((inmate? or prison* or convict? or felon? or gang member? or delinquent? or justice-involved or perpetrator? or probation) adj5 (release or integration or reintegrate or re-integrat* or rehabilitat* or re-habilitat* or desistance)).ti,ab,id. 2588
138 (community adj2 (reentry or re-entry or rehabilitat* or re-habilitat*)).ti,ab,id. 1758
139 or/132-138 44392
140 15 and (131 or 139) 85
141 24 and (131 or 139) 313
142 human rights/ or exp civil rights/ or exp freedom/ 41054
143 (((citizen? or civil* or human or legal or social or voting) adj rights) or social justice or equal protection or social protection).ti,ab,id. 27995
144 *needs/ 5787
145 rights.ti,hw. 16424
146 *government policy making/ 16885
147 ((public or social) adj polic*).ti,ab,id. 22128
148 equity-focus*.ti,ab,id. 159
149 *health disparities/ 9025
150 ((social or community or neighbo?rhood?) adj3 (equit* or inequit* or inequalit* or dispar*)).ti,ab,id. 9401
151 digital divide/ or information literacy/ 1169
152 internet access.ti,ab,id. 1331
153 (digital adj (inclusion or exclusion or divide or disparit* or equit* or inequit* or inequalit*)).ti,ab,id. 1686
154 or/142-153 120795
155 15 and 154 34
156 24 and 154 109
157 SOCIAL ISOLATION/ 9426
158 loneliness/ 7323
159 (loneliness or lonely).ti,ab,id. 15834
160 social isolation.ti,ab,id. 10712
161 ((social* or societ* or communit*) adj3 (isolated or isolation)).ti,ab,id. 13730

162 ((alleviat* or ease or manag* or mitigat* or prevent* or overcom* or reduc* or stop*) adj2 (isolation or isolated)).ti,ab,id. 1191
163 ((address* or enhanc* or improv* or increas* or promot* or target*) adj2 (inclusion or inclusivity)).ti,ab,id. 2026
164 or/157-163 34961
165 limit 164 to yr="2020 -Current" 9958
166 15 and 165 43
167 24 and 165 232
168 SOCIAL PARTICIPATION.mp. 4343
169 (social alienation or social inclusion).ti,ab,id. 3432
170 (community involvement or social support or social network or psychosocial environment or psychosocial rehabilitation).ti,id,hw. 66491
171 abandonment/ or alienation/ 3554
172 (abandonment or alienation).ti,id. 3797
173 ((social or societ* or community) adj (confine* or contact or contacts or connect* or inclusion or network* or participation or relations*)).ti,id,hw. 42232
174 ((social* or societ* or communit*) adj3 (alienat* or discriminat* or excluded or exclusion)).ti,ab,id,hw. 13385
175 ((alleviat* or ease or manag* or mitigat* or prevent* or overcom* or reduc* or stop*) adj2 (exclusion or excluded)).ti,ab,id. 323
176 (social capital or social* mobil*).ti,ab,id,hw. 14138
177 (navigator or navigators).ti,id. 342
178 peer? support*.ti,id. 2517
179 Peers/ and Social Support/ 1456
180 (anti-stigma* or ((intervention* or alleviat* or prevent* or reduc* or stop*) adj2 stigma*)).ti,ab,id. 5062
181 (social learning theory or (social adj3 interaction* adj3 (counsel* or educat* or intervention* or program* or therap* or train*)) or SCIT).ti,ab,id. 4159
182 community integration.mp. 1662
183 (peer? adj (support* or navigat*)).ti,id. 2550
184 community involvement/ or *social support/ or *social networks/ 51263
185 ((social or societ* or psychosocial) adj support*).ti,id,hw. 49099
186 ((social* or societ* or communit*) adj network*).ti,id,hw. 29176
187 ((address* or enhanc* or improv* or increas* or promot* or target*) adj3 (social* or communit*) adj3 (network? or support)).ti,ab,id. 4903
188 Psychosocial Rehabilitation/ 4760
189 ((address* or enhanc* or improv* or increas* or promot* or target*) adj2 (inclusion or inclusivity)).ti,ab,id. 2026
190 or/168-189 143458
191 15 and 190 193
192 24 and 190 630
193 30 or 52 or 76 or 98 or 113 or 140 or 155 or 166 or 191 520
194 31 or 53 or 77 or 99 or 116 or 141 or 156 or 167 or 192 3353
195 193 or 194 3489

### Web of Science Search Strategy (v0.1)

-

### Database: SciELO Citation Index

### Entitlements:
- SCIELO.SCIELO: 2002 to 2024

### Searches:
1: (TI=(trial) OR AB=(trial) OR TI=(randomized or randomised) OR AB=(”randomized controlled trial” or “randomised controlled trial” or RCT) or AB=(random* SAME (administ* or allocat* or assign* or class* or control* or crossover or cross-over or determine* or divide* or division or distribut* or expose* or fashion or number* or place* or recruit* or split or subsitut* or treat*) ))
Timespan: 2020-07-01 to 2024-02-28 Date Run: Wed Feb 28 2024 11:02:19 GMT+0000 (Greenwich Mean Time) Results: 10264

2: TI=(children or boys or girls or kids or adolescen* or teen* or school or schools)
Timespan: 2020-07-01 to 2024-02-28 Date Run: Wed Feb 28 2024 11:02:54 GMT+0000 (Greenwich Mean Time) Results: 10638

3: (#1 not #2) Timespan: 2020-07-01 to 2024-02-28 Date Run: Wed Feb 28 2024 11:03:16 GMT+0000 (Greenwich Mean Time) Results: 9901

4: (TS=(”serious* mental*” or schizo*) or TI=(bipolar or cyclothymi* or mania or manic or hypermani* or “rapid cycling” or “conversion disorder*” or “dissociative amnesi*” or “dissociative fugue*” or “dissociative disorder*” or “borderline state*” or catatoni* or “character disorder*” or delusion* or “capgras syndrom*” or “diogenes syndrom*” or depersonalization or depersonalisation or de-personalization or de-personalisation or “perceptual disorder*” or “personality disorder*” or BPD or paranoi* or psychiatr* or psychopathol* or psycho-pathol* or psychotic* or psychosis* or psychoses* or hebephreni* or SMI)) Timespan: 2020-07-01 to 2024-02-28
Date Run: Wed Feb 28 2024 11:03:37 GMT+0000 (Greenwich Mean Time)
Results: 990

5: (#4 AND #3) Timespan: 2020-07-01 to 2024-02-28 Date Run: Wed Feb 28 2024 11:03:53 GMT+0000 (Greenwich Mean Time) Results: 49

6: (TS=(“mental health” or “mental* ill*” or “common mental” or “mental disorder*” or psychiatric) or TI=(“acute stress” or “adjustment disorder*” or ADNOS or “affective disorder*” or agoraphobi* or “anorexia nervosa” or anxiety or astheni* or “attachment disorder*” or “binge eat*” or binging or “body dysmorphi*” or bulimi* or “combat disorder*” or “obsessive or compulsi*” or OCD or depression or depressed or depressive or dyssomni* or dyspareunia* or dysphori* or dysthymi* or dystoni* or “eating disorder*” or EDNOS or “emotional trauma” or fear or “health anxiety” or hoarding or hyperactivity or hypochondri* or hysteri* or “medically unexplained” or malingering or MDD or mental* or mood or moods or munchausen or MUPS or mutism or neurastheni* or neurotic or neuros* or panic or phobi* or PND or “post-traumatic stress” or “posttraumatic stress*” or psychogenic or psychosocial* or psychosomatic or PTSD or “self injur*” or “self harm” or “self mutilat*” or psychosexual or (psychological and sexual and dysfunction*) or “social anxi*” or “socially anxi*” or somati* or somatoform or suicid* or parasuicid* or trichotillomani*))
Timespan: 2020-07-01 to 2024-02-28 Date Run: Wed Feb 28 2024 11:04:29 GMT+0000 (Greenwich Mean Time) Results: 6784

7: (#6 AND #3) Timespan: 2020-07-01 to 2024-02-28 Date Run: Wed Feb 28 2024 11:04:50 GMT+0000 (Greenwich Mean Time) Results: 283

8: ((TS=(“social prescribing") ) OR (TS=((chang* or develop* or enhanc* or initiative* or intervention* or program* or mitigat* or address* or improv* or target*) SAME (“community condition*” or “conditions in the community” OR “living condition*” or “social condition*” OR “community circumstance**” or “living circumstance*” or “social circumstance*”) )) OR (TS=("communit* connect*" or "community engagement*" or "community link*" or "community referral*" or "community intervention*" or "community wellbeing” or "social connect*" or "social engagement*" or "social link*" or "social referral*" or "social intervention*" or "social wellbeing” or "social and emotional wellbeing”) )) Timespan: 2020-07-01 to 2024-02-28 Date Run: Wed Feb 28 2024 11:05:09 GMT+0000 (Greenwich Mean Time) Results: 714

9: (#8 AND #5) Timespan: 2020-07-01 to 2024-02-28 Date Run: Wed Feb 28 2024 11:05:29 GMT+0000 (Greenwich Mean Time) Results: 0

10: (#8 AND #7) Timespan: 2020-07-01 to 2024-02-28 Date Run: Wed Feb 28 2024 11:06:54 GMT+0000 (Greenwich Mean Time) Results: 1

11: ((TI=(housing or dwelling* or neighbourhood* or neighborhood* or “residential facilit*” or “independent living”) ) OR (TI=(homeless*) ) OR (AB=(“housing first” or “housing stability” or “housing instability” or “permanent housing” or “housing strateg*” or “housing polic*” or “housing project*” or “housing program*” or “housing quality” or “social housing*”) ) OR (TI=((autonomous or assisted or sheltered or support*) SAME accommodation)) OR (AB=(“autonomous housing” or “assisted housing” or “sheltered housing” or “autonomous accommodation” or “assisted accommodation” or “sheltered accommodation” or “autonomous dwelling*” or “assisted dwelling*” or “sheltered dwelling*”) ) OR (TS=(clubhouse or “club house”) ) OR (TI=(“autonomous living” or “independent living” or “assisted living” or “halfway house*” or “halfway home*” or “satellite hous*” or “satellite home*” or “community residences” or “group homes” or “community living” or “supervised apartments” or “therapeutic social clubs”) ) OR (AB=(“halfway house*” or “halfway home*” or “satellite hous*” or “satellite home*” or “supervised apartments” or “therapeutic social clubs”) ) OR (AB=((“autonomous living” or “independent living” or “assisted living” or “community residences” or “group homes” or “community living”) SAME (intervention* or program*) )) OR (TI=((independ* or assist* or support* or secur* or sustain* or maint*) SAME (tenanc* or tenure*) )) OR (AB=((independ* or assist* or support* or secur* or sustain* or maint*) SAME (tenanc* or tenure*) )) OR TI=("neighbourhood* characteristic*") OR (TS=("neighbourhood* intervention*" or "neighborhood* intervention*" or "neighborhood* program*" or "neighbourhood* program*") ) OR (TI=((environment* or housing or neighborhood* or neighbourhood*) SAME infrastructure)) OR (AB=((environment* or housing or neighborhood* or neighbourhood*) SAME infrastructure)) or (TI=(“built environment*” or “urban planning”) )) Timespan: 2020-07-01 to 2024-02-28 Date Run: Wed Feb 28 2024 11:07:17 GMT+0000 (Greenwich Mean Time) Results: 1596

12: (#11 AND #5) Timespan: 2020-07-01 to 2024-02-28 Date Run: Wed Feb 28 2024 11:07:34 GMT+0000 (Greenwich Mean Time) Results: 0

13: (#11 AND #7) Timespan: 2020-07-01 to 2024-02-28 Date Run: Wed Feb 28 2024 11:07:50 GMT+0000 (Greenwich Mean Time) Results: 0

14: ((TI=(money or cash or finan* or savings) ) OR (TI=("economic status" or pensions or remuneration or salary or salaries or "personal financ*" or "financial* autonom*" or "financial* secur*" or "paid work" or "financial* insecur*" or loans or borrowing or budgeting or "dealing with money" or microcredit or microfinance or "social fund*" or "economic hardship*" or "financial hardship*" or "family income*" or "family tax credit*" or "universal credit") ) OR (AB=("economic status" or pensions or remuneration or salary or salaries or "personal financ*" or "financial* autonom*" or "financial* secur*" or "paid work" or "financial* insecur*" or loans or borrowing or budgeting or "dealing with money" or microcredit or microfinance or "social fund*" or "economic hardship*" or "financial hardship*" or "family income*" or "family tax credit*" or "universal credit") ) OR (TI=(poverty or debt or debts or "social welfare") ) OR (AB=(poverty SAME prevent*) ) OR (AB=("high poverty" or "out of poverty" or "food poverty" or "fuel poverty" or "food bank*" or "welfare benefit*") ) OR (AB=("basic wage*" or "basic income*" or "minimum wage*" or "minimum income*" or "regular income*") ) OR (TI=("basic wage*" or "basic income*" or "minimum wage*" or "minimum income*" or "regular income*") )) Timespan: 2020-07-01 to 2024-02-28
Date Run: Wed Feb 28 2024 11:08:05 GMT+0000 (Greenwich Mean Time)
Results: 3577

15: (#5 and #14) Timespan: 2020-07-01 to 2024-02-28 Date Run: Wed Feb 28 2024 11:08:20 GMT+0000 (Greenwich Mean Time) Results: 1

**Search Name: CCRT_Barnett_Family**

Date Run: 28/02/2024 09:45:32

Comment:

| ID | Search | Hits |
|---|---|---|
| #1 | SR-DEPRESSN OR HS-DEPRESSN | 35371 |
| #2 | MeSH descriptor: [Mental Disorders] this term only | 5377 |
| #3 | MeSH descriptor: [Anxiety Disorders] explode all trees | 10069 |
| #4 | MeSH descriptor: [Trichotillomania] this term only | 96 |
| #5 | MeSH descriptor: [Feeding and Eating Disorders] this term only | 1185 |
| #6 | MeSH descriptor: [Anorexia Nervosa] this term only | 754 |
| #7 | MeSH descriptor: [Binge-Eating Disorder] this term only | 482 |
| #8 | MeSH descriptor: [Bulimia Nervosa] this term only | 362 |
| #9 | MeSH descriptor: [Mood Disorders] this term only | 1127 |
| #10 | MeSH descriptor: [Depressive Disorder] explode all trees | 16414 |
| #11 | MeSH descriptor: [Premenstrual Dysphoric Disorder] this term only | 51 |
| #12 | MeSH descriptor: [Reactive Attachment Disorder] this term only | 24 |
| #13 | MeSH descriptor: [Sexual Dysfunctions, Psychological] this term only | 504 |
| #14 | MeSH descriptor: [Somatoform Disorders] explode all trees | 945 |
| #15 | MeSH descriptor: [Munchausen Syndrome] explode all trees | 2 |
| #16 | MeSH descriptor: [Trauma and Stressor Related Disorders] explode all trees | 4508 |
| #17 | MeSH descriptor: [Trauma and Stressor Related Disorders] explode all trees | 4508 |
| #18 | MeSH descriptor: [Impulsive Behavior] explode all trees | 1672 |
| #19 | MeSH descriptor: [Hoarding] this term only | 7 |
| #20 | MeSH descriptor: [Self-Injurious Behavior] explode all trees | 2427 |

#21 (“anorexia nervosa” or “binge eating disorder” or bulimia or “eating disorder” or automutilation or “suicidal behaviour” or “self poisoning” or “suicidal ideation” or suicide or “suicide attempt” or depression or “agitated depression” or “atypical depression” or “chronic depression” or “depressive psychosis” or dysphoria or dysthymia or “endogenous depression” or involutional or “late life depression” or “major depression” or melancholia or “minor depression” or “mourning syndrome” or “organic depression” or “perinatal depression” or “antenatal depression” or “postnatal depression” or “post-stroke depression” or “postoperative depression” or “premenstrual dysphoric disorder” or “reactive depression” or “recurrent brief depression” or “seasonal affective disorder” or “treatment resistant depression” or neurosis or

“affective neurosis” or “anxiety neurosis” or dysthymia or hysteria or neurasthenia or psychasthenia or “adjustment disorder” or “anxiety disorder” or “acute stress disorder” or “generalized anxiety disorder” or panic or “posttraumatic stress disorder” or “separation anxiety” or “obsessive compulsive disorder” or compulsion or obsession or phobia or agoraphobia or claustrophobia or neophobia or “social phobia” or “somatoform disorder” or “body dysmorphic disorder” or “conversion disorder” or “delusional pregnancy” or hypochondriasis or “masked depression” or “psychogenic pain” or somatization or “mood disorder” or “affective neurosis” or “affective psychosis” or “blunted affect” or “major affective disorder” or “minor affective disorder” or “munchausen syndrome by proxy” or “munchausen syndrome” or “psychosexual disorder” or kleptomania or “trichotillomania” or “emotional disorder”):kw 63877

#22 (“MENTAL DISORDERS” or anhedonia or neurosis or “ACUTE STRESS DISORDER” or “adjustment disorders” or “attachment disorders” or “disinhibited social engagement disorder” or “posttraumatic stress disorder” or “complex ptsd” or desnos or “acute stress disorder” or “post-traumatic stress” or “traumatic neurosis” or “emotional trauma” or “AFFECTIVE DISORDERS” or “disruptive mood dysregulation disorder” or “dysthymic disorder” or “seasonal affective disorder” or “major depression” or “anaclitic depression” or “dysthymic disorder” or “endogenous depression” or “late life depression” or “postpartum depression” or “reactive depression” or “recurrent depression” or “treatment resistant depression” or “premenstrual dysphoric disorder” or “ANXIETY DISORDERS” or “generalized anxiety disorder” or “obsessive compulsive disorder” or “panic attack” or “panic disorder” or phobias or trichotillomania or acrophobia or agoraphobia or claustrophobia or ophidiophobia or “social phobia” or “separation anxiety disorder” or “EATING DISORDERS” or “anorexia nervosa” or “binge eating disorder” or bulimia or “SELF-INJURIOUS BEHAVIOR” or “self-destructive behavior” or “self-inflicted wounds” or “self-mutilation” or “self-poisoning” or suicide or “attempted suicide” or suicidality or “suicidal ideation” or “suicide prevention” or “SOMATOFORM DISORDERS” or “body dysmorphic disorder” or hypochondriasis or hysteria or neurasthenia or neurodermatitis or “somatization disorder” or “somatoform pain disorder” or “munchausen syndrome” or “munchausen syndrome by proxy” or (depression near/2 emotion) or "stress and trauma related disorders" or “mental health” or “mental health services” or “community mental health services” or “community counseling” or “community psychiatry”):kw45016

#23 ("acute stress" or (adjustment next disorder*) or ADNOS or (affective next disorder*) or agoraphobi* or "anorexia nervosa" or anxiety or astheni* or (attachment next disorder*) or (binge next eat*) or binging or (body next dysmorphi*) or bulimi* or (combat next disorder*) or (obsessive next compulsi*) or depression or depressed or depressive or dyssomni* or dyspareunia* or dysphori* or dysthymi* or dystoni* or (eating next disorder*) or EDNOS or (emotional next trauma) or fear or (health next anxiety) or hoarding or hyperactivity or hypochondri* or hysteri* or (medically next unexplained) or malingering or MDD or (common next mental) or (mental* near/2 (health or well*)) or mood or moods or munchausen or MUPS or mutism or neurastheni* or neurotic or neuros* or panic or phobi* or PND or ((post-trauma* or posttrauma*) next stress*) or psychogenic or psychosomatic or PTSD or (self next (injur* or harm or mutilat*)) or psychosexual or (psychological near/3 sexual near/3 dysfunction*) or "social anxiety" or somati* or somatoform or suicid* or parasuicid* or trichotillomani*):ti,ab 196731

#24 (depress* NEAR (employment or employability or unemployment or "return to work" or vocation* or absenteeism or presenteeism or "job security" or "work related" or "work focussed")):ab 241

#25 (#1 OR #2 OR #3 OR #4 OR #5 OR #6 OR #7 OR #8 OR #9 OR #10 OR #11 OR #12 OR #13 OR #14 OR #15 OR #16 OR #17 OR #18 OR #19 OR #20 OR #21 OR #22 OR #23 OR #24) 221261

#26 SR-SCIHZ OR HS-SCHIZ 941

#27 (bipolar or cyclothymi* or mania or manic or hypermani* or “rapid cycling” or (conversion next disorder*) or (dissociative next (amnesi* or fugue* or disorder*)) or "borderline state" or catatoni* or "character disorder" or delusion* or (capgras next syndrom*) or (diogenes next syndrom*) or depersonalization or depersonalisation or de-personalization or de-personalisation or (perceptual next disorder*) or (personality next disorder*) or BPD or paranoi* or psychiatr* or psychopathol* or psycho-pathol* or psychotic or psychosis* or psychoses* or psychotic* or schizo* or hebephreni* or “serious mental” or SMI):ti,ab,kw 69764

#28 MeSH descriptor: [Schizophrenia Spectrum and Other Psychotic Disorders] explode all trees 12178

#29 MeSH descriptor: [Personality Disorders] explode all trees 1828

#30 MeSH descriptor: [Bipolar and Related Disorders] explode all trees 3561

#31 (#26 OR #27 OR #28 OR #29 OR #30) 70436

#32 ("living arrangements" or "home environment" or "family conflict" or "family functioning" or "family relations" or "family reunification" or "intergenerational relations" or "marital conflict" or "marital relations" or divorce or divorced or "separated parent" or "single parent" or "living alone" or parenting or "parental involvement" or "parental role"):kw 4551

#33 ("child custody" or "joint custody" or "custodial care" or (child next visit*)):kw 100

#34 ((living next arrangement*) or (home next environment*) or (family and conflict*) or (family next function*) or (family next relations*) or "family reunification" or (intergenerational next relations*) or "marital conflict" or "marital relations" or divorce or divorced or "separated parent" or "single parent" or "living alone" or parenting or "parental involvement" or (parent* next role*)):ti 2235

#35 ((family or families or intergenerat* or inter-generat*) next (relation* or conflict or conflicts)):ti,ab 770

#36 ((sexual or intimate or partner or partners or marital) next (relation* or conflict or conflicts)):ti,ab 982

#37 ((develop* or enhanc* or initiative* or intervention* or program* or address* or improv* or promot* or target*) near/2 (relationship or relationships or relations)):ti,ab2011

#38 ((carer or carers or partner or partners or relations or relationship or relationships or marital) next support*):ti,ab 288

#39 (child* near/2 (access or contact or custody or maintenance)):ti,ab 487

#40 (((care next proceeding*) or (family next (court or courts)) or "child removal" or fostercare or “foster care”) and (parent* or mother* or father*)):ti,ab 141

#41 parents:ti or (parent* next outcome*):ti,ab 3767

#42 (parent* next (mental health)):ti,ab 537

#43 (#32 OR #33 OR #34 OR #35 OR #36 OR #37 OR #38 OR #39 OR #40 OR #41 OR #42) 12865

#44 #25 AND #43 with Cochrane Library publication date Between Jul 2020 and Feb 2024, in Trials 1649

#45 #31 AND #43 with Cochrane Library publication date Between Jul 2020 and Feb 2022, in Trials 114

**Search Name: CCRT_Barnett_Housing_CMD_SMI**

Date Run: 28/02/2024 09:32:03

Comment:

| ID | Search | Hits |
|---|---|---|
| #1 | SR-DEPRESSN OR HS-DEPRESSN | 35371 |
| #2 | MeSH descriptor: [Mental Disorders] this term only | 5377 |
| #3 | MeSH descriptor: [Anxiety Disorders] explode all trees | 10069 |
| #4 | MeSH descriptor: [Trichotillomania] this term only | 96 |
| #5 | MeSH descriptor: [Feeding and Eating Disorders] this term only | 1185 |
| #6 | MeSH descriptor: [Anorexia Nervosa] this term only | 754 |
| #7 | MeSH descriptor: [Binge-Eating Disorder] this term only | 482 |
| #8 | MeSH descriptor: [Bulimia Nervosa] this term only | 362 |
| #9 | MeSH descriptor: [Mood Disorders] this term only | 1127 |
| #10 | MeSH descriptor: [Depressive Disorder] explode all trees | 16414 |
| #11 | MeSH descriptor: [Premenstrual Dysphoric Disorder] this term only | 51 |
| #12 | MeSH descriptor: [Reactive Attachment Disorder] this term only | 24 |
| #13 | MeSH descriptor: [Sexual Dysfunctions, Psychological] this term only | 504 |
| #14 | MeSH descriptor: [Somatoform Disorders] explode all trees | 945 |
| #15 | MeSH descriptor: [Munchausen Syndrome] explode all trees | 2 |
| #16 | MeSH descriptor: [Trauma and Stressor Related Disorders] explode all trees | 4508 |
| #17 | MeSH descriptor: [Trauma and Stressor Related Disorders] explode all trees | 4508 |
| #18 | MeSH descriptor: [Impulsive Behavior] explode all trees | 1672 |
| #19 | MeSH descriptor: [Hoarding] this term only | 7 |
| #20 | MeSH descriptor: [Self-Injurious Behavior] explode all trees | 2427 |

#21 (“anorexia nervosa” or “binge eating disorder” or bulimia or “eating disorder” or automutilation or “suicidal behaviour” or “self poisoning” or “suicidal ideation” or suicide or “suicide attempt” or depression or “agitated depression” or “atypical depression” or “chronic depression” or “depressive psychosis” or dysphoria or dysthymia or “endogenous depression” or involutional or “late life depression” or “major depression” or melancholia or “minor depression” or “mourning syndrome” or “organic depression” or “perinatal depression” or “antenatal depression” or “postnatal depression” or “post-stroke depression” or “postoperative depression” or “premenstrual dysphoric disorder” or “reactive depression” or “recurrent brief depression” or “seasonal affective disorder” or “treatment resistant depression” or neurosis or

“affective neurosis” or “anxiety neurosis” or dysthymia or hysteria or neurasthenia or psychasthenia or “adjustment disorder” or “anxiety disorder” or “acute stress disorder” or “generalized anxiety disorder” or panic or “posttraumatic stress disorder” or “separation anxiety” or “obsessive compulsive disorder” or compulsion or obsession or phobia or agoraphobia or claustrophobia or neophobia or “social phobia” or “somatoform disorder” or “body dysmorphic disorder” or “conversion disorder” or “delusional pregnancy” or hypochondriasis or “masked depression” or “psychogenic pain” or somatization or “mood disorder” or “affective neurosis” or “affective psychosis” or “blunted affect” or “major affective disorder” or “minor affective disorder” or “munchausen syndrome by proxy” or “munchausen syndrome” or “psychosexual disorder” or kleptomania or “trichotillomania” or “emotional disorder”):kw 63877

#22 (“MENTAL DISORDERS” or anhedonia or neurosis or “ACUTE STRESS DISORDER” or “adjustment disorders” or “attachment disorders” or “disinhibited social engagement disorder” or “posttraumatic stress disorder” or “complex ptsd” or desnos or “acute stress disorder” or “post-traumatic stress” or “traumatic neurosis” or “emotional trauma” or “AFFECTIVE DISORDERS” or “disruptive mood dysregulation disorder” or “dysthymic disorder” or “seasonal affective disorder” or “major depression” or “anaclitic depression” or “dysthymic disorder” or “endogenous depression” or “late life depression” or “postpartum depression” or “reactive depression” or “recurrent depression” or “treatment resistant depression” or “premenstrual dysphoric disorder” or “ANXIETY DISORDERS” or “generalized anxiety disorder” or “obsessive compulsive disorder” or “panic attack” or “panic disorder” or phobias or trichotillomania or acrophobia or agoraphobia or claustrophobia or ophidiophobia or “social phobia” or “separation anxiety disorder” or “EATING DISORDERS” or “anorexia nervosa” or “binge eating disorder” or bulimia or “SELF-INJURIOUS BEHAVIOR” or “self-destructive behavior” or “self-inflicted wounds” or “self-mutilation” or “self-poisoning” or suicide or “attempted suicide” or suicidality or “suicidal ideation” or “suicide prevention” or “SOMATOFORM DISORDERS” or “body dysmorphic disorder” or hypochondriasis or hysteria or neurasthenia or neurodermatitis or “somatization disorder” or “somatoform pain disorder” or “munchausen syndrome” or “munchausen syndrome by proxy” or (depression near/2 emotion) or "stress and trauma related disorders" or “mental health” or “mental health services” or “community mental health services” or “community counseling” or “community psychiatry”):kw45016

#23 ("acute stress" or (adjustment next disorder*) or ADNOS or (affective next disorder*) or agoraphobi* or "anorexia nervosa" or anxiety or astheni* or (attachment next disorder*) or (binge next eat*) or binging or (body next dysmorphi*) or bulimi* or (combat next disorder*) or (obsessive next compulsi*) or depression or depressed or depressive or dyssomni* or dyspareunia* or dysphori* or dysthymi* or dystoni* or (eating next disorder*) or EDNOS or (emotional next trauma) or fear or (health next anxiety) or hoarding or hyperactivity or hypochondri* or hysteri* or (medically next unexplained) or malingering or MDD or (common next mental) or (mental* near/2 (health or well*)) or mood or moods or munchausen or MUPS or mutism or neurastheni* or neurotic or neuros* or panic or phobi* or PND or ((post-trauma* or posttrauma*) next stress*) or psychogenic or psychosomatic or PTSD or (self next (injur* or harm or mutilat*)) or psychosexual or (psychological near/3 sexual near/3 dysfunction*) or "social anxiety" or somati* or somatoform or suicid* or parasuicid* or trichotillomani*):ti,ab 196731

#24 (depress* NEAR (employment or employability or unemployment or "return to work" or vocation* or absenteeism or presenteeism or "job security" or "work related" or "work focussed")):ab 241

#25 (#1 OR #2 OR #3 OR #4 OR #5 OR #6 OR #7 OR #8 OR #9 OR #10 OR #11 OR #12 OR #13 OR #14 OR #15 OR #16 OR #17 OR #18 OR #19 OR #20 OR #21 OR #22 OR #23 OR #24) 221261

#26 SR-SCIHZ OR HS-SCHIZ 941

#27 (bipolar or cyclothymi* or mania or manic or hypermani* or “rapid cycling” or (conversion next disorder*) or (dissociative next (amnesi* or fugue* or disorder*)) or "borderline state" or catatoni* or "character disorder" or delusion* or (capgras next syndrom*) or (diogenes next syndrom*) or depersonalization or depersonalisation or de-personalization or de-personalisation or (perceptual next disorder*) or (personality next disorder*) or BPD or paranoi* or psychiatr* or psychopathol* or psycho-pathol* or psychotic or psychosis* or psychoses* or psychotic* or schizo* or hebephreni* or “serious mental” or SMI):ti,ab,kw 69764

#28 MeSH descriptor: [Schizophrenia Spectrum and Other Psychotic Disorders] explode all trees 12178

#29 MeSH descriptor: [Personality Disorders] explode all trees 1828

#30 MeSH descriptor: [Bipolar and Related Disorders] explode all trees 3561

#31 (#26 OR #27 OR #28 OR #29 OR #30) 70436

#32 (housing or “assisted living” or “independent living” or “community living” “group homes” or “public housing” or “living arrangements” or “emergency shelter” or homeless* or deinstitutionalization or “halfway house” or “halfway houses” or shelters or “residential care institutions” or “residence characteristics”):kw 3713

#33 (“built environment” or “environmental planning” or “city planning” or “urban planning” or neighborhood* or neighbourhood* or “social environment” or “psychosocial environment” or “social environments” or “poverty areas”):kw 2292

#34 ((chang* or develop* or enhanc* or initiative* or intervention* or program* or mitigat* or address* or improv* or target*) near/3 (housing or neighbo*)):ti,ab 580

#35 (dwelling* or housing):ti 2370

#36 homeless*:ti,ab,kw 1214

#37 (“housing first” or “permanent housing” or (housing near/2 (stability or instability))):ti,ab 255

#38 ((housing next (strateg* or polic* or project* or program* or quality)) or (new* next buil*) or “social housing”):ti,ab 126

#39 ((autonomous or assisted or sheltered or support* or public) near/3 (housing or accommodation or dwelling*)):ti,ab 467

#40 (((clubhouse or “club house”) next model*) or ((autonomous or independent or assisted) next living)):ti,ab,kw 1996

#41 (tenanc* or tenant* or tenure*):ti,ab,kw 206

#42 (((halfway or satellite) next (dwelling* or home* or house* or housing*)) or "community housing"):ti,ab,kw 63

#43 ((neighbo*) near/2 (characteristic* or intervention* or program*)):ti,ab 134

#44 ((neighbo* or housing) and infrastructure):ti,ab,kw 58

#45 neighbo*:ti or (neighbourhoods or neighborhoods):ti,ab,kw 968

#46 (#32 OR #33 OR #34 OR #35 OR #36 OR #37 OR #38 OR #39 OR #40 OR #41 OR #42 OR #43 OR #44 OR #45) 9447

#47 #25 AND #46 with Cochrane Library publication date Between Jul 2020 and Feb 2024, in Trials 726

#48 #31 AND #46 with Cochrane Library publication date Between Jul 2020 and Feb 2024, in Trials 155

**Search Name: CCRT_Barnett_Linking**

Date Run: 28/02/2024 11:12:33

Comment:

| ID | Search | Hits |
|---|---|---|
| #1 | SR-DEPRESSN OR HS-DEPRESSN | 35371 |
| #2 | MeSH descriptor: [Mental Disorders] this term only | 5377 |
| #3 | MeSH descriptor: [Anxiety Disorders] explode all trees | 10069 |
| #4 | MeSH descriptor: [Trichotillomania] this term only | 96 |
| #5 | MeSH descriptor: [Feeding and Eating Disorders] this term only | 1185 |
| #6 | MeSH descriptor: [Anorexia Nervosa] this term only | 754 |
| #7 | MeSH descriptor: [Binge-Eating Disorder] this term only | 482 |
| #8 | MeSH descriptor: [Bulimia Nervosa] this term only | 362 |
| #9 | MeSH descriptor: [Mood Disorders] this term only | 1127 |
| #10 | MeSH descriptor: [Depressive Disorder] explode all trees | 16414 |
| #11 | MeSH descriptor: [Premenstrual Dysphoric Disorder] this term only | 51 |
| #12 | MeSH descriptor: [Reactive Attachment Disorder] this term only | 24 |
| #13 | MeSH descriptor: [Sexual Dysfunctions, Psychological] this term only | 504 |
| #14 | MeSH descriptor: [Somatoform Disorders] explode all trees | 945 |
| #15 | MeSH descriptor: [Munchausen Syndrome] explode all trees | 2 |
| #16 | MeSH descriptor: [Trauma and Stressor Related Disorders] explode all trees | 4508 |
| #17 | MeSH descriptor: [Trauma and Stressor Related Disorders] explode all trees | 4508 |
| #18 | MeSH descriptor: [Impulsive Behavior] explode all trees | 1672 |
| #19 | MeSH descriptor: [Hoarding] this term only | 7 |
| #20 | MeSH descriptor: [Self-Injurious Behavior] explode all trees | 2427 |

#21 (“anorexia nervosa” or “binge eating disorder” or bulimia or “eating disorder” or automutilation or “suicidal behaviour” or “self poisoning” or “suicidal ideation” or suicide or “suicide attempt” or depression or “agitated depression” or “atypical depression” or “chronic depression” or “depressive psychosis” or dysphoria or dysthymia or “endogenous depression” or involutional or “late life depression” or “major depression” or melancholia or “minor depression” or “mourning syndrome” or “organic depression” or “perinatal depression” or “antenatal depression” or “postnatal depression” or “post-stroke depression” or “postoperative depression” or “premenstrual dysphoric disorder” or “reactive depression” or “recurrent brief depression” or “seasonal affective disorder” or “treatment resistant depression” or neurosis or

“affective neurosis” or “anxiety neurosis” or dysthymia or hysteria or neurasthenia or psychasthenia or “adjustment disorder” or “anxiety disorder” or “acute stress disorder” or “generalized anxiety disorder” or panic or “posttraumatic stress disorder” or “separation anxiety” or “obsessive compulsive disorder” or compulsion or obsession or phobia or agoraphobia or claustrophobia or neophobia or “social phobia” or “somatoform disorder” or “body dysmorphic disorder” or “conversion disorder” or “delusional pregnancy” or hypochondriasis or “masked depression” or “psychogenic pain” or somatization or “mood disorder” or “affective neurosis” or “affective psychosis” or “blunted affect” or “major affective disorder” or “minor affective disorder” or “munchausen syndrome by proxy” or “munchausen syndrome” or “psychosexual disorder” or kleptomania or “trichotillomania” or “emotional disorder”):kw 63877

#22 (“MENTAL DISORDERS” or anhedonia or neurosis or “ACUTE STRESS DISORDER” or “adjustment disorders” or “attachment disorders” or “disinhibited social engagement disorder” or “posttraumatic stress disorder” or “complex ptsd” or desnos or “acute stress disorder” or “post-traumatic stress” or “traumatic neurosis” or “emotional trauma” or “AFFECTIVE DISORDERS” or “disruptive mood dysregulation disorder” or “dysthymic disorder” or “seasonal affective disorder” or “major depression” or “anaclitic depression” or “dysthymic disorder” or “endogenous depression” or “late life depression” or “postpartum depression” or “reactive depression” or “recurrent depression” or “treatment resistant depression” or “premenstrual dysphoric disorder” or “ANXIETY DISORDERS” or “generalized anxiety disorder” or “obsessive compulsive disorder” or “panic attack” or “panic disorder” or phobias or trichotillomania or acrophobia or agoraphobia or claustrophobia or ophidiophobia or “social phobia” or “separation anxiety disorder” or “EATING DISORDERS” or “anorexia nervosa” or “binge eating disorder” or bulimia or “SELF-INJURIOUS BEHAVIOR” or “self-destructive behavior” or “self-inflicted wounds” or “self-mutilation” or “self-poisoning” or suicide or “attempted suicide” or suicidality or “suicidal ideation” or “suicide prevention” or “SOMATOFORM DISORDERS” or “body dysmorphic disorder” or hypochondriasis or hysteria or neurasthenia or neurodermatitis or “somatization disorder” or “somatoform pain disorder” or “munchausen syndrome” or “munchausen syndrome by proxy” or (depression near/2 emotion) or "stress and trauma related disorders" or “mental health” or “mental health services” or “community mental health services” or “community counseling” or “community psychiatry”):kw45016

#23 ("acute stress" or (adjustment next disorder*) or ADNOS or (affective next disorder*) or agoraphobi* or "anorexia nervosa" or anxiety or astheni* or (attachment next disorder*) or (binge next eat*) or binging or (body next dysmorphi*) or bulimi* or (combat next disorder*) or (obsessive next compulsi*) or depression or depressed or depressive or dyssomni* or dyspareunia* or dysphori* or dysthymi* or dystoni* or (eating next disorder*) or EDNOS or (emotional next trauma) or fear or (health next anxiety) or hoarding or hyperactivity or hypochondri* or hysteri* or (medically next unexplained) or malingering or MDD or (common next mental) or (mental* near/2 (health or well*)) or mood or moods or munchausen or MUPS or mutism or neurastheni* or neurotic or neuros* or panic or phobi* or PND or ((post-trauma* or posttrauma*) next stress*) or psychogenic or psychosomatic or PTSD or (self next (injur* or harm or mutilat*)) or psychosexual or (psychological near/3 sexual near/3 dysfunction*) or "social anxiety" or somati* or somatoform or suicid* or parasuicid* or trichotillomani*):ti,ab 196731

#24 (depress* NEAR (employment or employability or unemployment or "return to work" or vocation* or absenteeism or presenteeism or "job security" or "work related" or "work focussed")):ab 241

#25 (#1 OR #2 OR #3 OR #4 OR #5 OR #6 OR #7 OR #8 OR #9 OR #10 OR #11 OR #12 OR #13 OR #14 OR #15 OR #16 OR #17 OR #18 OR #19 OR #20 OR #21 OR #22 OR #23 OR #24) 221261

#26 SR-SCIHZ OR HS-SCHIZ 941

#27 (bipolar or cyclothymi* or mania or manic or hypermani* or “rapid cycling” or (conversion next disorder*) or (dissociative next (amnesi* or fugue* or disorder*)) or "borderline state" or catatoni* or "character disorder" or delusion* or (capgras next syndrom*) or (diogenes next syndrom*) or depersonalization or depersonalisation or de-personalization or de-personalisation or (perceptual next disorder*) or (personality next disorder*) or BPD or paranoi* or psychiatr* or psychopathol* or psycho-pathol* or psychotic or psychosis* or psychoses* or psychotic* or schizo* or hebephreni* or “serious mental” or SMI):ti,ab,kw 69764

#28 MeSH descriptor: [Schizophrenia Spectrum and Other Psychotic Disorders] explode all trees 12178

#29 MeSH descriptor: [Personality Disorders] explode all trees 1828

#30 MeSH descriptor: [Bipolar and Related Disorders] explode all trees 3561

#31 (#26 OR #27 OR #28 OR #29 OR #30) 70436

#32 ((chang* or develop* or enhanc* or initiative* or intervention* or program* or mitigat* or address* or improv* or target*) NEAR (community or living or social) NEAR (condition or conditions or circumstance*)):ti,ab,kw 815

#33 (social* NEAR/3 prescri*):ti,ab,kw 77

#34 (communit* next (connect* or engagement* or link* or referral* or intervention* or wellbeing)):ti,ab,kw 2341

#35 "sense of belonging":ti,ab,kw 83

#36 (#32 OR #33 OR #34 OR #35) 3251

#37 (#25 and #36) with Cochrane Library publication date Between Jul 2020 and Feb 2024, in Trials 268

#38 (#31 and #36) with Cochrane Library publication date Between Jul 2020 and Feb 2024, in Trials 53

**Search Name: CCRT_Barnett_Money**

Date Run: 28/02/2024 09:23:07

Comment:

| ID | Search | Hits |
|---|---|---|
| #1 | SR-DEPRESSN OR HS-DEPRESSN | 35371 |
| #2 | MeSH descriptor: [Mental Disorders] this term only | 5377 |
| #3 | MeSH descriptor: [Anxiety Disorders] explode all trees | 10069 |
| #4 | MeSH descriptor: [Trichotillomania] this term only | 96 |
| #5 | MeSH descriptor: [Feeding and Eating Disorders] this term only | 1185 |
| #6 | MeSH descriptor: [Anorexia Nervosa] this term only | 754 |
| #7 | MeSH descriptor: [Binge-Eating Disorder] this term only | 482 |
| #8 | MeSH descriptor: [Bulimia Nervosa] this term only | 362 |
| #9 | MeSH descriptor: [Mood Disorders] this term only | 1127 |
| #10 | MeSH descriptor: [Depressive Disorder] explode all trees | 16414 |
| #11 | MeSH descriptor: [Premenstrual Dysphoric Disorder] this term only | 51 |
| #12 | MeSH descriptor: [Reactive Attachment Disorder] this term only | 24 |
| #13 | MeSH descriptor: [Sexual Dysfunctions, Psychological] this term only | 504 |
| #14 | MeSH descriptor: [Somatoform Disorders] explode all trees | 945 |
| #15 | MeSH descriptor: [Munchausen Syndrome] explode all trees | 2 |
| #16 | MeSH descriptor: [Trauma and Stressor Related Disorders] explode all trees | 4508 |
| #17 | MeSH descriptor: [Trauma and Stressor Related Disorders] explode all trees | 4508 |
| #18 | MeSH descriptor: [Impulsive Behavior] explode all trees | 1672 |
| #19 | MeSH descriptor: [Hoarding] this term only | 7 |
| #20 | MeSH descriptor: [Self-Injurious Behavior] explode all trees | 2427 |

#21 (“anorexia nervosa” or “binge eating disorder” or bulimia or “eating disorder” or automutilation or “suicidal behaviour” or “self poisoning” or “suicidal ideation” or suicide or “suicide attempt” or depression or “agitated depression” or “atypical depression” or “chronic depression” or “depressive psychosis” or dysphoria or dysthymia or “endogenous depression” or involutional or “late life depression” or “major depression” or melancholia or “minor depression” or “mourning syndrome” or “organic depression” or “perinatal depression” or “antenatal depression” or “postnatal depression” or “post-stroke depression” or “postoperative depression” or “premenstrual dysphoric disorder” or “reactive depression” or “recurrent brief depression” or “seasonal affective disorder” or “treatment resistant depression” or neurosis or

“affective neurosis” or “anxiety neurosis” or dysthymia or hysteria or neurasthenia or psychasthenia or “adjustment disorder” or “anxiety disorder” or “acute stress disorder” or “generalized anxiety disorder” or panic or “posttraumatic stress disorder” or “separation anxiety” or “obsessive compulsive disorder” or compulsion or obsession or phobia or agoraphobia or claustrophobia or neophobia or “social phobia” or “somatoform disorder” or “body dysmorphic disorder” or “conversion disorder” or “delusional pregnancy” or hypochondriasis or “masked depression” or “psychogenic pain” or somatization or “mood disorder” or “affective neurosis” or “affective psychosis” or “blunted affect” or “major affective disorder” or “minor affective disorder” or “munchausen syndrome by proxy” or “munchausen syndrome” or “psychosexual disorder” or kleptomania or “trichotillomania” or “emotional disorder”):kw 63877

#22 (“MENTAL DISORDERS” or anhedonia or neurosis or “ACUTE STRESS DISORDER” or “adjustment disorders” or “attachment disorders” or “disinhibited social engagement disorder” or “posttraumatic stress disorder” or “complex ptsd” or desnos or “acute stress disorder” or “post-traumatic stress” or “traumatic neurosis” or “emotional trauma” or “AFFECTIVE DISORDERS” or “disruptive mood dysregulation disorder” or “dysthymic disorder” or “seasonal affective disorder” or “major depression” or “anaclitic depression” or “dysthymic disorder” or “endogenous depression” or “late life depression” or “postpartum depression” or “reactive depression” or “recurrent depression” or “treatment resistant depression” or “premenstrual dysphoric disorder” or “ANXIETY DISORDERS” or “generalized anxiety disorder” or “obsessive compulsive disorder” or “panic attack” or “panic disorder” or phobias or trichotillomania or acrophobia or agoraphobia or claustrophobia or ophidiophobia or “social phobia” or “separation anxiety disorder” or “EATING DISORDERS” or “anorexia nervosa” or “binge eating disorder” or bulimia or “SELF-INJURIOUS BEHAVIOR” or “self-destructive behavior” or “self-inflicted wounds” or “self-mutilation” or “self-poisoning” or suicide or “attempted suicide” or suicidality or “suicidal ideation” or “suicide prevention” or “SOMATOFORM DISORDERS” or “body dysmorphic disorder” or hypochondriasis or hysteria or neurasthenia or neurodermatitis or “somatization disorder” or “somatoform pain disorder” or “munchausen syndrome” or “munchausen syndrome by proxy” or (depression near/2 emotion) or "stress and trauma related disorders" or “mental health” or “mental health services” or “community mental health services” or “community counseling” or “community psychiatry”):kw45016

#23 ("acute stress" or (adjustment next disorder*) or ADNOS or (affective next disorder*) or agoraphobi* or "anorexia nervosa" or anxiety or astheni* or (attachment next disorder*) or (binge next eat*) or binging or (body next dysmorphi*) or bulimi* or (combat next disorder*) or (obsessive next compulsi*) or depression or depressed or depressive or dyssomni* or dyspareunia* or dysphori* or dysthymi* or dystoni* or (eating next disorder*) or EDNOS or (emotional next trauma) or fear or (health next anxiety) or hoarding or hyperactivity or hypochondri* or hysteri* or (medically next unexplained) or malingering or MDD or (common next mental) or (mental* near/2 (health or well*)) or mood or moods or munchausen or MUPS or mutism or neurastheni* or neurotic or neuros* or panic or phobi* or PND or ((post-trauma* or posttrauma*) next stress*) or psychogenic or psychosomatic or PTSD or (self next (injur* or harm or mutilat*)) or psychosexual or (psychological near/3 sexual near/3 dysfunction*) or "social anxiety" or somati* or somatoform or suicid* or parasuicid* or trichotillomani*):ti,ab 196731

#24 (depress* NEAR (employment or employability or unemployment or "return to work" or vocation* or absenteeism or presenteeism or "job security" or "work related" or "work focussed")):ab 241

#25 (#1 OR #2 OR #3 OR #4 OR #5 OR #6 OR #7 OR #8 OR #9 OR #10 OR #11 OR #12 OR #13 OR #14 OR #15 OR #16 OR #17 OR #18 OR #19 OR #20 OR #21 OR #22 OR #23 OR #24) 221261

#26 SR-SCIHZ OR HS-SCHIZ 941

#27 (bipolar or cyclothymi* or mania or manic or hypermani* or “rapid cycling” or (conversion next disorder*) or (dissociative next (amnesi* or fugue* or disorder*)) or "borderline state" or catatoni* or "character disorder" or delusion* or (capgras next syndrom*) or (diogenes next syndrom*) or depersonalization or depersonalisation or de-personalization or de-personalisation or (perceptual next disorder*) or (personality next disorder*) or BPD or paranoi* or psychiatr* or psychopathol* or psycho-pathol* or psychotic or psychosis* or psychoses* or psychotic* or schizo* or hebephreni* or “serious mental” or SMI):ti,ab,kw 69764

#28 MeSH descriptor: [Schizophrenia Spectrum and Other Psychotic Disorders] explode all trees 12178

#29 MeSH descriptor: [Personality Disorders] explode all trees 1828

#30 MeSH descriptor: [Bipolar and Related Disorders] explode all trees 3561

#31 (#26 OR #27 OR #28 OR #29 OR #30) 70436

#32 (money or remuneration or salary or salaries or socioeconomics):kw 1843

#33 ((personal near/2 financ*) or (income near/2 (family or household or personal or level)) or "financial management" or "fringe benefit" or bonuses or pension or pensions):kw 1839

#34 (disadvantaged or “social deprivation” or “human needs” or “basic needs” or “personal needs” or “social needs” or welfare):kw 2079

#35 (“economic status” or “socioeconomic status” or “economic security” or “financial strain” or (employee and (assistance or benefits)) or “health insurance” or “workers compensation” or Medicaid):kw 2662

#36 ((access* or improv* or manag* or supplement*) NEAR/2 (cash or money or finance or finances or financial or income or incomes or savings)):ti,ab 552

#37 (financ* near/2 (autonomy or autonomous or security or insecurity or insecurities)):ti,ab 71

#38 (loans or borrowing or budgeting or microcredit or microfinance or (social* next fund*) or “high poverty” or ((income* or wage*) near/2 supplement*)):ti,ab,kw 469

#39 ((address* or escap* or improv* or support* or target*) near/2 (deprived or deprivation or deprivations or poor or poverty)):ti,ab,kw 630

#40 (“food poverty” or “fuel poverty” or “food bank” or “food banks”):ti,ab,kw 38

#41 ((alleviat* or ease or easing or manag* or prevent* or reduc* or stop*) near/2 (poverty or “economic hardship” or “economic hardships” or “financial hardship” or “financial hardships”)):ti,ab 142

#42 ((alleviat* or eas* or easing or manag* or prevent* or reduc* or relief or stop*) near/2 (debt or debts)):ti,ab 11

#43 ((basic or minimum) NEAR/3 (wage* or income*)):ti,ab,kw 55

#44 (“zero hours” or “paid work” or “paid employment” or "paid leave" or “family income” or “family incomes” or “tax credit” or “tax credits” or “welfare benefit” or “welfare benefits”):ti,ab,kw 783

#45 ("out of poverty" or "dealing with money"):ab 9

#46 (money or poverty or debt* or welfare):ti 577

#47 (#32 OR #33 OR #34 OR #35 OR #36 OR #37 OR #38 OR #39 OR #40 OR #41 OR #42 OR #43 OR #44 OR #45 OR #46) 10518

#48 #25 AND #47 with Cochrane Library publication date Between Jul 2020 and Feb 2024 707

#49 #31 AND #47 with Cochrane Library publication date Between Jul 2020 and Feb 2024 141

**Search Name: CCRT_Barnett_Rights**

Date Run: 28/02/2024 09:58:13

Comment:

| ID | Search | Hits |
|---|---|---|
| #1 | SR-DEPRESSN OR HS-DEPRESSN | 35371 |
| #2 | MeSH descriptor: [Mental Disorders] this term only | 5377 |
| #3 | MeSH descriptor: [Anxiety Disorders] explode all trees | 10069 |
| #4 | MeSH descriptor: [Trichotillomania] this term only | 96 |
| #5 | MeSH descriptor: [Feeding and Eating Disorders] this term only | 1185 |
| #6 | MeSH descriptor: [Anorexia Nervosa] this term only | 754 |
| #7 | MeSH descriptor: [Binge-Eating Disorder] this term only | 482 |
| #8 | MeSH descriptor: [Bulimia Nervosa] this term only | 362 |
| #9 | MeSH descriptor: [Mood Disorders] this term only | 1127 |
| #10 | MeSH descriptor: [Depressive Disorder] explode all trees | 16414 |
| #11 | MeSH descriptor: [Premenstrual Dysphoric Disorder] this term only | 51 |
| #12 | MeSH descriptor: [Reactive Attachment Disorder] this term only | 24 |
| #13 | MeSH descriptor: [Sexual Dysfunctions, Psychological] this term only | 504 |
| #14 | MeSH descriptor: [Somatoform Disorders] explode all trees | 945 |
| #15 | MeSH descriptor: [Munchausen Syndrome] explode all trees | 2 |
| #16 | MeSH descriptor: [Trauma and Stressor Related Disorders] explode all trees | 4508 |
| #17 | MeSH descriptor: [Trauma and Stressor Related Disorders] explode all trees | 4508 |
| #18 | MeSH descriptor: [Impulsive Behavior] explode all trees | 1672 |
| #19 | MeSH descriptor: [Hoarding] this term only | 7 |
| #20 | MeSH descriptor: [Self-Injurious Behavior] explode all trees | 2427 |

#21 (“anorexia nervosa” or “binge eating disorder” or bulimia or “eating disorder” or automutilation or “suicidal behaviour” or “self poisoning” or “suicidal ideation” or suicide or “suicide attempt” or depression or “agitated depression” or “atypical depression” or “chronic depression” or “depressive psychosis” or dysphoria or dysthymia or “endogenous depression” or involutional or “late life depression” or “major depression” or melancholia or “minor depression” or “mourning syndrome” or “organic depression” or “perinatal depression” or “antenatal depression” or “postnatal depression” or “post-stroke depression” or “postoperative depression” or “premenstrual dysphoric disorder” or “reactive depression” or “recurrent brief depression” or “seasonal affective disorder” or “treatment resistant depression” or neurosis or

“affective neurosis” or “anxiety neurosis” or dysthymia or hysteria or neurasthenia or psychasthenia or “adjustment disorder” or “anxiety disorder” or “acute stress disorder” or “generalized anxiety disorder” or panic or “posttraumatic stress disorder” or “separation anxiety” or “obsessive compulsive disorder” or compulsion or obsession or phobia or agoraphobia or claustrophobia or neophobia or “social phobia” or “somatoform disorder” or “body dysmorphic disorder” or “conversion disorder” or “delusional pregnancy” or hypochondriasis or “masked depression” or “psychogenic pain” or somatization or “mood disorder” or “affective neurosis” or “affective psychosis” or “blunted affect” or “major affective disorder” or “minor affective disorder” or “munchausen syndrome by proxy” or “munchausen syndrome” or “psychosexual disorder” or kleptomania or “trichotillomania” or “emotional disorder”):kw 63877

#22 (“MENTAL DISORDERS” or anhedonia or neurosis or “ACUTE STRESS DISORDER” or “adjustment disorders” or “attachment disorders” or “disinhibited social engagement disorder” or “posttraumatic stress disorder” or “complex ptsd” or desnos or “acute stress disorder” or “post-traumatic stress” or “traumatic neurosis” or “emotional trauma” or “AFFECTIVE DISORDERS” or “disruptive mood dysregulation disorder” or “dysthymic disorder” or “seasonal affective disorder” or “major depression” or “anaclitic depression” or “dysthymic disorder” or “endogenous depression” or “late life depression” or “postpartum depression” or “reactive depression” or “recurrent depression” or “treatment resistant depression” or “premenstrual dysphoric disorder” or “ANXIETY DISORDERS” or “generalized anxiety disorder” or “obsessive compulsive disorder” or “panic attack” or “panic disorder” or phobias or trichotillomania or acrophobia or agoraphobia or claustrophobia or ophidiophobia or “social phobia” or “separation anxiety disorder” or “EATING DISORDERS” or “anorexia nervosa” or “binge eating disorder” or bulimia or “SELF-INJURIOUS BEHAVIOR” or “self-destructive behavior” or “self-inflicted wounds” or “self-mutilation” or “self-poisoning” or suicide or “attempted suicide” or suicidality or “suicidal ideation” or “suicide prevention” or “SOMATOFORM DISORDERS” or “body dysmorphic disorder” or hypochondriasis or hysteria or neurasthenia or neurodermatitis or “somatization disorder” or “somatoform pain disorder” or “munchausen syndrome” or “munchausen syndrome by proxy” or (depression near/2 emotion) or "stress and trauma related disorders" or “mental health” or “mental health services” or “community mental health services” or “community counseling” or “community psychiatry”):kw45016

#23 ("acute stress" or (adjustment next disorder*) or ADNOS or (affective next disorder*) or agoraphobi* or "anorexia nervosa" or anxiety or astheni* or (attachment next disorder*) or (binge next eat*) or binging or (body next dysmorphi*) or bulimi* or (combat next disorder*) or (obsessive next compulsi*) or depression or depressed or depressive or dyssomni* or dyspareunia* or dysphori* or dysthymi* or dystoni* or (eating next disorder*) or EDNOS or (emotional next trauma) or fear or (health next anxiety) or hoarding or hyperactivity or hypochondri* or hysteri* or (medically next unexplained) or malingering or MDD or (common next mental) or (mental* near/2 (health or well*)) or mood or moods or munchausen or MUPS or mutism or neurastheni* or neurotic or neuros* or panic or phobi* or PND or ((post-trauma* or posttrauma*) next stress*) or psychogenic or psychosomatic or PTSD or (self next (injur* or harm or mutilat*)) or psychosexual or (psychological near/3 sexual near/3 dysfunction*) or "social anxiety" or somati* or somatoform or suicid* or parasuicid* or trichotillomani*):ti,ab 196731

#24 (depress* NEAR (employment or employability or unemployment or "return to work" or vocation* or absenteeism or presenteeism or "job security" or "work related" or "work focussed")):ab 241

#25 (#1 OR #2 OR #3 OR #4 OR #5 OR #6 OR #7 OR #8 OR #9 OR #10 OR #11 OR #12 OR #13 OR #14 OR #15 OR #16 OR #17 OR #18 OR #19 OR #20 OR #21 OR #22 OR #23 OR #24) 221261

#26 SR-SCIHZ OR HS-SCHIZ 941

#27 (bipolar or cyclothymi* or mania or manic or hypermani* or "rapid cycling" or (conversion next disorder*) or (dissociative next (amnesi* or fugue* or disorder*)) or "borderline state" or catatoni* or "character disorder" or delusion* or (capgras next syndrom*) or (diogenes next syndrom*) or depersonalization or depersonalisation or de-personalization or de-personalisation or (perceptual next disorder*) or (personality next disorder*) or BPD or paranoi* or psychiatr* or psychopathol* or psycho-pathol* or psychotic or psychosis* or psychoses* or psychotic* or schizo* or hebephreni* or "serious mental" or SMI):ti,ab,kw 69764

#28 MeSH descriptor: [Schizophrenia Spectrum and Other Psychotic Disorders] explode all trees 12178

#29 MeSH descriptor: [Personality Disorders] explode all trees 1828

#30 MeSH descriptor: [Bipolar and Related Disorders] explode all trees 3561

#31 (#26 OR #27 OR #28 OR #29 OR #30) 70436

#32 rights:kw,ti 165

#33 (freedoms or citizenship):ti,ab,kw 261

#34 ("human rights" or "civil rights" or "reproductive rights" or ((women* or woman*) next rights)):ti,ab,kw 215

#35 ((citizen* or civil* or human* or legal or social or vote or voting) near/2 (right or rights)):ti,ab 222

#36 ("social justice" or "social protection" or "equal protection" or "personal autonomy"):ti,ab,kw 600

#37 ("public policy" or "public policies" or "government policy" or "policy making"):kw,ti 296

#38 ((government* or public or social) next polic*):ti,kw 206

#39 ((equity or equalit* or inequit* or inequalit* or dispar*) near focus*):ti,ab 107

#40 ("Social Determinants of Health" or "health disparities"):ti,kw 464

#41 ((social or societ* or communit* or neighbo*) near/3 (equit* or inequit* or inequalit* or dispar*)):ti,ab,kw 318

#42 ("digital divide" or "information literacy"):ti,ab,kw 90

#43 ((accessib* or "access to") near (computer* or internet)):ti,kw 32

#44 (digital* near/2 (inclusi* or exclusi* or divide* or disparit* or equit* or inequit* or inequalit*)):ti,ab,kw 309

#45 (#32 OR #33 OR #34 OR #35 OR #36 OR #37 OR #38 OR #39 OR #40 OR #41 OR #42 OR #43 OR #44) 2718

#46 (#25 and #45) 534

#47 CINAHL:AN 29651

#48 (#46 NOT #47) with Cochrane Library publication date Between Jul 2020 and Feb 2024, in Trials211

#49 (#31 and #45) 144

#50 (#49 NOT #47) with Cochrane Library publication date Between Jul 2020 and Feb 2024, in Trials36

**Search Name: CCRT_Barnett_Social Isolation**

Date Run: 28/02/2024 10:47:37

Comment:

ID Search Hits

#1 SR-DEPRESSN OR HS-DEPRESSN 35371

#2 MeSH descriptor: [Mental Disorders] this term only 5377

#3 MeSH descriptor: [Anxiety Disorders] explode all trees 10069

#4 MeSH descriptor: [Trichotillomania] this term only 96

#5 MeSH descriptor: [Feeding and Eating Disorders] this term only 1185

#6 MeSH descriptor: [Anorexia Nervosa] this term only 754

#7 MeSH descriptor: [Binge-Eating Disorder] this term only 482

#8 MeSH descriptor: [Bulimia Nervosa] this term only 362

#9 MeSH descriptor: [Mood Disorders] this term only 1127

#10 MeSH descriptor: [Depressive Disorder] explode all trees 16414

#11 MeSH descriptor: [Premenstrual Dysphoric Disorder] this term only 51

#12 MeSH descriptor: [Reactive Attachment Disorder] this term only 24

#13 MeSH descriptor: [Sexual Dysfunctions, Psychological] this term only 504

#14 MeSH descriptor: [Somatoform Disorders] explode all trees 945

#15 MeSH descriptor: [Munchausen Syndrome] explode all trees 2

#16 MeSH descriptor: [Trauma and Stressor Related Disorders] explode all trees 4508

#17 MeSH descriptor: [Trauma and Stressor Related Disorders] explode all trees 4508

#18 MeSH descriptor: [Impulsive Behavior] explode all trees 1672

#19 MeSH descriptor: [Hoarding] this term only 7

#20 MeSH descriptor: [Self-Injurious Behavior] explode all trees 2427

#21 (“anorexia nervosa” or “binge eating disorder” or bulimia or “eating disorder” or automutilation or “suicidal behaviour” or “self poisoning” or “suicidal ideation” or suicide or “suicide attempt” or depression or “agitated depression” or “atypical depression” or “chronic depression” or “depressive psychosis” or dysphoria or dysthymia or “endogenous depression” or involutional or “late life depression” or “major depression” or melancholia or “minor depression” or “mourning syndrome” or “organic depression” or “perinatal depression” or “antenatal depression” or “postnatal depression” or “post-stroke depression” or “postoperative depression” or “premenstrual dysphoric disorder” or “reactive depression” or “recurrent brief depression” or “seasonal affective disorder” or “treatment resistant depression” or neurosis or

“affective neurosis” or “anxiety neurosis” or dysthymia or hysteria or neurasthenia or psychasthenia or “adjustment disorder” or “anxiety disorder” or “acute stress disorder” or “generalized anxiety disorder” or panic or “posttraumatic stress disorder” or “separation anxiety” or “obsessive compulsive disorder” or compulsion or obsession or phobia or agoraphobia or claustrophobia or neophobia or “social phobia” or “somatoform disorder” or “body dysmorphic disorder” or “conversion disorder” or “delusional pregnancy” or hypochondriasis or “masked depression” or “psychogenic pain” or somatization or “mood disorder” or “affective neurosis” or “affective psychosis” or “blunted affect” or “major affective disorder” or “minor affective disorder” or “munchausen syndrome by proxy” or “munchausen syndrome” or “psychosexual disorder” or kleptomania or “trichotillomania” or “emotional disorder”):kw 63877

#22 (“MENTAL DISORDERS” or anhedonia or neurosis or “ACUTE STRESS DISORDER” or “adjustment disorders” or “attachment disorders” or “disinhibited social engagement disorder” or “posttraumatic stress disorder” or “complex ptsd” or desnos or “acute stress disorder” or “post-traumatic stress” or “traumatic neurosis” or “emotional trauma” or “AFFECTIVE DISORDERS” or “disruptive mood dysregulation disorder” or “dysthymic disorder” or “seasonal affective disorder” or “major depression” or “anaclitic depression” or “dysthymic disorder” or “endogenous depression” or “late life depression” or “postpartum depression” or “reactive depression” or “recurrent depression” or “treatment resistant depression” or “premenstrual dysphoric disorder” or “ANXIETY DISORDERS” or “generalized anxiety disorder” or “obsessive compulsive disorder” or “panic attack” or “panic disorder” or phobias or trichotillomania or acrophobia or agoraphobia or claustrophobia or ophidiophobia or “social phobia” or “separation anxiety disorder” or “EATING DISORDERS” or “anorexia nervosa” or “binge eating disorder” or bulimia or “SELF-INJURIOUS BEHAVIOR” or “self-destructive behavior” or “self-inflicted wounds” or “self-mutilation” or “self-poisoning” or suicide or “attempted suicide” or suicidality or “suicidal ideation” or “suicide prevention” or “SOMATOFORM DISORDERS” or “body dysmorphic disorder” or hypochondriasis or hysteria or neurasthenia or neurodermatitis or “somatization disorder” or “somatoform pain disorder” or “munchausen syndrome” or “munchausen syndrome by proxy” or (depression near/2 emotion) or "stress and trauma related disorders" or “mental health” or “mental health services” or “community mental health services” or “community counseling” or “community psychiatry”):kw45016

#23 ("acute stress" or (adjustment next disorder*) or ADNOS or (affective next disorder*) or agoraphobi* or "anorexia nervosa" or anxiety or astheni* or (attachment next disorder*) or (binge next eat*) or binging or (body next dysmorphi*) or bulimi* or (combat next disorder*) or (obsessive next compulsi*) or depression or depressed or depressive or dyssomni* or dyspareunia* or dysphori* or dysthymi* or dystoni* or (eating next disorder*) or EDNOS or (emotional next trauma) or fear or (health next anxiety) or hoarding or hyperactivity or hypochondri* or hysteri* or (medically next unexplained) or malingering or MDD or (common next mental) or (mental* near/2 (health or well*)) or mood or moods or munchausen or MUPS or mutism or neurastheni* or neurotic or neuros* or panic or phobi* or PND or ((post-trauma* or posttrauma*) next stress*) or psychogenic or psychosomatic or PTSD or (self next (injur* or harm or mutilat*)) or psychosexual or (psychological near/3 sexual near/3 dysfunction*) or "social anxiety" or somati* or somatoform or suicid* or parasuicid* or trichotillomani*):ti,ab 196731

#24 (depress* NEAR (employment or employability or unemployment or "return to work" or vocation* or absenteeism or presenteeism or "job security" or "work related" or "work focussed")):ab 241

#25 (#1 OR #2 OR #3 OR #4 OR #5 OR #6 OR #7 OR #8 OR #9 OR #10 OR #11 OR #12 OR #13 OR #14 OR #15 OR #16 OR #17 OR #18 OR #19 OR #20 OR #21 OR #22 OR #23 OR #24) 221261

#26 SR-SCIHZ OR HS-SCHIZ 941

#27 (bipolar or cyclothymi* or mania or manic or hypermani* or “rapid cycling” or (conversion next disorder*) or (dissociative next (amnesi* or fugue* or disorder*)) or "borderline state" or catatoni* or "character disorder" or delusion* or (capgras next syndrom*) or (diogenes next syndrom*) or depersonalization or depersonalisation or de-personalization or de-personalisation or (perceptual next disorder*) or (personality next disorder*) or BPD or paranoi* or psychiatr* or psychopathol* or psycho-pathol* or psychotic or psychosis* or psychoses* or psychotic* or schizo* or hebephreni* or “serious mental” or SMI):ti,ab,kw 69764

#28 MeSH descriptor: [Schizophrenia Spectrum and Other Psychotic Disorders] explode all trees 12178

#29 MeSH descriptor: [Personality Disorders] explode all trees 1828

#30 MeSH descriptor: [Bipolar and Related Disorders] explode all trees 3561

#31 (#26 OR #27 OR #28 OR #29 OR #30) 70436

#32 (loneliness or lonely):ti,ab,kw 1395

#33 ((social* or societ* or communit*) near (isolated or isolation)):ti,ab,kw 1724

#34 ((alleviat* or ease or manag* or mitigat* or prevent* or overcom* or reduc* or stop*) near/2 (isolation or isolated)):ti,ab,kw 324

#35 ((address* or enhanc* or improv* or increas* or promot* or target*) near/2 (inclusion or inclusivity)):ti,ab,kw 673

#36 (#32 or #33 or #34 or #35) 3739

#37 (#25 and #36) with Cochrane Library publication date Between Jul 2020 and Feb 2024, in Trials 916

#38 (#31 and #36) with Cochrane Library publication date Between Jul 2020 and Feb 2024, in Trials 173

**Search Name: CCRT_Barnett_Social Participation CMD**

Date Run: 28/02/2024 11:04:53

Comment:

ID Search Hits

#1 SR-DEPRESSN OR HS-DEPRESSN 35371

#2 MeSH descriptor: [Mental Disorders] this term only 5377

#3 MeSH descriptor: [Anxiety Disorders] explode all trees 10069

#4 MeSH descriptor: [Trichotillomania] this term only 96

#5 MeSH descriptor: [Feeding and Eating Disorders] this term only 1185

#6 MeSH descriptor: [Anorexia Nervosa] this term only 754

#7 MeSH descriptor: [Binge-Eating Disorder] this term only 482

#8 MeSH descriptor: [Bulimia Nervosa] this term only 362

#9 MeSH descriptor: [Mood Disorders] this term only 1127

#10 MeSH descriptor: [Depressive Disorder] explode all trees 16414

#11 MeSH descriptor: [Premenstrual Dysphoric Disorder] this term only 51

#12 MeSH descriptor: [Reactive Attachment Disorder] this term only 24

#13 MeSH descriptor: [Sexual Dysfunctions, Psychological] this term only 504

#14 MeSH descriptor: [Somatoform Disorders] explode all trees 945

#15 MeSH descriptor: [Munchausen Syndrome] explode all trees 2

#16 MeSH descriptor: [Trauma and Stressor Related Disorders] explode all trees 4508

#17 MeSH descriptor: [Trauma and Stressor Related Disorders] explode all trees 4508

#18 MeSH descriptor: [Impulsive Behavior] explode all trees 1672

#19 MeSH descriptor: [Hoarding] this term only 7

#20 MeSH descriptor: [Self-Injurious Behavior] explode all trees 2427

#21 (“anorexia nervosa” or “binge eating disorder” or bulimia or “eating disorder” or automutilation or “suicidal behaviour” or “self poisoning” or “suicidal ideation” or suicide or “suicide attempt” or depression or “agitated depression” or “atypical depression” or “chronic depression” or “depressive psychosis” or dysphoria or dysthymia or “endogenous depression” or involutional or “late life depression” or “major depression” or melancholia or “minor depression” or “mourning syndrome” or “organic depression” or “perinatal depression” or “antenatal depression” or “postnatal depression” or “post-stroke depression” or “postoperative depression” or “premenstrual dysphoric disorder” or “reactive depression” or “recurrent brief depression” or “seasonal affective disorder” or “treatment resistant depression” or neurosis or

“affective neurosis” or “anxiety neurosis” or dysthymia or hysteria or neurasthenia or psychasthenia or “adjustment disorder” or “anxiety disorder” or “acute stress disorder” or “generalized anxiety disorder” or panic or “posttraumatic stress disorder” or “separation anxiety” or “obsessive compulsive disorder” or compulsion or obsession or phobia or agoraphobia or claustrophobia or neophobia or “social phobia” or “somatoform disorder” or “body dysmorphic disorder” or “conversion disorder” or “delusional pregnancy” or hypochondriasis or “masked depression” or “psychogenic pain” or somatization or “mood disorder” or “affective neurosis” or “affective psychosis” or “blunted affect” or “major affective disorder” or “minor affective disorder” or “munchausen syndrome by proxy” or “munchausen syndrome” or “psychosexual disorder” or kleptomania or “trichotillomania” or “emotional disorder”):kw 63877

#22 (“MENTAL DISORDERS” or anhedonia or neurosis or “ACUTE STRESS DISORDER” or “adjustment disorders” or “attachment disorders” or “disinhibited social engagement disorder” or “posttraumatic stress disorder” or “complex ptsd” or desnos or “acute stress disorder” or “post-traumatic stress” or “traumatic neurosis” or “emotional trauma” or “AFFECTIVE DISORDERS” or “disruptive mood dysregulation disorder” or “dysthymic disorder” or “seasonal affective disorder” or “major depression” or “anaclitic depression” or “dysthymic disorder” or “endogenous depression” or “late life depression” or “postpartum depression” or “reactive depression” or “recurrent depression” or “treatment resistant depression” or “premenstrual dysphoric disorder” or “ANXIETY DISORDERS” or “generalized anxiety disorder” or “obsessive compulsive disorder” or “panic attack” or “panic disorder” or phobias or trichotillomania or acrophobia or agoraphobia or claustrophobia or ophidiophobia or “social phobia” or “separation anxiety disorder” or “EATING DISORDERS” or “anorexia nervosa” or “binge eating disorder” or bulimia or “SELF-INJURIOUS BEHAVIOR” or “self-destructive behavior” or “self-inflicted wounds” or “self-mutilation” or “self-poisoning” or suicide or “attempted suicide” or suicidality or “suicidal ideation” or “suicide prevention” or “SOMATOFORM DISORDERS” or “body dysmorphic disorder” or hypochondriasis or hysteria or neurasthenia or neurodermatitis or “somatization disorder” or “somatoform pain disorder” or “munchausen syndrome” or “munchausen syndrome by proxy” or (depression near/2 emotion) or "stress and trauma related disorders" or “mental health” or “mental health services” or “community mental health services” or “community counseling” or “community psychiatry”):kw45016

#23 ("acute stress" or (adjustment next disorder*) or ADNOS or (affective next disorder*) or agoraphobi* or "anorexia nervosa" or anxiety or astheni* or (attachment next disorder*) or (binge next eat*) or binging or (body next dysmorphi*) or bulimi* or (combat next disorder*) or (obsessive next compulsi*) or depression or depressed or depressive or dyssomni* or dyspareunia* or dysphori* or dysthymi* or dystoni* or (eating next disorder*) or EDNOS or (emotional next trauma) or fear or (health next anxiety) or hoarding or hyperactivity or hypochondri* or hysteri* or (medically next unexplained) or malingering or MDD or (common next mental) or (mental* near/2 (health or well*)) or mood or moods or munchausen or MUPS or mutism or neurastheni* or neurotic or neuros* or panic or phobi* or PND or ((post-trauma* or posttrauma*) next stress*) or psychogenic or psychosomatic or PTSD or (self next (injur* or harm or mutilat*)) or psychosexual or (psychological near/3 sexual near/3 dysfunction*) or "social anxiety" or somati* or somatoform or suicid* or parasuicid* or trichotillomani*):ti,ab 196731

#24 (depress* NEAR (employment or employability or unemployment or "return to work" or vocation* or absenteeism or presenteeism or "job security" or "work related" or "work focussed")):ab 241

#25 (#1 OR #2 OR #3 OR #4 OR #5 OR #6 OR #7 OR #8 OR #9 OR #10 OR #11 OR #12 OR #13 OR #14 OR #15 OR #16 OR #17 OR #18 OR #19 OR #20 OR #21 OR #22 OR #23 OR #24) 221261

#26 SR-SCIHZ OR HS-SCHIZ 941

#27 (bipolar or cyclothymi* or mania or manic or hypermani* or “rapid cycling” or (conversion next disorder*) or (dissociative next (amnesi* or fugue* or disorder*)) or "borderline state" or catatoni* or "character disorder" or delusion* or (capgras next syndrom*) or (diogenes next syndrom*) or depersonalization or depersonalisation or de-personalization or de-personalisation or (perceptual next disorder*) or (personality next disorder*) or BPD or paranoi* or psychiatr* or psychopathol* or psycho-pathol* or psychotic or psychosis* or psychoses* or psychotic* or schizo* or hebephreni* or “serious mental” or SMI):ti,ab,kw 69764

#28 MeSH descriptor: [Schizophrenia Spectrum and Other Psychotic Disorders] explode all trees 12178

#29 MeSH descriptor: [Personality Disorders] explode all trees 1828

#30 MeSH descriptor: [Bipolar and Related Disorders] explode all trees 3561

#31 (#26 OR #27 OR #28 OR #29 OR #30) 70436

#32 (loneliness or lonely):ti,ab,kw 1395

#33 ((social* or societ* or communit*) near (isolated or isolation)):ti,ab,kw 1724

#34 ((alleviat* or ease or manag* or mitigat* or prevent* or overcom* or reduc* or stop*) near/2 (isolation or isolated)):ti,ab,kw 324

#35 ((address* or enhanc* or improv* or increas* or promot* or target*) near/2 (inclusion or inclusivity)):ti,ab,kw 673

#36 (#32 or #33 or #34 or #35) 3739

#37 (#25 and #36) with Cochrane Library publication date Between Jul 2020 and Feb 2024, in Trials 916

#38 (#31 and #36) with Cochrane Library publication date Between Jul 2020 and Feb 2024, in Trials 173

#39 (“social alienation” or “social inclusion” or “social participation”):ti,ab,kw 1202

#40 (“community involvement” or “social support” or “social network” or “psychosocial environment” or “psychosocial rehabilitation”):ti,kw 7825

#41 (abandonment or alienation):ti,kw 77

#42 ((social or societ*or community) next (confine* or contact or contacts or connect* or inclusion or network* or participation or relations*)):ti,kw 2725

#43 ((social* or societ* or communit*) near/3 (alienat* or discriminat* or excluded or exclusion)):ti,ab,kw 459

#44 ((alleviat* or ease or manag* or mitigat* or prevent* or overcom* or reduc* or stop*) near/2 (exclusion or excluded)):ti,ab,kw 135

#45 ((social* or societ* or communit*) next network*):ti,ab,kw 2626

#46 ((social or societ* or psychosocial) adj support*):ti,ab,kw 21

#47 ("social capital" or (social* next mobil*)):ti,ab,kw 239

#48 (navigator or navigators):ti,kw 338

#49 ((peer or peers) next support*):ti,kw 604

#50 (anti-stigma* or ((intervention* or alleviat* or prevent* or reduc* or stop*) near/2 stigma*)):ti,ab,kw 944

#51 ("social learning theory" or ((social near interaction*) next (counsel* or educat* or intervention* or program* or therap* or train*))):ti,ab,kw 375

#52 (#39 OR #40 OR #41 OR #42 OR #43 OR #44 OR #45 OR #46 OR #47 OR #48 OR #50 OR #51) 13310

#53 (#31 AND #52) with Cochrane Library publication date Between Jul 2020 and Feb 2024, in Trials444

#54 ("social alienation" or "social inclusion" or "social participation"):ti kw 0

#55 ("community involvement" or "social network" or "psychosocial environment" or "psychosocial rehabilitation"):ti,kw 1045

#56 (abandonment or alienation):ti,kw 77

#57 ((chang* or develop* or enhanc* or initiative* or intervention* or program* or mitigat* or address* or improv* or target*) NEAR ((social or societ*or community) next (confine* or contact or contacts or connect* or inclusion or network* or participation or relations*))):ti,ab,kw 1563

#58 ((social* or societ* or communit*) near/3 (alienat* or discriminat* or excluded or exclusion)):ti,ab,kw 459

#59 ((alleviat* or ease or manag* or mitigat* or prevent* or overcom* or reduc* or stop*) near/2 (exclusion or excluded)):ti,ab,kw 135

#60 ((social* or societ* or communit*) next network*):ti,kw 1356

#61 ((social or societ* or psychosocial) next support*):ti 980

#62 ("social capital" or (social* next mobil*)):ti,ab,kw 239

#63 (navigator or navigators):ti,kw 338

#64 ((peer or peers) next support*):ti,kw 604

#65 (anti-stigma* or ((intervention* or alleviat* or prevent* or reduc* or stop*) near/2 stigma*)):ti,ab,kw 944

#66 ("social learning theory" or ((social near interaction*) next (counsel* or educat* or intervention* or program* or therap* or train*))):ti,ab,kw 375

#67 (#54 OR #55 OR #56 OR #57 OR #58 OR #59 OR #60 OR #61 OR #62 OR #63 OR #64 OR #65 OR #66) 6548

#68 (#31 AND #67) with Cochrane Library publication date Between Jul 2020 and Feb 2024, in Trials338

**Search Name: CCRT_Barnett_Social Participation SMI**

Date Run: 28/02/2024 10:55:56

Comment:

ID Search Hits

#1 SR-DEPRESSN OR HS-DEPRESSN 35371

#2 MeSH descriptor: [Mental Disorders] this term only 5377

#3 MeSH descriptor: [Anxiety Disorders] explode all trees 10069

#4 MeSH descriptor: [Trichotillomania] this term only 96

#5 MeSH descriptor: [Feeding and Eating Disorders] this term only 1185

#6 MeSH descriptor: [Anorexia Nervosa] this term only 754

#7 MeSH descriptor: [Binge-Eating Disorder] this term only 482

#8 MeSH descriptor: [Bulimia Nervosa] this term only 362

#9 MeSH descriptor: [Mood Disorders] this term only 1127

#10 MeSH descriptor: [Depressive Disorder] explode all trees 16414

#11 MeSH descriptor: [Premenstrual Dysphoric Disorder] this term only 51

#12 MeSH descriptor: [Reactive Attachment Disorder] this term only 24

#13 MeSH descriptor: [Sexual Dysfunctions, Psychological] this term only 504

#14 MeSH descriptor: [Somatoform Disorders] explode all trees 945

#15 MeSH descriptor: [Munchausen Syndrome] explode all trees 2

#16 MeSH descriptor: [Trauma and Stressor Related Disorders] explode all trees 4508

#17 MeSH descriptor: [Trauma and Stressor Related Disorders] explode all trees 4508

#18 MeSH descriptor: [Impulsive Behavior] explode all trees 1672

#19 MeSH descriptor: [Hoarding] this term only 7

#20 MeSH descriptor: [Self-Injurious Behavior] explode all trees 2427

#21 (“anorexia nervosa” or “binge eating disorder” or bulimia or “eating disorder” or automutilation or “suicidal behaviour” or “self poisoning” or “suicidal ideation” or suicide or “suicide attempt” or depression or “agitated depression” or “atypical depression” or “chronic depression” or “depressive psychosis” or dysphoria or dysthymia or “endogenous depression” or involutional or “late life depression” or “major depression” or melancholia or “minor depression” or “mourning syndrome” or “organic depression” or “perinatal depression” or “antenatal depression” or “postnatal depression” or “post-stroke depression” or “postoperative depression” or “premenstrual dysphoric disorder” or “reactive depression” or “recurrent brief depression” or “seasonal affective disorder” or “treatment resistant depression” or neurosis or

“affective neurosis” or “anxiety neurosis” or dysthymia or hysteria or neurasthenia or psychasthenia or “adjustment disorder” or “anxiety disorder” or “acute stress disorder” or “generalized anxiety disorder” or panic or “posttraumatic stress disorder” or “separation anxiety” or “obsessive compulsive disorder” or compulsion or obsession or phobia or agoraphobia or claustrophobia or neophobia or “social phobia” or “somatoform disorder” or “body dysmorphic disorder” or “conversion disorder” or “delusional pregnancy” or hypochondriasis or “masked depression” or “psychogenic pain” or somatization or “mood disorder” or “affective neurosis” or “affective psychosis” or “blunted affect” or “major affective disorder” or “minor affective disorder” or “munchausen syndrome by proxy” or “munchausen syndrome” or “psychosexual disorder” or kleptomania or “trichotillomania” or “emotional disorder”):kw 63877

#22 (“MENTAL DISORDERS” or anhedonia or neurosis or “ACUTE STRESS DISORDER” or “adjustment disorders” or “attachment disorders” or “disinhibited social engagement disorder” or “posttraumatic stress disorder” or “complex ptsd” or desnos or “acute stress disorder” or “post-traumatic stress” or “traumatic neurosis” or “emotional trauma” or “AFFECTIVE DISORDERS” or “disruptive mood dysregulation disorder” or “dysthymic disorder” or “seasonal affective disorder” or “major depression” or “anaclitic depression” or “dysthymic disorder” or “endogenous depression” or “late life depression” or “postpartum depression” or “reactive depression” or “recurrent depression” or “treatment resistant depression” or “premenstrual dysphoric disorder” or “ANXIETY DISORDERS” or “generalized anxiety disorder” or “obsessive compulsive disorder” or “panic attack” or “panic disorder” or phobias or trichotillomania or acrophobia or agoraphobia or claustrophobia or ophidiophobia or “social phobia” or “separation anxiety disorder” or “EATING DISORDERS” or “anorexia nervosa” or “binge eating disorder” or bulimia or “SELF-INJURIOUS BEHAVIOR” or “self-destructive behavior” or “self-inflicted wounds” or “self-mutilation” or “self-poisoning” or suicide or “attempted suicide” or suicidality or “suicidal ideation” or “suicide prevention” or “SOMATOFORM DISORDERS” or “body dysmorphic disorder” or hypochondriasis or hysteria or neurasthenia or neurodermatitis or “somatization disorder” or “somatoform pain disorder” or “munchausen syndrome” or “munchausen syndrome by proxy” or (depression near/2 emotion) or "stress and trauma related disorders" or “mental health” or “mental health services” or “community mental health services” or “community counseling” or “community psychiatry”):kw45016

#23 ("acute stress" or (adjustment next disorder*) or ADNOS or (affective next disorder*) or agoraphobi* or "anorexia nervosa" or anxiety or astheni* or (attachment next disorder*) or (binge next eat*) or binging or (body next dysmorphi*) or bulimi* or (combat next disorder*) or (obsessive next compulsi*) or depression or depressed or depressive or dyssomni* or dyspareunia* or dysphori* or dysthymi* or dystoni* or (eating next disorder*) or EDNOS or (emotional next trauma) or fear or (health next anxiety) or hoarding or hyperactivity or hypochondri* or hysteri* or (medically next unexplained) or malingering or MDD or (common next mental) or (mental* near/2 (health or well*)) or mood or moods or munchausen or MUPS or mutism or neurastheni* or neurotic or neuros* or panic or phobi* or PND or ((post-trauma* or posttrauma*) next stress*) or psychogenic or psychosomatic or PTSD or (self next (injur* or harm or mutilat*)) or psychosexual or (psychological near/3 sexual near/3 dysfunction*) or "social anxiety" or somati* or somatoform or suicid* or parasuicid* or trichotillomani*):ti,ab 196731

#24 (depress* NEAR (employment or employability or unemployment or "return to work" or vocation* or absenteeism or presenteeism or "job security" or "work related" or "work focussed")):ab 241

#25 (#1 OR #2 OR #3 OR #4 OR #5 OR #6 OR #7 OR #8 OR #9 OR #10 OR #11 OR #12 OR #13 OR #14 OR #15 OR #16 OR #17 OR #18 OR #19 OR #20 OR #21 OR #22 OR #23 OR #24) 221261

#26 SR-SCIHZ OR HS-SCHIZ 941

#27 (bipolar or cyclothymi* or mania or manic or hypermani* or “rapid cycling” or (conversion next disorder*) or (dissociative next (amnesi* or fugue* or disorder*)) or "borderline state" or catatoni* or "character disorder" or delusion* or (capgras next syndrom*) or (diogenes next syndrom*) or depersonalization or depersonalisation or de-personalization or de-personalisation or (perceptual next disorder*) or (personality next disorder*) or BPD or paranoi* or psychiatr* or psychopathol* or psycho-pathol* or psychotic or psychosis* or psychoses* or psychotic* or schizo* or hebephreni* or “serious mental” or SMI):ti,ab,kw 69764

#28 MeSH descriptor: [Schizophrenia Spectrum and Other Psychotic Disorders] explode all trees 12178

#29 MeSH descriptor: [Personality Disorders] explode all trees 1828

#30 MeSH descriptor: [Bipolar and Related Disorders] explode all trees 3561

#31 (#26 OR #27 OR #28 OR #29 OR #30) 70436

#32 (loneliness or lonely):ti,ab,kw 1395

#33 ((social* or societ* or communit*) near (isolated or isolation)):ti,ab,kw 1724

#34 ((alleviat* or ease or manag* or mitigat* or prevent* or overcom* or reduc* or stop*) near/2 (isolation or isolated)):ti,ab,kw 324

#35 ((address* or enhanc* or improv* or increas* or promot* or target*) near/2 (inclusion or inclusivity)):ti,ab,kw 673

#36 (#32 or #33 or #34 or #35) 3739

#37 (#25 and #36) with Cochrane Library publication date Between Jul 2020 and Feb 2024, in Trials 916

#38 (#31 and #36) with Cochrane Library publication date Between Jul 2020 and Feb 2024, in Trials 173

#39 (“social alienation” or “social inclusion” or “social participation”):ti,ab,kw 1202

#40 (“community involvement” or “social support” or “social network” or “psychosocial environment” or “psychosocial rehabilitation”):ti,kw 7825

#41 (abandonment or alienation):ti,kw 77

#42 ((social or societ*or community) next (confine* or contact or contacts or connect* or inclusion or network* or participation or relations*)):ti,kw 2725

#43 ((social* or societ* or communit*) near/3 (alienat* or discriminat* or excluded or exclusion)):ti,ab,kw 459

#44 ((alleviat* or ease or manag* or mitigat* or prevent* or overcom* or reduc* or stop*) near/2 (exclusion or excluded)):ti,ab,kw 135

#45 ((social* or societ* or communit*) next network*):ti,ab,kw 2626

#46 ((social or societ* or psychosocial) adj support*):ti,ab,kw 21

#47 ("social capital" or (social* next mobil*)):ti,ab,kw 239

#48 (navigator or navigators):ti,kw 338

#49 ((peer or peers) next support*):ti,kw 604

#50 (anti-stigma* or ((intervention* or alleviat* or prevent* or reduc* or stop*) near/2 stigma*)):ti,ab,kw 944

#51 ("social learning theory" or ((social near interaction*) next (counsel* or educat* or intervention* or program* or therap* or train*))):ti,ab,kw 375

#52 (#39 OR #40 OR #41 OR #42 OR #43 OR #44 OR #45 OR #46 OR #47 OR #48 OR #50 OR #51) 13310

#53 (#31 AND #52) with Cochrane Library publication date Between Jul 2020 and Feb 2024, in Trials444

#### Full reference list for studies included in the Systematic Review

1. Adamus, C., Mötteli, S., Jäger, M., & Richter, D. (2022). Independent Supported Housing for non-homeless individuals with severe mental illness: Comparison of two effectiveness studies using a randomised controlled and an observational study design. *Frontiers in Psychiatry*, *13*, 1033328. https://doi.org/10.3389/fpsyt.2022.1033328
2. Agrest, M., Le, P., & LH, Y. (2019). Implementing a community-based task-shifting psychosocial intervention for individuals with psychosis in Chile: Perspectives from users. *Int J Soc Psychiatry*, *65*, 38–45.
3. Agudelo-Hernández, F., Guapacha-Montoya, M., & Rojas-Andrade, R. (2024). Mutual Aid Groups for Loneliness, Psychosocial Disability, and Continuity of Care. *Community Mental Health Journal*, *60*(3), 608–619. https://doi.org/10.1007/s10597-023-01216-9
4. Aivalioti, E. I., Simos, P., Basta, M., & Vgontzas, A. N. (2022). Brief Solution Focused Therapy on schizophrenia: A preliminary study of family characteristics and psychopathology. *Psychiatriki*.https://doi.org/10.22365/jpsych.2022.085
5. Albers, W. M. M., Roeg, D. P. K., Nijssen, Y. A. M., Deen, M. L., Bongers, I. M. B., & Van Weeghel, J. (2021). Intervention to prevent and manage the effects of

victimization related to social participation for people with severe mental illness: Results from a cluster randomized controlled trial. *Psychiatric Rehabilitation Journal*, *44*(3), 254–265. https://doi.org/10.1037/prj0000449
6. Al-HadiHasan, A., Callaghan, P., & Lymn, J. S. (2017). Qualitative process evaluation of a psycho-educational intervention targeted at people diagnosed with schizo-phrenia and their primary caregivers in Jordan. *BMC Psychiatry*, *17*(68).
7. Ali, A., McKenzie, E., Hassiotis, A., Priebe, S., Lloyd-Evans, B., Jones, R., Panca, M., Omar, R., Finning, S., Moore, S., Roe, C., & King, M. (2021). A pilot randomised controlled trial of befriending by volunteers in people with intellectual disability and depressive symptoms. *Journal of Intellectual Disability Research*, *65*(11), 1010–1019. https://doi.org/10.1111/jir.12886
8. Aubry, T., Bourque, J., & P, G. (2019). A randomised controlled trial of the effectiveness of Housing First in a small Canadian city. *BMC Public Health*, *19*(1154).
9. Aubry, T., Goering, P., Veldhuizen, S., Adair, C. E., Bourque, J., & Distasio, J. (2016). A multiple-city RCT of housing first with assertive community treatment for homeless Canadians with serious mental illness. *Psychiatr Serv*, *67*(3), 275–281.
10. Baller, J. B., Blyler, C. R., Bronnikov, S., Xie, H., Bond, G. R., Filion, K., & Hale, T. (2020). Long-Term Follow-Up of a Randomized Trial of Supported Employment for SSDI Beneficiaries With Mental Illness. *Psychiatric Services*, *71*(3), 243–249. https://doi.org/10.1176/appi.ps.201800554
11. Battle, C. L., Cardemil, E. V., Rossi, R., O'Hara, M. W., & Miller, I. W. (2023). Family treatment for postpartum depression: Acceptability, feasibility, and preliminary clinical outcomes. *Archives of Women's Mental Health*, *26*(1), 127–134. https://doi.org/10.1007/s00737-022-01282-0
12. Beavan, V., Jager, A., & Santos, B. (2017). Do peer-support groups for voice-hearers work? A small scale study of Hearing Voices Network support groups in Australia. *Psychosis*, *9*, 57–66.
13. Bejerholm, U., Larsson, M. E., & Johanson, S. (2017). Supported employment adapted for people with affective disorders—A randomized controlled trial. *J Affect Disord*, *207*, 212–220.
14. Bell, M., Bryson, G., Greig, T., Fiszdon, J., & Wexler, B. (2005). Neurocognitive enhancement therapy with work therapy: Productivity outcomes at 6-and 12-month follow-ups. *J Rehabil Res Dev*, *42*(6).
15. Bell, M., Laws, H., Pittman, B., & Johannesen, J. (2018). Comparison of focused cognitive training and portable "brain-games" on functional outcomes for vocational rehabilitation participants. *Sci Rep*, *8*(1), 1–9.

16. Bell, M., Lysaker, P., & Bryson, G. (2003). A behavioral intervention to improve work performance in schizophrenia: Work behavior inventory feedback. *J Vocat Rehabil*, *18*(1), 43–50.
17. Bell, M., Milstein, R., & Lysaker, P. (1993). Pay as an incentive in work participation by patients with severe mental illness. *Psychiatr Serv*, *44*(7), 684–686.
18. Bell, M., Zito, W., Greig, T., & Wexler, B. (2008). Neurocognitive enhancement therapy with vocational services: Work outcomes at two-year follow-up. *Schizophr Res*, *105*(1–3), 18–29.
19. Beutel, M. E., Zwerenz, R., Bleichner, F., Vorndran, A., Gustson, D., & Knickenberg, R. J. (2005). Vocational training integrated into inpatient psychosomatic rehabilitation–short and long-term results from a controlled study. *Disabil Rehabil*, *27*(15), 891–900.
20. Bitter, N., Roeg, D., & M, A. (2017). How effective is the comprehensive approach to rehabilitation (CARe) methodology? A cluster randomized con-trolled trial. *BMC Psychiatry*, *17*(396).
21. Bjørkedal, S.-T. B., Bejerholm, U., Hjorthøj, C., Møller, T., & Eplov, L. F. (2023). Meaningful Activities and Recovery (MA&R): A co-led peer occupational therapy intervention for people with psychiatric disabilities. Results from a randomized controlled trial. *BMC Psychiatry*, *23*(1), 406. https://doi.org/10.1186/s12888-023-04875-w
22. Blajeski, S. M., Smith, M. J., Harrington, M. M., Johnson, J. M., Oulvey, E. A., Mueser, K. T., McGurk, S. R., & Razzano, L. A. (2023). Critical elements in the experience of virtual reality job interview training for unemployed individuals with serious mental illness: Implications for IPS supported employment. *Psychiatric Rehabilitation Journal*, *46*(4), 353–359. https://doi.org/10.1037/prj0000574
23. Blajeski, S., Smith, M. J., Harrington, M., Johnson, J., Ross, B., Weaver, A., Razzano, L. A., Pashka, N., Brown, A., Prestipino, J., Nelson, K., Lieberman, T., Jordan, N., Oulvey, E. A., Mueser, K. T., McGurk, S. R., Bell, M. D., & Smith, J. D. (2024). A Mixed-Methods Implementation Evaluation of Virtual Reality Job Interview Training in IPS Supported Employment. *Psychiatric Services*, *75*(3), 228–236. https://doi.org/10.1176/appi.ps.20230023
24. Boevink, W., Kroon, H., Vugt, M., Delespaul, P., & Os, J. (2016). A user-developed, user run recovery programme for people with severe mental illness: A randomised control trial. *Psychosis*, *8*(4), 287–300.
25. Brown, M., Jason, L., & D, M. (2016). Housing first as an effective model for community stabilization among vulnerable individuals with chronic and nonchronic homelessness histories. *J Community Psychol*, *44*, 384–390.
26. Burnam, M. A., Morton, S. C., McGlynn, E. A., Petersen, L. P., Stecher, B. M., & Hayes, C. (1996). An experimental evaluation of residential and nonresidential treatment for dually diagnosed homeless adults. *J Addict Dis*, *14*(4), 111–134.

27. Burns, A. M. N., & Erickson, D. H. (2023). Adding Cognitive Remediation to Employment Support Services: A Randomized Controlled Trial. *Psychiatric Services*, *74*(3), 222–228. https://doi.org/10.1176/appi.ps.202100249
28. Caplan, R. A., Nelson, G., Distasio, J., Isaak, C., Edel, B., Macnaughton, E., Piat, M., Patterson, M., Kirst, M., Aubry, T., Stergiopoulos, V., & Goering, P. (2023). Parent–child relationship outcomes in a randomized controlled trial of housing first for indigenous and non-Indigenous parents experiencing homelessness, mental illness, and separation from their children. *Psychiatric Rehabilitation Journal*, *46*(4), 335–342. https://doi.org/10.1037/prj0000575
29. Castelein, S., Bruggeman, R., Busschbach, J. T., Gaag, M., Stant, A., & Knegtering, H. (2008). The effectiveness of peer support groups in psychosis: A randomized controlled trial. *Acta Psychiatr Scand*, *118*(1), 64–72.
30. Cella, M., Sedgwick, O., Lawrence, M., Grant, N., Tsapekos, D., Harrison, L., & Wykes, T. (2023). Evaluating the mechanisms of social cognition intervention in schizophrenia: A proof-of-concept trial. *Psychiatry Research*, *319*, 114963. https://doi.org/10.1016/j.psychres.2022.114963
31. Cervello, S., Dubreucq, J., Trichanh, M., Dubrulle, A., Amado, I., Bralet, M. C., Chirio-Espitalier, M., Delille, S., Fakra, E., Francq, C., Guillard-Bouhet, N., Graux, J., Lançon, C., Zakoian, J. M., Gauthier, E., Demily, C., & Franck, N. (2021). Cognitive remediation and professional insertion of people with schizophrenia: RemedRehab, a randomized controlled trial. *European Psychiatry*, *64*(1), e31. https://doi.org/10.1192/j.eurpsy.2021.25
32. Chandler, D. W., & Spicer, G. (2006). Integrated treatment for jail recidivists with co-occurring psychiatric and substance use disorders. *Community Ment Health J*, *42*(4), 405–425.
33. Chaudhry, N., Sattar, R., Kiran, T., Wan, M. W., Husain, M., Hidayatullah, S., Ali, B., Shafique, N., Suhag, Z., Saeed, Q., Maqbool, S., & Husain, N. (2023). Supporting Depressed Mothers of Young Children with Intellectual Disability: Feasibility of an Integrated Parenting Intervention in a Low-Income Setting. *Children*, *10*(6), 913. https://doi.org/10.3390/children10060913
34. Chen, Y., Yau, E., & C, L. (2020). A 6-month randomized controlled pilot study on the effects of the clubhouse model of psychosocial rehabilitation with Chinese individuals with schizophrenia. *Adm Policy Ment Health*, *47*, 107–114.
35. Christensen, T. N., Wallstrøm, I. G., Bojesen, A. B., Nordentoft, M., & Eplov, L. F. (2021). Predictors of work and education among people with severe mental illness who participated in the Danish individual placement and support study: Findings from a randomized clinical trial. *Social Psychiatry and Psychiatric Epidemiology*, *56*(9), 1669–1677. https://doi.org/10.1007/s00127-021-02107-8
36. Christensen, T. N., Wallstrøm, I. G., Stenager, E., Bojesen, A. B., Gluud, C., & Nordentoft, M. (2019). Effects of individual placement and support supplemented with cognitive remediation and work-focused social skills training

for people with severe mental illness: A randomized clinical trial. *JAMA Psychiat*, *76*(12), 1232–1240.

37. Compton, M. T., Kelley, M. E., Anderson, S., Ellis, S., Graves, J., Broussard, B., Pauselli, L., Zern, A., Pope, L. G., Johnson, M., & Haynes, N. L. (2023). Opening Doors to Recovery: A Randomized Controlled Trial of a Recovery-Oriented Community Navigation Service for Individuals With Serious Mental Illnesses and Repeated Hospitalizations. *The Journal of Clinical Psychiatry*, *84*(2).https://doi.org/10.4088/JCP.22m14498
38. Conoley, C. W., & Garber, R. A. (1985). Effects of reframing and self-control directives on loneliness, depression, and controllability. *J Couns Psychol*, *32*(1).
39. Cook, J., Blyler, C., Burke-Miller, J., McFarlane, W., Leff, H., & Mueser, K. (2008). Effectiveness of supported employment for individuals with schizophrenia: Results of a multi-site, randomized trial. *Clin Schizophr Relat Psychoses*, *2*(1), 37–46.
40. Cook, J., Burke-Miller, J., & Roessel, E. (2016). Long-term effects of evidence-based supported employment on earnings and on SSI and SSDI participation among individuals with psychiatric disabilities. *Am J Psychiatry*, *173*, 1007–1014.
41. Cosden, M., Ellens, J., Schnell, J., & Yamini-Diouf, Y. (2005). Efficacy of a mental health treatment court with assertive community treatment. *Behav Sci Law*, *23*(2), 199–214.
42. Creech, S. K., Pearson, R., Saenz, J. J., Braciszewski, J. M., Riggs, S. A., & Taft, C. T. (2022). Pilot trial of Strength at Home Parents: A trauma-informed parenting support treatment for veterans. *Couple and Family Psychology: Research and Practice*, *11*(3), 205–216. https://doi.org/10.1037/cfp0000232
43. Cusack, K. J., Morrissey, J. P., Cuddeback, G. S., Prins, A., & Williams, D. M. (2010). Criminal justice involvement, behavioral health service use, and costs of forensic assertive community treatment: A randomized trial. *Community Ment Health J*, *46*(4), 356–363.
44. Dabit, S., Quraishi, S., Jordan, J., & Biagianti, B. (2021). Improving social functioning in people with schizophrenia-spectrum disorders via mobile experimental interventions: Results from the CLIMB pilot trial. *Schizophrenia Research: Cognition*, *26*, 100211. https://doi.org/10.1016/j.scog.2021.100211
45. Danielsson, L., Waern, M., Hensing, G., & Holmgren, K. (2020). Work-directed rehabilitation or physical activity to support work ability and mental health in common mental disorders: A pilot randomized controlled trial. *Clinical Rehabilitation*, *34*(2), 170–181. https://doi.org/10.1177/0269215519880230
46. Dark, F., Scott, J. G., Baker, A., Parker, S., Gordon, A., Newman, E., Gore-Jones, V., Lim, C. C. W., Jones, L., & Penn, D. L. (2020). Randomized controlled trial of social cognition and interaction training compared to befriending group. *British Journal of Clinical Psychology*, *59*(3), 384–402. https://doi.org/10.1111/bjc.12252

47. Davidson, L., Shahar, G., Stayner, D. A., Chinman, M. J., Rakfeldt, J., & Tebes, J. K. (2004). Supported socialization for people with psychiatric disabilities: Lessons from a randomized controlled trial. *J Community Psychol*, *32*(4), 453–477.
48. Davis, L., Kyriakides, T., Suris, A., Ottomanelli, L., Mueller, L., & Parker, P. (2018). Effect of evidence-based supported employment vs transitional work on achieving steady work among veterans with posttraumatic stress disorder: A randomized clinical trial. *JAMA Psychiat*, *75*(4), 316–324.
49. Davis, L. L., Mumba, M. N., Toscano, R., Pilkinton, P., Blansett, C. M., McCall, K., MacVicar, D., & Bartolucci, A. (2022). A Randomized Controlled Trial Evaluating the Effectiveness of Supported Employment Integrated in Primary Care. *Psychiatric Services*, *73*(6), 620–627. https://doi.org/10.1176/appi.ps.202000926
50. Davis, L., Leon, A., Toscano, R., Drebing, C., Ward, L. C., & Parker, P. (2012). A randomized controlled trial of supported employment among veterans with post-traumatic stress disorder. *Psychiatr Serv*, *63*(5), 464–470.
51. Davis, L., Lysaker, P. H., Kristeller, J. L., Salyers, M. P., Kovach, A. C., & Woller, S. (2015). Effect of mindfulness on vocational rehabilitation outcomes in stable phase schizophrenia. *Psychol Serv*, *12*(3).
52. De Waal, M. M., Blankers, M., Lommerse, N. M., Kikkert, M. J., Dekker, J. J. M., & Goudriaan, A. E. (2021). Economic Evaluation of the SOS Training to Reduce Victimization in Dual Diagnosis Patients. *Journal of Dual Diagnosis*, *17*(4), 333–343. https://doi.org/10.1080/15504263.2021.1965409
53. De Waal, M. M., Dekker, J. J., Kikkert, M., Christ, C., Chmielewska, J., & Staats, M. (2019). Other-wise, Streetwise (SOS) training, an intervention to prevent victimization in dual-diagnosis patients: Results from a randomized clinical trial. *Addiction*, *114*(4), 730–740.
54. de Weerd, B. J., van Dijk, van der Linden, J. N., Roelen, J. N., & Verbraak, M. (2016). The effectiveness of a convergence dialogue meeting with the employer in promoting return to work as part of the cognitive-behavioural treatment of common mental disorders: A randomized controlled trial. *Work*, *54*(3), 647–655.
55. Dehn, L. B., Beblo, T., Richter, D., Wienberg, G., Kremer, G., Steinhart, I., & Driessen, M. (2022). Effectiveness of supported housing versus residential care in severe mental illness: A multicenter, quasi-experimental study. *Social Psychiatry and Psychiatric Epidemiology*, *57*(5), 927–937. https://doi.org/10.1007/s00127-021-02214-6
56. Doğu, S., Kayıhan, H., Kokurcan, A., & Örsel, S. (2021). The effectiveness of a combination of Occupational Therapy and Social Skills Training in people with schizophrenia: A rater-blinded randomized controlled trial. *British Journal of Occupational Therapy*, *84*(11), 684–693. https://doi.org/10.1177/03080226211022953
57. Doré-Gauthier, V., Miron, J., Jutras-Aswad, D., Ouellet-Plamondon, C., & Abdel-Baki, A. (2020). Specialized assertive community treatment intervention for

homeless youth with first episode psychosis and substance use disorder: A 2-year follow-up study. *Early Intervention in Psychiatry*, *14*(2), 203–210. https://doi.org/10.1111/eip.12846

58. Dubreucq, J., Gabayet, F., Ycart, B., Faraldo, M., Melis, F., Lucas, T., Arnaud, B., Bacconnier, M., Bakri, M., Cambier, G., Carmona, F., Chereau, I., Challe, T., Morel, S., Pires, S., Roussel, C., Lamy, P., Legrand, G., Pages, E., … the RemedRugby Group. (2020). Improving social function with real-world social-cognitive remediation in schizophrenia: Results from the RemedRugby quasi-experimental trial. *European Psychiatry*, *63*(1), e41. https://doi.org/10.1192/j.eurpsy.2020.42
59. Easter, M., Swanson, J., & A, R. (2021). Impact of psychiatric advance directive facilitation on mental health consumers: Empowerment, treatment attitudes and the role of peer support specialists. *J Mental Health*, *30*, 585–593.
60. Ebrahim, S., Glascott, A., & H, M. (2018). Recovery Colleges: How effective are they? *J Ment Health Train Educ Pract*, *13*, 209–218.
61. Edge, D., Degnan, A., & S, C. (2018). Culturally-adapted Family Intervention (CaFI) for African Caribbeans with schizophrenia and their families: A fea-sibility study of implementation and acceptability. *Health Serv Deliv Res*, *6*(32).
62. Elbogen, E. B., Hamer, R. M., Swanson, J. W., & Swartz, M. S. (2016). A randomized clinical trial of a money management intervention for veterans with psychiatric disabilities. *Psychiatr Serv*, *67*(10), 1142–1145.
63. Ellison, M. L., Schutt, R. K., Yuan, L.-H., Mitchell-Miland, C., Glickman, M. E., & McCarthy, S. (2020). Impact of peer specialist services on residential stability and behavioral health status among formerly homeless veterans with cooccurring mental health and substance use conditions. *Med Care*, *58*(4), 307–313.
64. Erickson, D. H., Roes, M. M., DiGiacomo, A., & Burns, A. (2021). Individual Placement and Support” boosts employment for early psychosis clients, even when baseline rates are high. *Early Interven Psychiatry*, *15*(3), 662–668.
65. Favrod, J., Nguyen, A., & A, T. (2019). Impact of positive emotion regulation training on negative symptoms and social functioning in schizophrenia: A field test. *Front Psychiatry*, *10*(532).
66. Finnes, A., Hoch, J. S., Enebrink, P., Dahl, J., Ghaderi, A., Nager, A., & Feldman, I. (2022). Economic evaluation of return-to-work interventions for mental disorder-related sickness absence: Two years follow-up of a randomized clinical trial. *Scandinavian Journal of Work, Environment & Health*, *48*(4), 264–272. https://doi.org/10.5271/sjweh.4012
67. Fitzpatrick, S., Wagner, A. C., Crenshaw, A. O., Varma, S., Whitfield, K. M., Valela, R., Di Bartolomeo, A. A., Fulham, L., Martin-Newnham, C., Mensah, D. H., Collins, A., Landy, M. S. H., Morland, L., Doss, B. D., & Monson, C. M. (2021). Initial outcomes of couple HOPES: A guided online couple intervention for PTSD

and relationship enhancement. *Internet Interventions*, *25*, 100423. https://doi.org/10.1016/j.invent.2021.100423

68. Fletcher, T. D., Cunningham, J. L., Calsyn, R. J., Morse, G. A., & Klinkenberg, W. D. (2008). Evaluation of treatment programs for dual disorder individuals: Modeling longitudinal and mediation effects. *Adm Policy Ment Health Ment Health Serv Res*, *35*(4), 319–336.
69. Fowler, D., Hodgekins, J., & French, P. (2019). Social recovery therapy in improving activity and social outcomes in early psychosis: Current evidence and longer term outcomes. *Schizophr Res*, *203*, 99–104.
70. Gammelgaard, I., Christensen, T., & L, E. (2017). I have potential": Experiences of recovery in the individual placement and support intervention. *Int J Soc Psychiatry*, *63*, 400–406.
71. Gelkopf, M., Sigal, M., & Kramer, R. (1994). Therapeutic use of humor to improve social support in an institutionalized schizophrenic inpatient community. *J Soc Psychol*, *134*(2), 175–182.
72. Gjengedal, R. G. H., Reme, S. E., Osnes, K., Lagerfeld, S. E., Blonk, R. W. B., Sandin, K., Berge, T., & Hjemdal, O. (2020). Work-focused therapy for common mental disorders: A naturalistic study comparing an intervention group with a waitlist control group. *Work*, *66*(3), 657–667. https://doi.org/10.3233/WOR-203208
73. Glynn, S. M., Marder, L., SR, RP, B., K, W., WC, W., & D.A. (2004). Supplementing clinic-based skills training with manual-based community support sessions: Effects on social adjustment of patients with schizophrenia. *Focus*, *159*(1), 829–877.
74. Glynn, S., Marder, S., & D, N. (2017). An RCT evaluating the effects of skills training and medication type on work outcomes among patients with schizophrenia. *Psychiatr Serv*, *68*, 271–277.
75. Goldfinger, S. M., Schutt, R. K., Tolomiczenko, G. S., Seidman, L., Penk, W. E., & Turner, W. (1999). Housing placement and subsequent days homeless among formerly homeless adults with mental illness. *Psychiatr Serv*, *50*(5), 674–679.
76. Granholm, E., McQuaid, M., JR, FS, A., LA, P., D, P., & P. (2005). A randomized, controlled trial of cognitive behavioral social skills training for middle-aged and older outpatients with chronic schizophrenia. *Am J Psychiatry*, *162*(3), 520–529.
77. Gutman, S. A., Kerner, R., Zombek, I., Dulek, J., & Ramsey, C. A. (2009). Supported education for adults with psychiatric disabilities: Effectiveness of an occupational therapy program. *Am J Occup Ther*, *63*(3), 245–254.
78. Gutman, S., & Raphael-Greenfield, E. (2017). Effectiveness of a supportive housing program for homeless adults with mental illness and substance use: A two-group controlled trial. *Br J Occup Ther*, *80*, 286–293.
79. Hall, T., Jordan, H., & L, R. (2018). A process and intermediate outcomes evaluation of an Australian recovery college. *J Rec Mental Health*, *1*, 7–20.

80. Hansen, H. G., Starzer, M., Nilsson, S. F., Hjorthøj, C., Albert, N., & Nordentoft, M. (2023). Clinical Recovery and Long-Term Association of Specialized Early Intervention Services vs Treatment as Usual Among Individuals With First-Episode Schizophrenia Spectrum Disorder: 20-Year Follow-up of the OPUS Trial. *JAMA Psychiatry*, *80*(4), 371. https://doi.org/10.1001/jamapsychiatry.2022.5164
81. Hanssen, E., Balvert, S., Oorschot, M., Borkelmans, K., Van Os, J., Delespaul, P., & Fett, A.-K. (2020). An ecological momentary intervention incorporating personalised feedback to improve symptoms and social functioning in schizophrenia spectrum disorders. *Psychiatry Research*, *284*, 112695. https://doi.org/10.1016/j.psychres.2019.112695
82. Harris, A. W., Kosic, T., Xu, J., Walker, C., Gye, W., & Hodge, A. R. (2017). Web-based cognitive remediation improves supported employment outcomes in severe mental illness: Randomized controlled trial. *JMIR Mental Health*, *4*(3).
83. Haslam, C., Cruwys, T., Chang, M. X.-L., Bentley, S. V., Haslam, S. A., & Dingle, G. A. (2019). GROUPS 4 HEALTH reduces loneliness and social anxiety in adults with psychological distress: Findings from a randomized controlled trial. *J Consult Clin Psychol*, *87*(9).
84. Hasson-Ohayon, I., Mashiach-Eizenberg, M., & A, L.-R. (2019). Ran-domized controlled trial of adjunctive social cognition and interaction training, adjunctive therapeutic alliance focused therapy, and treatment as usual among persons with serious mental illness. *Front Psychiatry*, *10*(364).
85. Hasson-Ohayon, I., Mashiach-Eizenberg, M., Avidan, M., Roberts, D. L., & Roe, D. (2014). Social cognition and interaction training: Preliminary results of an RCT in a community setting in Israel. *Psychiatr Serv*, *65*(4), 555–558.
86. Heatherington, L., Bonner, B., & D, R. (2019). Sustaining outcomes research in residential treatment: A 15-year study of the Gould Farm program. *Psychol Serv*, *16*, 675–686.
87. Hees, H. L., Vries, G., Koeter, M. W., & Schene, A. H. (2013). Adjuvant occupational therapy improves long-term depression recovery and return-to-work in good health in sick-listed employees with major depression: Results of a randomised controlled trial. *Occup Environ Med*, *70*(4), 252–260.
88. Hellström, L., Bech, P., Hjorthøj, C., Nordentoft, M., Lindschou, J., & Eplov, L. F. (2017). Effect on return to work or education of individual placement and support modified for people with mood and anxiety disorders: Results of a randomised clinical trial. *Occup Environ Med*, *74*(10), 717–725.
89. Henderson, C., Brohan, E., Clement, S., Williams, P., Lassman, F., & Schauman, O. (2013). Decision aid on disclosure of mental health status to an employer: Feasibility and outcomes of a randomised controlled trial. *Br J Psychiatry*, *203*(5), 350–357.

90. Herman, D. B., Conover, S., Gorroochurn, P., Hinterland, K., Hoepner, L., & Susser, E. S. (2011). Randomized trial of critical time intervention to prevent homelessness after hospital discharge. *Psychiatr Serv*, *62*(7), 713–719.
91. Higgins, A., Downes, C., & M, M. (2019). Family members' perspectives on the acceptability and impact of a co-facilitated information programme: The EOLAS mental health programme. *Ir J Psychol Med*, *10*(1017).
92. Higgins, A., Hevey, D., & F, B. (2018). Outcomes of a co-facilitation skills training programme for mental health service users, family members, and clinicians: The EOLAS project. *Int J Ment Health Nurs*, *27*, 911–921.
93. Himle, J. A., Bybee, D., Steinberger, E., Laviolette, W. T., Weaver, A., & Vlnka, S. (2014). Work-related CBT versus vocational services as usual for unemployed persons with social anxiety disorder: A randomized controlled pilot trial. *Behav Res Ther*, *63*, 169–176.
94. Holmås, T. H., Monstad, K., & Reme, S. E. (2021). Regular employment for people with mental illness – An evaluation of the individual placement and support programme. *Social Science & Medicine*, *270*, 113691. https://doi.org/10.1016/j.socscimed.2021.113691
95. Holmes, A., Carlisle, T., & Z, V. (2017). Housing First: Permanent supported accommodation for people with psychosis who have experienced chronic homelessness. *Australas Psychiatry*, *25*, 56–59.
96. Holt, C., Gentilleau, C., Gemmill, A. W., & Milgrom, J. (2021). Improving the mother-infant relationship following postnatal depression: A randomised controlled trial of a brief intervention (HUGS). *Archives of Women's Mental Health*, *24*(6), 913–923. https://doi.org/10.1007/s00737-021-01116-5
97. Horan, W., Dolinsky, M., & J, L. (2018). Social cognitive skills training for psychosis with community-based training exercises: A randomized controlled trial. *Schizophr Bull*, *44*, 1254–1266.
98. Hui, C. L. M., Wong, A. K. H., Ho, E. C. N., Lam, B. S. T., Hui, P. W. M., Tao, T. J., Chang, W. C., Chan, S. K. W., Lee, E. H. M., Suen, Y. N., Lam, M. M. L., Chiu, C. P. Y., Li, F. W. S., Leung, K. F., McGhee, S. M., Law, C. W., Chung, D. W. S., Yeung, W. S., Yiu, M. G. C., … Chen, E. Y. H. (2023). Effectiveness and optimal duration of early intervention treatment in adult-onset psychosis: A randomized clinical trial. *Psychological Medicine*, *53*(6), 2339–2351. https://doi.org/10.1017/S0033291721004189
99. Hurlburt, M. S., Hough, R. L., & Wood, P. A. (1996). Effects of substance abuse on housing stability of homeless mentally ill persons in supported housing. *Psychiatr Serv*, *47*(731).
100. Hutchinson, J., Gilbert, D., & R, P. (2018). Implementing supported employ-ment. Lessons from the making IPS Work Project. *Int J Env Res Pub Health*, *15*(1545).

101. Inman, J., Bannigan, K., & Akhurst, J. (2021). Occupational therapy and psychosis: POINTER feasibility study for a pragmatic clinical trial. *British Journal of Occupational Therapy*, *84*(9), 541–549. https://doi.org/10.1177/03080226211000257

102. Johnson, D. M., Zlotnick, C., Hoffman, L., Palmieri, P. A., Johnson, N. L., Holmes, S. C., & Ceroni, T. L. (2020). A Randomized Controlled Trial Comparing HOPE Treatment and Present-Centered Therapy in Women Residing in Shelter With PTSD From Intimate Partner Violence. *Psychology of Women Quarterly*, *44*(4), 539–553. https://doi.org/10.1177/0361684320953120

103. Karasz, A., Anne, S., Hamadani, J. D., & Tofail, F. (2021). The ASHA (Hope) Project: Testing an Integrated Depression Treatment and Economic Strengthening Intervention in Rural Bangladesh: A Pilot Randomized Controlled Trial. *International Journal of Environmental Research and Public Health*, *18*(1), 279. https://doi.org/10.3390/ijerph18010279

104. Kayo, M., Scemes, S., & M, S. (2020). A randomized controlled trial of social skills training for patients with treatment-resistant schizophrenia with pre-dominantly negative symptoms. *Psychiatry Res*, *287*(112914).

105. Kerman, N., Aubry, T., Adair, C. E., Distasio, J., Latimer, E., Somers, J., & Stergiopoulos, V. (2020). Effectiveness of Housing First for Homeless Adults with Mental Illness Who Frequently Use Emergency Departments in a Multisite Randomized Controlled Trial. *Administration and Policy in Mental Health and Mental Health Services Research*, *47*(4), 515–525. https://doi.org/10.1007/s10488-020-01008-3

106. Kern, R. S., Reddy, L. F., Horan, W. P., Glynn, S. M., Stigers, P. J., Sugar, C. A., & Green, M. F. (2022). Social cognition and social problem solving skills training to improve job functioning and tenure in veterans with psychotic disorders. *Psychiatric Rehabilitation Journal*, *45*(3), 291–298. https://doi.org/10.1037/prj0000518

107. Kern, R. S., Zarate, R., Glynn, S. M., Turner, L. R., Smith, K. M., & Mitchell, S. S. (2018). Improving work outcome in supported employment for serious mental illness: Results from 2 independent studies of errorless learning. *Schizophr Bull*, *44*(1), 38–45.

108. Khalifa, N., Talbot, E., Barber, S., Schneider, J., Bird, Y., Attfield, J., Bates, P., Walker, D.-M., & Völlm, B. (2020). A Feasibility Cluster Randomized Controlled Trial of Individual Placement and Support (IPS) for Patients With Offending Histories. *Frontiers in Psychiatry*, *10*, 952. https://doi.org/10.3389/fpsyt.2019.00952

109. Kidd, S. A., Mutschler, C., Lichtenstein, S., Yan, S., Virdee, G., Blair, F., Mihalakakos, G., McKinney, C., Collins, A., Guimond, T., George, T. P., Davidson, L., Velligan, D., & Voineskos, A. (2021). Randomized trial of a brief peer support intervention for individuals with schizophrenia transitioning from hospital to

community. *Schizophrenia Research*, *231*, 214–220. https://doi.org/10.1016/j.schres.2021.03.019

110. Killackey, E., Allott, K., Jackson, H. J., Scutella, R., Tseng, Y.-P., & Borland, J. (2019). Individual placement and support for vocational recovery in first-episode psychosis: Randomised controlled trial. *Br J Psychiatry*, *214*(2), 76–82.

111. Killaspy, H., Priebe, S., & P, M. (2020). Predictors of moving on from mental health supported accommodation in England: National cohort study. *Br J Psychiatry*, *216*, 331–337.

112. Killaspy, H., Priebe, S., & S, B. (2016). Quality of life, autonomy, satisfaction, and costs associated with mental health supported accommodation services in England: A national survey. *Lancet Psychiatry*, *3*, 1129–1137.

113. Kingston, D. A., Olver, M. E., McDonald, J., & Cameron, C. (2018). A randomised controlled trial of a cognitive skills programme for offenders with mental illness. *Crim Behav Ment Health*, *28*(4), 369–382.

114. Kirst, M., Friesdorf, R., Ta, M., Amiri, A., Hwang, S. W., Stergiopoulos, V., & O'Campo, P. (2020). Patterns and effects of social integration on housing stability, mental health and substance use outcomes among participants in a randomized controlled Housing First trial. *Social Science & Medicine*, *265*, 113481. https://doi.org/10.1016/j.socscimed.2020.113481

115. Knight, M. J., Lyrtzis, E., Fourrier, C., Aboustate, N., Sampson, E., Hori, H., Cearns, M., Morgan, J., Toben, C., & Baune, B. T. (2021). Psychological training to improve psychosocial function in patients with major depressive disorder: A randomised clinical trial. *Psychiatry Research*, *300*, 113906. https://doi.org/10.1016/j.psychres.2021.113906

116. Korr, W. S., & Joseph, A. (1995). Housing the homeless mentally ill: Findings from Chicago. *J Soc Serv Res*, *21*(1), 53–68.

117. Kukla, M., Bell, M. D., & Lysaker, P. H. (2018). A randomized controlled trial examining a cognitive behavioral therapy intervention enhanced with cognitive remediation to improve work and neurocognition outcomes among persons with schizophrenia spectrum disorders. *Schizophr Res*, *197*, 400–406.

118. Kumar, R., Nischal, A., & P, D. (2020). Impact of brief psychosocial intervention on key relatives of patients with schizophrenia: A randomized controlled trial. *Indian J Psychiatry*, *62*, 137–144.

119. Lachaud, J., Mejia-Lancheros, C., Nisenbaum, R., Stergiopoulos, V., O'Campo, P., & Hwang, S. W. (2021). Housing First and Severe Mental Disorders: The Challenge of Exiting Homelessness. *The ANNALS of the American Academy of Political and Social Science*, *693*(1), 178–192. https://doi.org/10.1177/0002716220987220

120. Lamberti, J. S., Weisman, R. L., Cerulli, C., Williams, G. C., Jacobowitz, D. B., & Mueser, K. T. (2017). A randomized controlled trial of the Rochester forensic assertive community treatment model. *Psychiatr Serv*, *68*(10), 1016–1024.
121. Latimer, E. A., Rabouin, D., Cao, Z., Ly, A., Powell, G., Aubry, T., Distasio, J., Hwang, S. W., Somers, J. M., Bayoumi, A. M., Mitton, C., Moodie, E. E. M., Goering, P. N., & For the At Home/Chez Soi Investigators. (2020). Cost-Effectiveness of Housing First With Assertive Community Treatment: Results From the Canadian At Home/Chez Soi Trial. *Psychiatric Services*, *71*(10), 1020–1030. https://doi.org/10.1176/appi.ps.202000029
122. Laurila, M., Lindfors, O., Knekt, P., & Heinonen, E. (2024). The effect of individual short- and long-term psychotherapy on perceived social support: Analysis of secondary outcomes of a randomized clinical trial. *Nordic Journal of Psychiatry*, *78*(3), 230–237. https://doi.org/10.1080/08039488.2024.2306229
123. Lecomte, T., Corbière, M., Giguère, C.-E., Titone, D., & Lysaker, P. (2020). Group cognitive behaviour therapy for supported employment–results of a randomized controlled cohort trial. *Schizophr Res*, *215*, 126–133.
124. Lehman, A. F., Dixon, L. B., Kernan, E., DeForge, B. R., & Postrado, L. T. (1997). A randomized trial of assertive community treatment for homeless persons with severe mental illness. *Arch Gen Psychiatry*, *54*(11), 1038–1043.
125. Lemoine, C., Loubière, S., Boucekine, M., Girard, V., Tinland, A., & Auquier, P. (2021). Cost-effectiveness analysis of housing first intervention with an independent housing and team support for homeless people with severe mental illness: A Markov model informed by a randomized controlled trial. *Social Science & Medicine*, *272*, 113692. https://doi.org/10.1016/j.socscimed.2021.113692
126. Lerner, D., Adler, D. A., Rogers, W. H., Ingram, E., & Oslin, D. W. (2020). Effect of Adding a Work-Focused Intervention to Integrated Care for Depression in the Veterans Health Administration: A Randomized Clinical Trial. *JAMA Network Open*, *3*(2), e200075. https://doi.org/10.1001/jamanetworkopen.2020.0075
127. Liang, Y., Li, Y., Lin, G., Cai, C., Yuan, H., & Sheng, Q. (2023). Effectiveness of Group Patient-Led Life Skills Training on Function and Self-Efficacy for People With Schizophrenia: A Quasi-Experimental Study. *Journal of Psychosocial Nursing and Mental Health Services*, *61*(2), 60–67. https://doi.org/10.3928/02793695-20221027-04
128. Lindenmayer, J.-P., McGurk, M., SR, KT, K., A, W., D, H., & L. (2008). A randomized controlled trial of cognitive remediation among inpatients with persistent mental illness. *Psychiatr Serv*, *59*(3), 241–247.
129. Lipton, F., Nutt, S., & Sabatini, A. (1988). Housing the homeless mentally Ill: A longitudinal study of a treatment approach. *Hosp Community Psychiatry*, *39*(1), 40–45.

130. Lloyd-Evans, B., Frerichs, J., Stefanidou, T., Bone, J., Pinfold, V., & Lewis, G. (2020). the community navigator study: Results from a feasibility randomised controlled trial of a programme to reduce loneliness for people with complex anxiety or depression. *PLoS ONE*, *15*(5).

131. Lobban, F., Akers, N., & D, A. (2020). Clinical effectiveness of a web-based peer-supported self-management intervention for relatives of people with psychosis or bipolar (REACT): Online, observer-blind, randomised con-trolled superiority trial. *BMC Psychiatry*, *20*(160).

132. Loubière, S., Lemoine, C., Boucekine, M., Boyer, L., Girard, V., Tinland, A., Auquier, P., & for the French Housing First Study Group. (2022). Housing First for homeless people with severe mental illness: Extended 4-year follow-up and analysis of recovery and housing stability from the randomized *Un Chez Soi d'Abord* trial. *Epidemiology and Psychiatric Sciences*, *31*, e14. https://doi.org/10.1017/S2045796022000026

133. Lysaker, P. H., Bond, G., Davis, L. W., Bryson, G. J., & Bell, M. D. (2005). Enhanced cognitive-behavioral therapy for vocational rehabilitation in schizophrenia: Effects on hope and work. *J Rehabil Res Dev*, *42*(5).

134. Lystad, J., Falkum, E., & V, H. (2017). Cognitive remediation and occupational outcome in schizophrenia spectrum disorders: A 2 year follow-up study. *Schizophr Res*, *185*, 122–129.

135. Macnaughton, E., Nelson, G., & SK, W. (2018). Navigating complex implemen-tation contexts: Overcoming barriers and achieving outcomes in a national initiative to scale out Housing First in Canada. *Am J Community Psychol*, *62*, 135–149.

136. Mahlke, C., Priebe, S., & K, H. (2017). Effectiveness of one-to-one peer sup-port for patients with severe mental illness – a randomised controlled trial. *Eur Psychiatry*, *42*, 103–110.

137. Marder, W., SR, WC, M., J, M., & J. (1996). Two-year outcome of social skills training and group psychotherapy for outpatients with schizophrenia. *Am J Psychiatry*, *153*(12).

138. Martin-Carrasco, M., Fernandez-Catalina, P., & A, D.-P. (2016). A randomized trial to assess the efficacy of a psychoeducational intervention on caregiver burden in schizophrenia. *Eur Psychiatry*, *33*, 9–17.

139. Martini, A., Rettore, E., Barbetta, G. P., & Sandrolini, F. (2022). When Non-Compliance Carries the Day: Evaluating the Effectiveness of an Employment Program for the Severely Mentally Ill. *Evaluation Review*, *46*(5), 555–577. https://doi.org/10.1177/0193841X211049685

140. Maru, M., Rogers, E. S., Nicolellis, D., Legere, L., Placencio-Castro, M., Magee, C., & Harbaugh, A. G. (2021). Vocational peer support for adults with psychiatric disabilities: Results of a randomized trial. *Psychiatric Rehabilitation Journal*, *44*(4), 327–336. https://doi.org/10.1037/prj0000484

141. McGurk, M., SR, KT, F., K, W., R, P., & A. (2007). Cognitive training for supported employment: 2–3 year outcomes of a randomized controlled trial. *Am J Psychiatry*, *164*(3), 437–441.
142. McGurk, M., SR, KT, X., H, F., K, S., Y, K., & L. (2016). Cognitive remediation for vocational rehabilitation nonresponders. *Schizophr Res*, *175*(1–3), 48–56.
143. McGurk, M., SR, KT, X., H, W., J, K., S, D., & R.E. (2015). Cognitive enhancement treatment for people with mental illness who do not respond to supported employment: A randomized controlled trial. *Am J Psychiatry*, *172*(9), 852–861.
144. McGurk, S., Mueser, K., & M, W. (2017). The feasibility of implementing cog-nitive remediation for work in community based psychiatric rehabilitation programs. *Psychiatr Rehabil J*, *40*(79).
145. McHugo, G. J., Bebout, R. R., Harris, M., Cleghorn, S., Herring, G., & Xie, H. (2004). A randomized controlled trial of integrated versus parallel housing services for homeless adults with severe mental illness. *Schizophr Bull*, *30*(4), 969–982.
146. Mejia-Lancheros, C., Lachaud, J., Stergiopoulos, V., Matheson, F. I., Nisenbaum, R., O'Campo, P., & Hwang, S. W. (2020). Effect of Housing First on violence-related traumatic brain injury in adults with experiences of homelessness and mental illness: Findings from the At Home/Chez Soi randomised trial, Toronto site. *BMJ Open*, *10*(12), e038443. https://doi.org/10.1136/bmjopen-2020-038443
147. Mervis, J. E., Fiszdon, J. M., Lysaker, P. H., Nienow, T. M., Mathews, L., & Wardwell, P. (2017). Effects of the Indianapolis Vocational Intervention Program (IVIP) on defeatist beliefs, work motivation, and work outcomes in serious mental illness. *Schizophr Res*, *182*, 129–134.
148. Metts, A. V., LeBeau, R. T., Craske, M. G., & Himle, J. A. (2023). Perceived interpersonal competence as a predictor of clinical outcomes in a randomized controlled trial for social anxiety and employment. *Cognitive Behaviour Therapy*, *52*(2), 146–162. https://doi.org/10.1080/16506073.2022.2137578
149. Milligan-Saville, J., Tan, L., Gayed, A., Barnes, C., Madan, I., Dobson, M., Bryant, R., Christensen, A., & Harvey, S. (2017). Workplace Mental Health training for managers and its effects on sick leave: A cluster randomised controlled trial. *The Lancet Psychiatry*, *4*(11), 850–858.
150. Minor, K. S., Marggraf, M. P., Davis, B. J., Mickens, J. L., Abel, D. B., Robbins, M. L., Buck, K. D., Wiehe, S. E., & Lysaker, P. H. (2022). Personalizing interventions using real-world interactions: Improving symptoms and social functioning in schizophrenia with tailored metacognitive therapy. *Journal of Consulting and Clinical Psychology*, *90*(1), 18–28. https://doi.org/10.1037/ccp0000672

151. Mirsepassi, Z., Tabatabaee, M., & V, S. (2018). Patient and family psychoeduca-tion: Service development and implementation in a center in Iran. *Int J Soc Psychiatry*, *64*, 73–79.

152. Monson, C. M., Wagner, A. C., Crenshaw, A. O., Whitfield, K. M., Newnham, C. M., Valela, R., Varma, S., Di Bartolomeo, A. A., Fulham, L., Collins, A., Donkin, V., Mensah, D. H., Landy, M. S. H., Samonas, C., Morland, L., Doss, B. D., & Fitzpatrick, S. (2022). An uncontrolled trial of couple HOPES: A guided online couple intervention for PTSD and relationship enhancement. *Journal of Family Psychology*, *36*(6), 1036–1042. https://doi.org/10.1037/fam0000976

153. Morse, G. A., Calsyn, R. J., Allen, G., Tempethoff, B., & Smith, R. (1992). Experimental comparison of the effects of three treatment programs for homeless mentally ill people. *Psychiatr Serv*, *43*(10), 1005–1010.

154. Morse, G. A., Calsyn, R. J., Klinkenberg, W. D., Helminiak, T. W., Wolff, N., & Drake, R. E. (2006). Treating homeless clients with severe mental illness and substance use disorders: Costs and outcomes. *Community Ment Health J*, *42*(4), 377–404.

155. Morse, G. A., Calsyn, R., Klinkenberg, W., Trusty, M., Gerber, F., & Smith, R. (1997). An experimental comparison of three types of case management for homeless mentally Ill persons. *Psychiatr Serv*, *48*(4), 497–503.

156. Mötteli, S., Adamus, C., Deb, T., Fröbel, R., Siemerkus, J., Richter, D., & Jäger, M. (2022). Independent Supported Housing for Non-homeless People With Serious Mental Illness: A Pragmatic Randomized Controlled Trial. *Frontiers in Psychiatry*, *12*, 798275.https://doi.org/10.3389/fpsyt.2021.798275

157. Moxham, L., Taylor, E., & C, P. (2017). Goal setting among people living with mental illness: A qualitative analysis of recovery camp. *Issues Ment Health Nurs*, *38*, 420–424.

158. Mueller, N. E., & Cougle, J. R. (2023). Building Closer Friendships in social anxiety disorder: A randomized control trial of an internet-based intervention. *Journal of Behavior Therapy and Experimental Psychiatry*, *78*, 101799. https://doi.org/10.1016/j.jbtep.2022.101799

159. Mueser, K. T., Aalto, S., Becker, D. R., Ogden, J. S., Wolfe, R. S., & Schiavo, D. (2005). The effectiveness of skills training for improving outcomes in supported employment. *Psychiatr Serv*,*56*(10), 1254–1260.

160. Mustafa, S., Malla, A., Joober, R., Abadi, S, Latimer, E., Schmitz, N., Jarvis, G., Margolese, H., Cascalenda, N., Abdel-Baki, A., & Iyer, S. (2022). Unfinished business: Functional outcomes in a randomized controlled trial of a three-year extension of early intervention versus regular care following two years of early intervention for psychosis. *Acta Psychiatrica Scandinavica*, *145*(1), 86–99. https://dx.doi.org/10.1111/acps.13377

161. Nezafat Ferizi, J., Ashouri, A., Gharraee, B., & Asgharnejad Farid, A. A. (2023). Comparison of the Effectiveness of Interpersonal Counseling and

Interpersonal Psychotherapy in Emotional Expression, Social Skills, and Depression Symptoms in Students. *Iranian Journal of Psychiatry and Behavioral Sciences*, *17*(2). https://doi.org/10.5812/ijpbs-130443

162. Nguyen, T., Tran, T., & S, G. (2020). Proof of concept of participant informed, psycho-educational, community-based intervention for people with severe mental illness in rural Vietnam. *Int J Soc Psychiatry*, *66*, 232–239.

163. Niedermoser, D. W., Kalak, N., Kiyhankhadiv, A., Brand, S., Walter, C., Schweinfurth, N., & Lang, U. E. (2020). Workplace-Related Interpersonal Group Psychotherapy to Improve Life at Work in Individuals With Major Depressive Disorders: A Randomized Interventional Pilot Study. *Frontiers in Psychiatry*, *11*, 168. https://doi.org/10.3389/fpsyt.2020.00168

164. Nijman, S. A., Pijnenborg, G. H. M., Vermeer, R. R., Zandee, C. E. R., Zandstra, D. C., Van Der Vorm, D., De Wit - De Visser, A. C., Meins, I. A., Geraets, C. N. W., & Veling, W. (2023). Dynamic Interactive Social Cognition Training in Virtual Reality (DiSCoVR) versus Virtual Reality Relaxation (VRelax) for People With a Psychotic Disorder: A Single-Blind Multicenter Randomized Controlled Trial. *Schizophrenia Bulletin*, *49*(2), 518–530. https://doi.org/10.1093/schbul/sbac166

165. Noordik, E., Klink, J. J., Geskus, R. B., Boer, M. R., Dijk, F. J., & Nieuwenhuijsen, K. (2013). Effectiveness of an exposure-based return-to-work program for workers on sick leave due to common mental disorders: A cluster-randomized controlled trial. *Scand J Work Environ Health*, *39*, 144–154.

166. Nuechterlein, K. H., Subotnik, K. L., Ventura, J., Turner, L. R., Gitlin, M. J., Gretchen-Doorly, D., Becker, D. R., Drake, R. E., Wallace, C. J., & Liberman, R. P. (2019). Enhancing return to work or school after a first episode of schizophrenia: The UCLA RCT of Individual Placement and Support and Workplace Fundamentals Module training. *Psychol Med*, *50*(1), 20–28.

167. O’Campo, P., Nisenbaum, R., Crocker, A. G., Nicholls, T., Eiboff, F., & Adair, C. E. (2023). Women experiencing homelessness and mental illness in a Housing First multi-site trial: Looking beyond housing to social outcomes and well-being. *PLOS ONE*, *18*(2), e0277074. https://doi.org/10.1371/journal.pone.0277074

168. O’Connell, M., Sledge, W., & M, S. (2018). Outcomes of a peer mentor inter-vention for persons with recurrent psychiatric hospitalization. *Psychiatr Serv*, *69*, 760–767.

169. O’Connell, M., Tsai, J., & Rosenheck, R. (2023). Beyond Supported Housing: Correlates of Improvements in Quality of Life Among Homeless Adults with Mental Illness. *Psychiatric Quarterly*, *94*(1), 49–59. https://doi.org/10.1007/s11126-022-10010-x

170. Ohki, Y., Igarashi, Y., & Yamauchi, K. (2021). Re-work Program in Japan—Overview and Outcome of the Program. *Frontiers in Psychiatry*, *11*, 616223. https://doi.org/10.3389/fpsyt.2020.616223

171. Okpaku, S. O., Anderson, K. H., Sibulkin, A. E., Butler, J., & Bickman, L. (1997). The effectiveness of a multidisciplinary case management intervention on the employment of SSDI applicants and beneficiaries. *Psychiatr Rehabil J*, *20*(3).

172. Øverland, S., Grasdal, A. L., & Reme, S. E. (2018). Long-term effects on income and sickness benefits after work-focused cognitive-behavioural therapy and individual job support: A pragmatic, multicentre, randomised controlled trial. *Occup Environ Med*, *75*(10), 703–708.

173. Oxford, M. L., Hash, J. B., Lohr, M. J., Bleil, M. E., Fleming, C. B., Unützer, J., & Spieker, S. J. (2021). Randomized trial of promoting first relationships for new mothers who received community mental health services in pregnancy. *Developmental Psychology*, *57*(8), 1228–1241. https://doi.org/10.1037/dev0001219

174. Padmakar, A., Wit, E., & S, M. (2020). Supported Housing as a recovery option for long-stay patients with severe mental illness in a psychiatric hospital in South India: Learning from an innovative de-hospitalization process. *PLoS One*, *15:e0230074*.

175. Pérez-Corrales, J., Pérez-de-Heredia-Torres, M., & R, M.-P. (2019). Be-ing normal' and self-identity: The experience of volunteering in individuals with severe mental disorders – a qualitative study. *BMJ Open*, *9:e025363*.

176. Perkins, R., Spiro, N., & Waddell, G. (2023). Online songwriting reduces loneliness and postnatal depression and enhances social connectedness in women with young babies: Randomised controlled trial. *Public Health*, *220*, 72–79. https://doi.org/10.1016/j.puhe.2023.04.017

177. Perlick, D., Jackson, C., & S, G. (2018). Randomized trial comparing caregiver-only family-focused treatment to standard health education on the 6-month outcome of bipolar disorder. *Bipolar Disord*, *20*, 622–633.

178. Pichler, E.-M., Stulz, N., Wyder, L., Heim, S., Watzke, B., & Kawohl, W. (2021). Long-Term Effects of the Individual Placement and Support Intervention on Employment Status: 6-Year Follow-Up of a Randomized Controlled Trial. *Frontiers in Psychiatry*, *12*, 709732.https://doi.org/10.3389/fpsyt.2021.709732

179. Pos, K., Franke, N., Smit, F., Wijnen, B. F., Staring, A. B., & Gaag, M. (2019). Cognitive behavioral therapy for social activation in recent-onset psychosis: Randomized controlled trial. *J Consult Clin Psychol*, *87*(2).

180. Pot-Kolder, R., Geraets, C., Veling, W., Beilen, M., Staring, A., & Gijsman, H. (2018). Virtual-reality-based cognitive behavioural therapy versus waiting list control for paranoid ideation and social avoidance in patients with psychotic disorders: A single-blind randomised controlled trial. *Lancet Psychiatry*, *5*(3).

181. Priebe, S., Chevalier, A., Hamborg, T., Golden, E., King, M., & Pistrang, N. (2020). Effectiveness of a volunteer befriending programme for patients with schizophrenia: Randomised controlled trial. *Br J Psychiatry*, *217*(3), 477–483.

182. Prince, J., Mora, O., & J, A. (2018). Nine ways that clubhouses foster interper-sonal connection for persons with severe mental illness: Lessons for other types of programs. *Soc Work Ment Health*, *16*, 321–336.

183. Puig, O., Thomas, K., & Twamley, E. (2016). Age and improved attention predict work attainment in combined compensatory cognitive training and supported employment for people with severe mental illness. *J Nerv Ment Dis*, *204*, 869–872.

184. Rajji, T. K., Mamo, D. C., Holden, J., Granholm, E., & Mulsant, B. H. (2022). Cognitive-Behavioral Social Skills Training for patients with late-life schizophrenia and the moderating effect of executive dysfunction. *Schizophrenia Research*, *239*, 160–167. https://doi.org/10.1016/j.schres.2021.11.051

185. Raven, M. C., Niedzwiecki, M. J., & Kushel, M. (2020). A randomized trial of permanent supportive housing for chronically homeless persons with high use of publicly funded services. *Health Services Research*, *55*(S2), 797–806. https://doi.org/10.1111/1475-6773.13553

186. Rebergen, D. S., Bruinvels, D. J., Bezemer, P. D., Beek, A. J., & Mechelen, W. (2009). Guideline-based care of common mental disorders by occupational physicians (CO-OP study): A randomized controlled trial. *J Occup Environ Med*, *51*(3), 305–312.

187. Reme, S. E., Monstad, K., Fyhn, T., Sveinsdottir, V., Løvvik, C., & Lie, S. A. (2019). A randomized controlled multicenter trial of individual placement and support for patients with moderate-to-severe mental illness. *Scand J Work Environ Health*, *45*(1).

188. Rhenter, P., Moreau, D., & C, L. (2018). Bread and shoulders: Reversing the down-ward spiral, a qualitatitve analyses of the effects of a Housing First-type pro-gram in France. *Int J Env Res Pub Health*, *15*(520).

189. Rivera, J. J., Sullivan, A. M., & Valenti, S. S. (2007). Adding consumer-providers to intensive case management: Does it improve outcome? *Psychiatr Serv*, *58*(6), 802–809.

190. Roberts, D. L., Combs, D. R., Willoughby, M., Mintz, J., Gibson, C., & Rupp, B. (2014). A randomized, controlled trial of Social Cognition and Interaction Training (SCIT) for outpatients with schizophrenia spectrum disorders. *Br J Clin Psychol*, *53*(3), 281–298.

191. Rodríguez Pulido, F., Caballero Estebaranz, N., Gonzalez Davilla, E., & Melian Cartaya, M. J. (2019). Cognitive remediation to improve the vocational outcomes of people with severe mental illness. *Neuropsychol Rehabil*, *31*(2), 1–23.

192. Rogers, E. S., Anthony, W. A., Lyass, A., & Penk, W. E. (2006). A randomized clinical trial of vocational rehabilitation for people with psychiatric disabilities. *Rehabil Couns Bull*, *49*(3), 143–156.

193. Roos, E., Bjerkeset, O., & E, S. (2016). A qualitative study of how people with severe mental illness experience living in sheltered housing with a private fully equipped apartment. *BMC Psychiatry*, *16*(186).

194. Rössler, W., Kawohl, W., Nordt, C., Haker, H., Rüsch, N., & Hengartner, M. P. (2020). Placement budgets' for supported employment: Impact on employment rates in a multicentre randomised controlled trial. *Br J Psychiatry*, *216*(6), 308–313.

195. Rouse, J., Mutschler, C., & K, M. (2017). Qualitative participatory evaluation of a psychosocial rehabilitation program for individuals with severe mental illness. *Int J Ment Health*, *46*, 139–156.

196. Rowe, M., Bellamy, C., Baranoski, M., Wieland, M., O'Connell, M. J., & Benedict, P. (2007). A peer-support, group intervention to reduce substance use and criminality among persons with severe mental illness. *Psychiatr Serv*, *58*(7), 955–961.

197. Ruiz-Comellas, A., Valmaña, G. S., Catalina, Q. M., Baena, I. G., Mendioroz Peña, J., Roura Poch, P., Sabata Carrera, A., Cornet Pujol, I., Casaldàliga Solà, À., Fusté Gamisans, M., Saldaña Vila, C., Vázquez Abanades, L., & Vidal-Alaball, J. (2022). Effects of Physical Activity Interventions in the Elderly with Anxiety, Depression, and Low Social Support: A Clinical Multicentre Randomised Trial. *Healthcare*, *10*(11), 2203. https://doi.org/10.3390/healthcare10112203

198. Russinova, Z., Gidugu, V., Bloch, P., Restrepo-Toro, M., & Rogers, E. S. (2018). Empowering individuals with psychiatric disabilities to work: Results of a randomized trial. *Psychiatr Rehabil J*, *41*(3).

199. Russinova, Z., Gidugu, V., Rogers, E. S., Legere, L., & Bloch, P. (2023). Fostering the community participation of individuals with psychiatric disabilities: Effectiveness of a new peer-led photovoice-based intervention. *Psychiatric Rehabilitation Journal*, *46*(3), 196–210. https://doi.org/10.1037/prj0000540

200. Saavedra, J., Arias, S., & P, C. (2018). Impact of creative workshops for people with severe mental health problems: Art as a means of recovery. *Arts Health*, *10*, 241–256.

201. Sacks, S., Chaple, M., Sacks, J. Y., McKendrick, K., & Cleland, C. M. (2012). Randomized trial of a reentry modified therapeutic community for offenders with co-occurring disorders: Crime outcomes. *J Subst Abuse Treat*, *42*(3), 247–259.

202. Sacks, S., Sacks, J. Y., McKendrick, K., Banks, S., & Stommel, J. (2004). Modified TC for MICA offenders: Crime outcomes. *Behav Sci Law*, *22*(4), 477–501.

203. Salomonsson, S., Santoft, F., Lindsäter, E., Ejeby, K., Ingvar, M., Ljótsson, B., Öst, L., Lekander, M., & Hedman-Lagerlöf, E. (2020). Effects of cognitive behavioural therapy and return-to-work intervention for patients on sick leave due to stress-related disorders: Results from a randomized trial. *Scandinavian Journal of Psychology*, *61*(2), 281–289. https://doi.org/10.1111/sjop.12590

204. Salzer, M. S., Rogers, J., Salandra, N., O'Callaghan, C., Fulton, F., & Balletta, A. A. (2016). Effectiveness of peer-delivered Center for Independent Living supports for individuals with psychiatric disabilities: A randomized, controlled trial. *Psychiatr Rehabil J*, *39*(3).

205. Sanches, S. A., Feenstra, T. L., Swildens, W. E., Van Busschbach, J. T., Van Weeghel, J., & Van Asselt, T. D. I. (2022). Cost Effectiveness and Budget Impact of the Boston University Approach to Psychiatric Rehabilitation for Increasing the Social Participation of Individuals With Severe Mental Illnesses. *Frontiers in Psychiatry*, *13*, 880482. https://doi.org/10.3389/fpsyt.2022.880482

206. Sanches, S. A., Swildens, W. E., Schaefer, B., Moerbeek, M., Feenstra, T. L., & Asselt, A. D. (2020). Effectiveness of the Boston University approach to psychiatric rehabilitation in improving social participation in people with severe mental illnesses: A randomized controlled trial. *Front Psych*, *11*(970).

207. Scanlan, J., Feder, K., & P, E. (2019). Outcomes of an individual placement and support programme incorporating principles of the collaborative recovery model. *Aust Occup Ther J*, *66*, 519–529.

208. Schene, A. H., Koeter, M. W., Kikkert, M. J., Swinkels, J. A., & McCrone, P. (2007). Adjuvant occupational therapy for work-related major depression works: Randomized trial including economic evaluation. *Psychol Med*, *37*(3).

209. Schneider, J., Akhtar, A., & N, B. (2016). Individual placement and support ver-sus individual placement and support enhanced with work-focused cogni-tive behaviour therapy: Feasibility study for a randomised controlled trial. *Br J Occup Ther*, *79*, 257–269.

210. Schramm, E., Mack, S., Thiel, N., Jenkner, C., Elsaesser, M., & Fangmeier, T. (2020). Interpersonal Psychotherapy vs. Treatment as Usual for Major Depression Related to Work Stress: A Pilot Randomized Controlled Study. *Frontiers in Psychiatry*, *11*, 193. https://doi.org/10.3389/fpsyt.2020.00193

211. Segal, S. P., Silverman, C. J., & Temkin, T. L. (2010). Self-help and community mental health agency outcomes: A recovery-focused randomized controlled trial. *Psychiatr Serv*, *61*(9), 905–910.

212. Seoane-Bouzas, M., De-Rosende-Celeiro, I., & Meijide-Failde, R. (2022). A pilot randomized controlled trial of aquatic-based activities in a group occupational therapy program for adults living with serious mental illness in Spain. *Health & Social Care in the Community*, *30*(4). https://doi.org/10.1111/hsc.13544

213. Shen, Z.-H., Liu, M.-H., Wu, Y., Lin, Q.-Q., & Wang, Y.-G. (2022). Virtual-reality-based social cognition and interaction training for patients with schizophrenia: A preliminary efficacy study. *Frontiers in Psychiatry*, *13*, 1022278. https://doi.org/10.3389/fpsyt.2022.1022278

214. Sheridan, A. J., Drennan, J., Coughlan, B., O'Keeffe, D., Frazer, K., & Kemple, M. (2015). Improving social functioning and reducing social isolation and loneliness among people with enduring mental illness: Report of a randomised controlled trial of supported socialisation. *Int J Soc Psychiatry*, *61*(3), 241–250.

215. Shern, D. L., Tsemberis, S., Anthony, W., Lovell, A. M., Richmond, L., & Felton, C. J. (2000). Serving street-dwelling individuals with psychiatric disabilities: Outcomes of a psychiatric rehabilitation clinical trial. *Am J Public Health*, *90*(12).

216. Shih, C.-A., & Yang, M.-H. (2023). Effect of Animal-Assisted Therapy (AAT) on Social Interaction and Quality of Life in Patients with Schizophrenia during the COVID-19 Pandemic: An Experimental Study. *Asian Nursing Research*, *17*(1), 37–43. https://doi.org/10.1016/j.anr.2023.01.002

217. Shimada, T., Inagaki, Y., Shimooka, Y., Kawano, K., Tanaka, S., & Kobayashi, M. (2022). Effect of individualized occupational therapy on social functioning in patients with schizophrenia: A five-year follow-up of a randomized controlled trial. *Journal of Psychiatric Research*, *156*, 476–484. https://doi.org/10.1016/j.jpsychires.2022.10.066

218. Sikira, H., Janković, S., Slatina, M. S., Muhić, M., Sajun, S., Priebe, S., & Džubur Kulenović, A. (2021). The effectiveness of volunteer befriending for improving the quality of life of patients with schizophrenia in Bosnia and Herzegovina – an exploratory randomised controlled trial. *Epidemiology and Psychiatric Sciences*, *30*, e48. https://doi.org/10.1017/S2045796021000330

219. Silverman, M. J. (2014). Effects of a live educational music therapy intervention on acute psychiatric inpatients' perceived social support and trust in the therapist: A four-group randomized effectiveness study. *J Music Ther*, *51*(3), 228–249.

220. Smidl, S., Mitchell, D., & Creighton, C. (2017). Outcomes of a therapeutic gardening pro-gram in a mental health recovery center. *Occup Ther Mental Health*, *33*, 374–385.

221. Smith, M. J., Smith, J. D., Blajeski, S., Ross, B., Jordan, N., Bell, M. D., McGurk, S. R., Mueser, K. T., Burke-Miller, J. K., Oulvey, E. A., Fleming, M. F., Nelson, K., Brown, A., Prestipino, J., Pashka, N. J., & Razzano, L. A. (2022). An RCT of Virtual Reality Job Interview Training for Individuals With Serious Mental Illness in IPS Supported Employment. *Psychiatric Services*, *73*(9), 1027–1038. https://doi.org/10.1176/appi.ps.202100516

222. Snethen, G., McCormick, B. P., Nagata, S., & Salzer, M. S. (2024). Independence through community access and navigation: A supported leisure intervention for individuals with negative symptoms. *Psychiatric Rehabilitation Journal*. https://doi.org/10.1037/prj0000593

223. Solar, A., Bennett, K., & Hulse, G. (2023). An intervention at an adult inpatient unit to engage people with schizophrenia and work goals with supported employment. *Australasian Psychiatry*, *31*(1), 82–89. https://doi.org/10.1177/10398562221147429

224. Somers, J., Moniruzzaman, A., & M, P. (2017). A randomized trial examining Housing First in congregate and scattered site formats. *PLoS One*, *12:e0168745*.

225. Sommer, J., Gill, K., & J, S.-P. (2019). The role of recovery colleges in supporting personal goal achievement. *Psychiatr Rehabil J*, *42*, 394–400.

226. Sroosh, S., ZareBahramabadi, M., & Nasrollahi, B. (2023). Comparing the Effect of Integrative-Behavioral and Emotion-Focused Couple Therapies on Marital Adjustment of Couples with Obsessive-Compulsive Disorder. *International Journal of Behavioral Sciences*, *16*(4). https://doi.org/10.30491/ijbs.2023.362831.1855

227. Stanhope, V., Choy-Brown, M., & E, T. (2016). Case manager perspectives on the role of treatment in supportive housing for people with severe mental illness. *J Soc Soc Work Res*, *7*, 507–525.

228. Stergiopoulos, V., Gozdzik, A., & V, M. (2016). The effectiveness of a Housing First adaptation for ethnic minority groups: Findings of a pragmatic randomized controlled trial. *BMC Public Health*, *16*(1).

229. Stergiopoulos, V., Hwang, S. W., Gozdzik, A., Nisenbaum, R., Latimer, E., & Rabouin, D. (2015). Effect of scattered-site housing using rent supplements and intensive case management on housing stability among homeless adults with mental illness: A randomized trial. *JAMA*, *313*(9), 905–915.

230. Stergiopoulos, V., Zerger, S., & J, J. (2016). Dynamic sustainability: Practitioners' perspectives on Housing First implementation challenges and model fidelity over time. *Res Soc Work Pract*, *26*, 61–68.

231. Stroupe, K. T., Jordan, N., Richman, J., Bond, G. R., Pogoda, T. K., Cao, L., Kertesz, S. G., Kyriakides, T. C., & Davis, L. L. (2022). Cost-Effectiveness of Individual Placement and Support Compared to Transitional Work Program for Veterans with Post-traumatic Stress Disorder. *Administration and Policy in Mental Health and Mental Health Services Research*, *49*(3), 429–439. https://doi.org/10.1007/s10488-021-01173-z

232. Susser, E., Valencia, E., Conover, S., Felix, A., Tsai, W.-Y., & Wyatt, R. J. (1997). Preventing recurrent homelessness among mentally ill men: A" critical time" intervention after discharge from a shelter. *Am J Public Health*, *87*(2), 256–262.

233. Sutton, R., Lawrence, K., & E, Z. (2019). Recovery college influences upon service users: A recovery academy exploration of employment and service use. *J Mental Health Train Educ Pract*, *14*, 141–148.

234. Swinkels, L. T. A., Van Der Pol, T. M., Twisk, J., Ter Harmsel, J. F., Dekker, J. J. M., & Popma, A. (2023). The effectiveness of an additive informal social network intervention for forensic psychiatric outpatients: Results of a randomized controlled trial. *Frontiers in Psychiatry*, *14*, 1129492. https://doi.org/10.3389/fpsyt.2023.1129492

235. Talbot, E., Bird, Y., & J, R. (2018). Implementation of individual placement and support (IPS) into community forensic mental health settings: Lessons learned. *Br J Occup Ther*, *81*, 338–347.

236. Taylor, C. T., Pearlstein, S. L., Kakaria, S., Lyubomirsky, S., & Stein, M. B. (2020). Enhancing Social Connectedness in Anxiety and Depression Through Amplification of Positivity: Preliminary Treatment Outcomes and Process of Change. *Cognitive Therapy and Research*, *44*(4), 788–800. https://doi.org/10.1007/s10608-020-10102-7

237. Terzian, E., Tognoni, G., Bracco, R., Ruggieri, E., Ficociello, R. A., & Mezzina, R. (2013). Social network intervention in patients with schizophrenia and marked social withdrawal: A randomized controlled study. *Can J Psychiatry*, *58*(11), 622–631.

238. Thomas, E., & Salzer, M. (2018). Associations between the peer support relationship, service satisfaction and recovery-oriented outcomes: A correlational study. *J Mental Health*, *27*, 352–358.

239. Thompson, A., Elahi, F., Realpe, A., Birchwood, M., Taylor, D., Vlaev, I., Leahy, F., & Bucci, S. (2020). A Feasibility and Acceptability Trial of Social Cognitive Therapy in Early Psychosis Delivered Through a Virtual World: The VEEP Study. *Frontiers in Psychiatry*, *11*, 219. https://doi.org/10.3389/fpsyt.2020.00219

240. Tinland, A., Loubiere, S., Boucekine, M., Boyer, L., Fond, G., & Girard, V. (2020). Effectiveness of a housing support team intervention with a recovery-oriented approach on hospital and emergency department use by homeless people with severe mental illness: A randomised controlled trial. *Epidemiol Psychiatr Sci*, *29:e169*.

241. Tjaden, C., Mulder, C. L., Den Hollander, W., Castelein, S., Delespaul, P., Keet, R., Van Weeghel, J., & Kroon, H. (2021). Effectiveness of Resource Groups for Improving Empowerment, Quality of Life, and Functioning of People With Severe Mental Illness: A Randomized Clinical Trial. *JAMA Psychiatry*, *78*(12), 1309. https://doi.org/10.1001/jamapsychiatry.2021.2880

242. Tse, S., Ng, C. S. M., Yuen, W. W. Y., Lo, I. W. K., Fukui, S., Goscha, R. J., Wan, E., Wong, S., & Chan, S.-K. (2021). Process research: Compare and contrast the recovery-orientated strengths model of case management and

usual community mental health care. *BMC Psychiatry*,*21*(1), 534. https://doi.org/10.1186/s12888-021-03523-5

243. Tsemberis, S., Gulcur, L., & Nakae, M. (2004). Housing first, consumer choice, and harm reduction for homeless individuals with a dual diagnosis. *Am J Public Health*, *94*(4), 651–656.

244. Twamley, E. W., Thomas, K. R., Burton, C. Z., Vella, L., Jeste, D. V., & Heaton, R. K. (2019). Compensatory cognitive training for people with severe mental illnesses in supported employment: A randomized controlled trial. *Schizophr Res*, *203*, 41–48.

245. Valentine, L., McEnery, C., O'Sullivan, S., Gleeson, J., Bendall, S., & Alvarez-Jimenez, M. (2020). Young People's Experience of a Long-Term Social Media–Based Intervention for First-Episode Psychosis: Qualitative Analysis. *Journal of Medical Internet Research*, *22*(6), e17570. https://doi.org/10.2196/17570

246. Valls, È., Bonnín, C. M., Torres, I., Brat, M., Prime-Tous, M., Morilla, I., Segú, X., Solé, B., Torrent, C., Vieta, E., Martínez-Arán, A., Reinares, M., & Sánchez-Moreno, J. (2022). Efficacy of an integrative approach for bipolar disorder: Preliminary results from a randomized controlled trial. *Psychological Medicine*, *52*(16), 4094–4105. https://doi.org/10.1017/S0033291721001057

247. van Beurden, K. M., Brouwers, E. P., Joosen, M. C., de Boer, M. R., van Weeghel, J. T. B., & van der Klink, J. J. (2017). Effectiveness of an intervention to enhance occupational physicians' guideline adherence on sickness absence duration in workers with common mental disorders: A cluster-randomized controlled trial. *Journal of Occupational Rehabilitation*, *27*(4), 559–567.

248. Van Der Stouwe, E. C. D., De Vries, B., Steenhuis, L. A., Waarheid, C. O., Jans, R., De Jong, S., Aleman, A., Pijnenborg, G. H. M., & Van Busschbach, J. T. (2022). BEATVIC, a body-oriented resilience therapy for individuals with psychosis: Short term results of a multi-center RCT. *PLOS ONE*, *17*(12), e0279185. https://doi.org/10.1371/journal.pone.0279185

249. Van Lieshout, R. J., Layton, H., Savoy, C. D., Brown, J. S. L., Ferro, M. A., Streiner, D. L., Bieling, P. J., Feller, A., & Hanna, S. (2021). Effect of Online 1-Day Cognitive Behavioral Therapy–Based Workshops Plus Usual Care vs Usual Care Alone for Postpartum Depression: A Randomized Clinical Trial. *JAMA Psychiatry*, *78*(11), 1200. https://doi.org/10.1001/jamapsychiatry.2021.2488

250. Van Veen, M., Koekkoek, B., Teerenstra, S., Adang, E., & Mulder, C. L. (2021). Effectiveness and cost effectiveness of interpersonal community psychiatric treatment (ICPT) for people with long-term severe non-psychotic mental disorders: A multi-Centre randomized controlled trial. *BMC Psychiatry*, *21*(1), 261. https://doi.org/10.1186/s12888-021-03264-5

251. Varga, E., Endre, S., & T, B. (2018). Community-based psychosocial treatment has an impact on social processing and functional outcome in schizophre-nia. *Front Psychiatry*, *9*(247).

252. Vauth, R., Corrigan, P. W., Clauss, M., Dietl, M., Dreher-Rudolph, M., & Stieglitz, R.-D. (2005). Cognitive strategies versus self-management skills as adjunct to vocational rehabilitation. *Schizophr Bull*, *31*(1), 55–66.

253. Vlasveld, M. C., Feltz-Cornelis, C. M., Adèr, H. J., Anema, H., JR, R, van M., & W. (2013). Collaborative care for sick-listed workers with major depressive disorder: A randomised controlled trial from the Netherlands depression Initiative aimed at return to work and depressive symptoms. *Occup Environ Med*, *70*(4), 223–230.

254. Vogel, J. S., Bruins, J., Swart, M., Liemburg, E., Van Der Gaag, M., & Castelein, S. (2023). Effects of an eating club for people with a psychotic disorder on personal recovery: Results of a randomized controlled trial. *Journal of Behavior Therapy and Experimental Psychiatry*, *81*, 101871. https://doi.org/10.1016/j.jbtep.2023.101871

255. Volker, D., Zijlstra-Vlasveld, M. C., Anema, B., JR, AT, B., EP, E., & W.H. (2015). Effectiveness of a blended web-based intervention on return to work for sick-listed employees with common mental disorders: Results of a cluster randomized controlled trial. *J Med Internet Res*, *17*(5).

256. Webber, M., Ngamaba, K., Moran, N., Pinfold, V., Boehnke, J. R., Knapp, M., Henderson, C., Rehill, A., & Morris, D. (2021). The Implementation of Connecting People in Community Mental Health Teams in England: A Quasi-Experimental Study. *The British Journal of Social Work*, *51*(3), 1080–1100. https://doi.org/10.1093/bjsw/bcaa159

257. Whitley, R., Sitter, K., & G, A. (2021). A meaningful focus: Investigating the impact of involvement in a participatory video program on the recovery of participants with severe mental illness. *Psychiatr Rehabil J*, *44*, 63–69.

258. Wilson, C., King, M., & Russell, J. (2019). A mixed-methods evaluation of a recovery college in South East Essex for people with mental health difficulties. *Health Soc Care Community*, *27*, 1353–1362.

259. Winter, L., Couwenbergh, C., & Weeghel, J. (2020). Fidelity and IPS: does quality of implementation predict vocational outcomes over time for organizations treating persons with severe mental illness in the Netherlands? *Soc Psychiatry Psychiatr Epidemiol*, *55*, 1607–1617.

260. Winter, L., Geldmacher, J., Plücker-Boss, K., & Kahl, K. G. (2020). Integration of a Return-to-Work Module in Cognitive Behavioral Therapy in Patients With Major Depressive Disorder and Long-Term Sick Leave—A Feasibility Study. *Frontiers in Psychiatry*, *11*, 512.https://doi.org/10.3389/fpsyt.2020.00512

261. Wong, D. F. K., Cheung, Y. C. H., Oades, L. G., Ye, S. S., & Ng, Y. P. (2023). Strength-based cognitive-behavioural therapy and peer-to-peer support in the recovery process for people with schizophrenia: A randomised control trial. *International Journal of Social Psychiatry*, 00207640231212096. https://doi.org/10.1177/00207640231212096

262. Worton, S., Hasford, J., & E, M. (2018). Understanding systems change in early implementation of Housing First in Canadian communities: An examination of facilitators/barriers, training/technical assistance, and points of leverage. *Am J Community Psychol*, *61*, 118–130.

263. Yamaguchi, S., Sato, S., Horio, N., Yoshida, K., Shimodaira, M., & Taneda, A. (2017). Cost-effectiveness of cognitive remediation and supported employment for people with mental illness: A randomized controlled trial. *Psychol Med*, *47*(1).

264. Yu, L., Lu, A., & M, T. (2016). Impact of integrated supported employment program on people with schizophrenia: Perspectives of participants and caregivers. *J Rehabil*, *82*, 11–17.

265. Zhang, G., Tsui, C., & A, L. (2017). Integrated supported employment for people with schizophrenia in mainland China: A randomized controlled trial. *Am J Occup Ther*, *71*(7106165020).

266. Zhu, X., Fan, H., Fan, F., Zhao, Y., Tan, Y., Yang, F., Wang, Z., Xue, F., Xiao, C., Li, W., Li, Z., Ma, L., Zou, Y., & Tan, S. (2020). Improving social functioning in community-dwelling patients with schizophrenia: A randomized controlled computer cognitive remediation therapy trial with six months follow-up. *Psychiatry Research*, *287*, 112913. https://doi.org/10.1016/j.psychres.2020.112913
